# Research Letter: Reactive balance responses after mild traumatic brain injury (mTBI): a scoping review

**DOI:** 10.1101/2020.12.17.20248433

**Authors:** Amanda Morris, Tallie Casucci, Mary M. McFarland, Benjamin Cassidy, Ryan Pelo, Nicholas Kreter, Leland E. Dibble, Peter C. Fino

**Author notes:** **Correspondence:** Amanda Morris, 250 South 1850 East, HPER Room E 107 D, Salt Lake City, UT, 84112. **Author E-mails:** TC; MMM; BC; RP; NK; LD; PF.

## Abstract

**Objective:** Balance testing after concussion or mild traumatic brain injury (mTBI) can be useful in determining acute and chronic neuromuscular deficits that are unapparent from symptom scores or cognitive testing alone. Current assessments of balance do not comprehensively evaluate all three classes of balance: maintaining a posture, voluntary movement, and reactive postural response. Despite the utility of reactive postural responses in predicting fall risk in other balance impaired populations, the effect of mTBI on reactive postural responses remains unclear. This review sought to (1) examine the extent and range of available research on reactive postural responses in people post-mTBI and (2) determine if reactive postural responses (balance recovery) are affected by mTBI.

**Design:** PRISMA Scoping review.

**Methods:** Studies were identified using Medline, Embase, CINAHL, Cochrane Library, Dissertations and Theses Global, PsycINFO, SportDiscus, and Web of Science. Inclusion criteria were: injury classified as mTBI with no confounding central or peripheral nervous system dysfunction beyond those stemming from the mTBI, quantitative measure of reactive postural response, and a discrete, externally driven perturbation was used to test reactive postural response.

**Results:** A total of 4,747 publications were identified and a total of three studies (5 publications) were included in the review.

**Conclusion:** The limited number of studies available on this topic highlight the lack of knowledge on reactive postural responses after mTBI. This review provides a new direction for balance assessments after mTBI and recommends incorporating all three classes of postural control in future research.

## 1 Introduction

Control of balance, or postural control, can be categorized into three different classes of activity: 1) maintaining posture/stability (e.g. standing), 2) voluntary transitional movements (e.g. gait), and 3) reactive postural responses (RPR) to restore stability after an unexpected disturbance ^1^. The three classes of balance are related and each involve complex interactions between motor and sensory processes ^2, 3^, but have unique neuromechanical demands.

The classes of postural control exist on a continuum of feedback and feedforward control ^4-8^ (Figure 1). Maintaining a posture requires controlling the center of mass (COM) through active postural muscle activation and integration of sensory feedback while maintaining a fixed base of support (BOS) ^8^. Unlike static posture, transitional movements involve voluntary movement of COM and changes in BOS through incorporation of pre-planning and anticipatory control based on central set ^8^, and sensory feedback for corrections ^8, 9^. In contrast to static or transitional movements, RPR involve involuntary, externally-driven motion of the COM or BOS that are corrected using rapid, time-constrained, automatic postural responses ^7, 8^ to quickly stop the initial falling motion by moving the COM or changing the BOS ^10, 11^ and stabilizing the body through subsequent postural adjustments ^8^. As RPR occur faster than voluntary movement (70-100 ms versus 180-250 ms) ^8, 12^, there is little time for pre-planning or feedback-driven control ^13^. Because of differences in neuromechanical demands, each class of postural control may yield unique, but complementary knowledge about the neuromechanical and neurophysiological clinical information regarding individuals’ postural competence.

**Figure 1.**
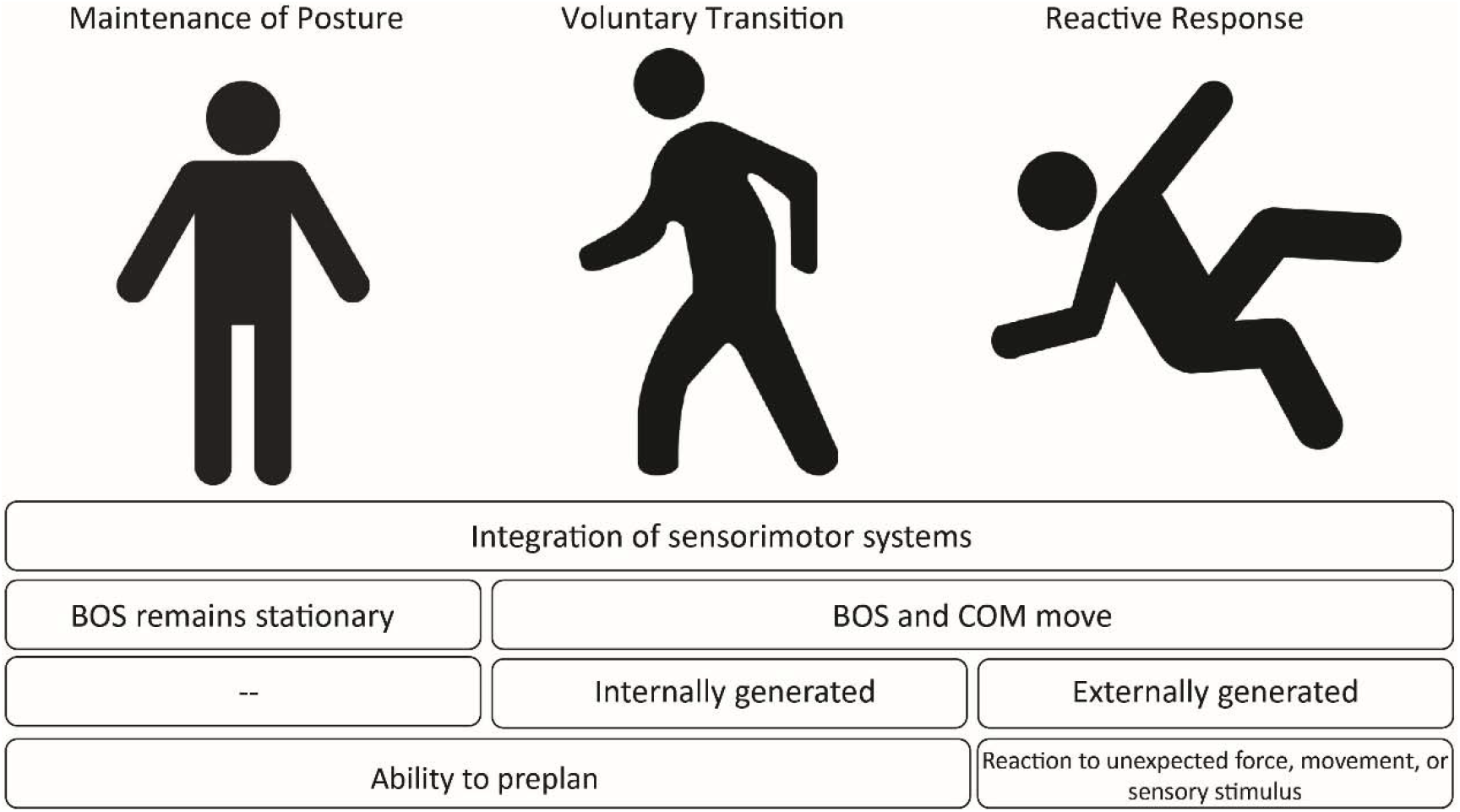
Similarities and differences between the three classes of postural control. BOS = Base of support; COM = Center of mass

Multiple reviews have concluded that concussion, hereafter mild traumatic brain injury (mTBI), impairs balance ^14-19^. The vast majority of studies on postural control after mTBI examine static posture ^20-23^ and a smaller, but significant, number of studies investigate voluntary transitional movements such as gait and turning ^24-29^. Yet, a comprehensive review of the literature surrounding RPR is lacking and the effect of mTBI on RPR remains unclear. Therefore, the purpose of this scoping review was to examine the extent and range of available research on RPR post-mTBI. We aimed to determine if RPR are affected by mTBI. Objectives, inclusion criteria, and methods for this review were specified in advance and documented in a protocol (Supplemental Digital Content 1) ^30^.

## 2 Methods

A scoping review was conducted using Arksey’s five stages ^31^ (detailed methods in Supplemental Digital Content 1). Covidence (Veritas Health Innovation, Melbourne, AU) was used to screen and select studies.

### 2.1 Inclusion Criteria

Articles were included if: 1) the injury was classified as mTBI (loss of consciousness <30 minutes, Glasgow Coma Scale >13, posttraumatic amnesia <24 hours), 2) the outcome was a measure of RPR (Supplemental Digital Content 1), 3) the perturbation was an externally driven change in support structure, visual surround, or vestibular information (sway referenced postural control tests such as the Sensory Organization Test (SOT), were not considered RPR paradigms as these perturbations were not externally driven), and 4) the perturbation was discrete.

Articles were excluded if there were confounding population factors such as: central or peripheral nervous system dysfunction beyond those stemming from the mTBI, pregnancy, or orthopedic injuries that affect postural control and gait.

### 2.2 Literature searching

Literature searching was conducted by a librarian (TC) and peer reviewed by an information specialist (MMM) with Peer Review of Electronic Search Strategies (PRESS) guidelines ^32^. The following databases were used: Medline, Embase, CINAHL, Cochrane Library, Dissertations and Theses Global, PsycINFO, SportDiscus, and Web of Science (Supplemental Digital Content 1). No date limits, nor search filters, were applied. The search was conducted January 31^st^ - February 4, 2020, and updated on March 9^th^, 2021. Search strategies were composed using database subject terms and multiple keywords for the main concepts of (brain injuries OR concussions) AND (balance) AND (posture OR postural OR perturbations) (Supplemental Digital Content 2 contains detailed search strategies). References of included studies were checked for relevancy.

Titles and abstracts were reviewed independently by four reviewers (AM, BC, RP, NK). Publications that did not meet the inclusion criteria were excluded from the review. Full-text screening and extraction of results was completed independently by three reviewers (AM, BC, RP). Approval by at least two reviewers was required for inclusion; if reviewers disagreed, all reviewers came to a consensus based on inclusion and exclusion criteria.

## 3 Results

### 3.1 Extent of Research Available

After duplicates were removed, 4,747 publications were identified (Figure 2). After title and abstract screening, 4,710 studies were excluded due to incorrect study type, population, or relevance to the aim of this review. Thirty-five full-text articles were assessed for eligibility. Of 35 articles, five articles were extracted, three of the articles ^33-35^ were from the same study (M.Walker, Personal Communication, June 5, 2020). The three selected studies and their characteristics are summarized in Table 1. Excluded studies are summarized in Supplemental Digital Content 3.

**Figure 2.**
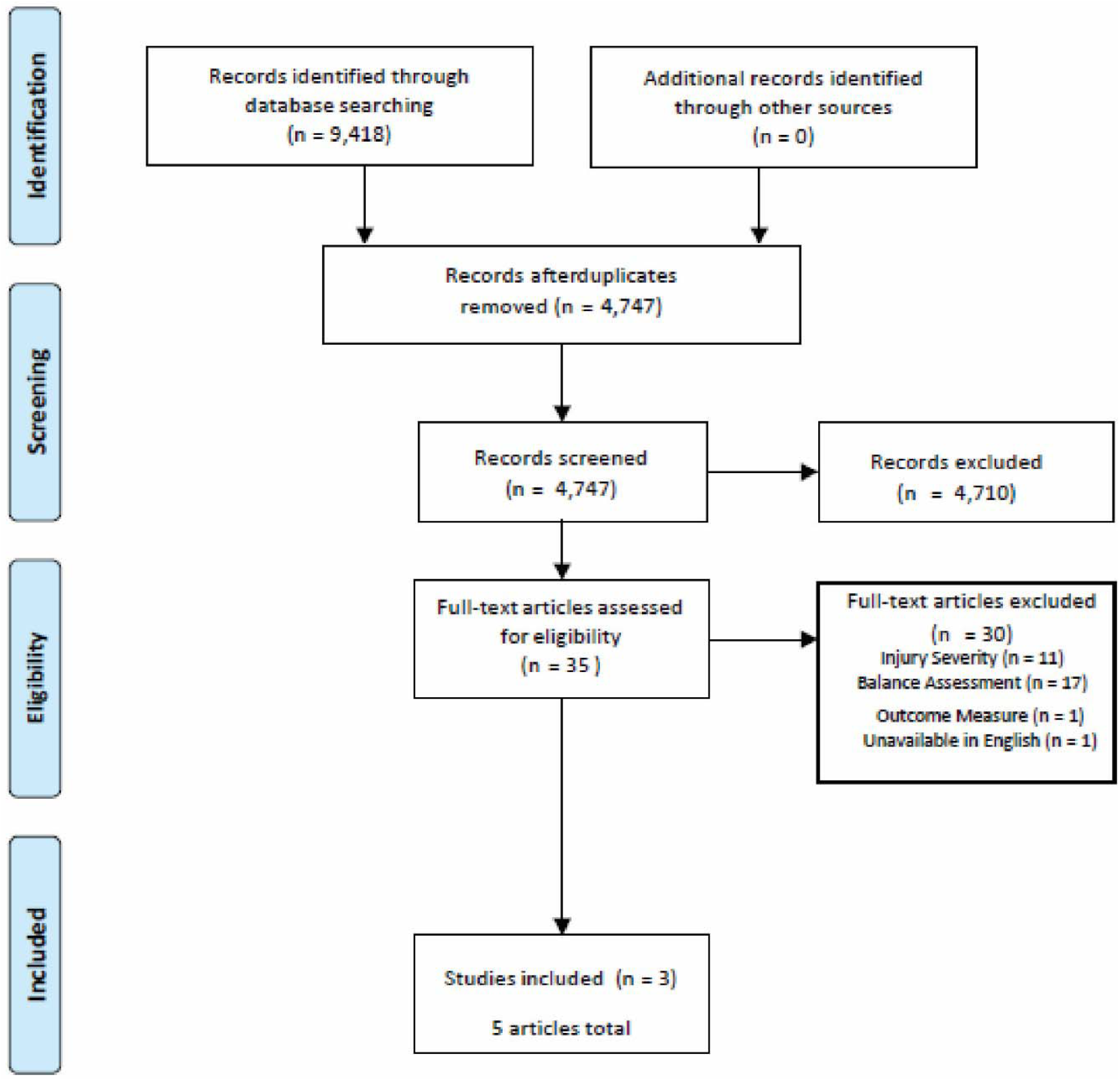
PRISMA Flow Diagram ^56^

**Table 1.**
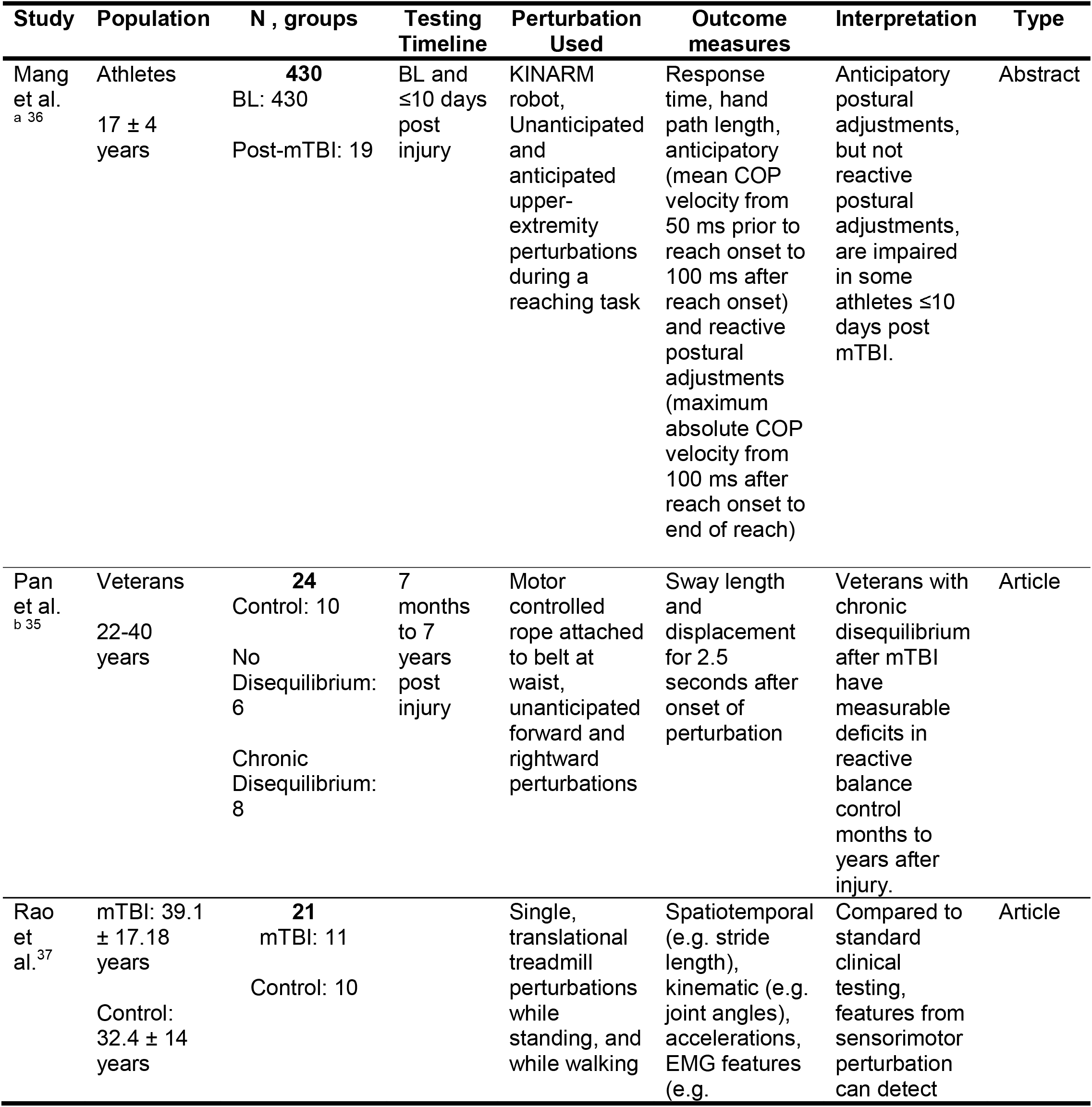
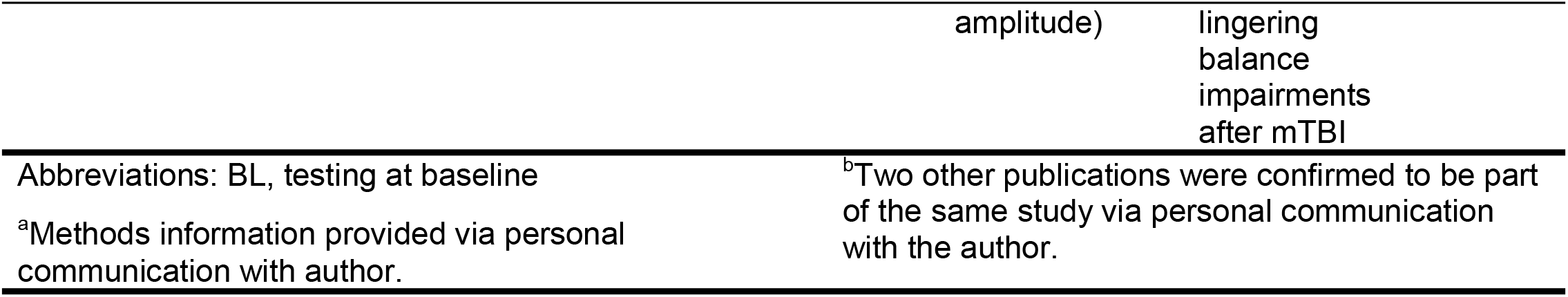
Characteristics of selected studies using discrete perturbations to examine reactive postural response after mTBI.

### 3.2 Perturbation Method and Postural Response Measures

Only three studies met our outlined criteria examining RPR to discrete perturbations after mTBI. Study one delivered expected and unexpected upper-extremity perturbations during a standing reaching task using the KINARM robot (BKIN Technologies Ltd., Kingston, ON)^36^. Perturbations were delivered after the reach started and consisted of a 12 N square pulse with a 10 second rise time applied to the left or right side of the participant (C.Mang, personal communication, June 9, 2020). Anticipatory and reactive postural adjustments were measured. RPR were quantified using the maximum absolute COP velocity from 100ms after reach onset to end of the reach.

Study two perturbed static stance by delivering unexpected forces (10% of participant’s body weight) to a waist-belt via a computer controlled motor. Perturbations were delivered for 0.5 seconds in two directions, forward and right ^35^. Motion capture was used to quantify postural sway length, displacement, and postural oscillations (frequency of marker acceleration) as RPR outcomes.

Study three perturbed static balance and walking using eight conditions on a Computer-Assisted Rehabilitation Environment (CAREN) system: 1) single, rapid (<1s) forward surface translations 2) slow rotational perturbations (platform rotated downward over 5s), 3) visual scene tilt, 4) visual scene translation, 5) visual scene congruent with tilt, 6) visual scene incongruent with tilt, 7) visual scene congruent with translation, and 8) visual scene incongruent with translation ^37^. Discrete perturbations were delivered with an interstimulus interval of 8-12 seconds. RPR outcomes were recorded using motion capture to quantify joint angles and spatiotemporal features (i.e. stride length), an accelerometer to quantify acceleration, and electromyography to record muscle activity.

### 3.3 Effect of mTBI on Reactive Postural Responses

Study 1 tested RPR in athletes at baseline and within ten days of mTBI ^36^. Before mTBI, anticipatory postural adjustments were seen only with an expected perturbation. After mTBI, anticipatory postural adjustments were seen in both expected and unexpected perturbations. However, there was no effect of mTBI on RPR.

Study 2 tested ten non-veterans without mTBI history and veterans with a history of mTBI (7 months – 7 years post mTBI)^35^. Veterans were divided into two groups, chronic complaints of imbalance (N=8) and no balance complaints (N=6). Those with chronic complaints of imbalance had significantly more upper trunk sway in response to perturbation than non-veterans and veterans without balance complaints. Additionally, those with complaints of imbalance had high-frequency postural oscillations (frequency of marker acceleration) after perturbations.

Study three tested ten healthy controls and eleven participants with mTBI who reported lingering balance deficits. Incongruent visual scene with translation or rotation, platform movement only, and visual only perturbations eliciting RPR were more sensitive than standard clinical tests (e.g.Berg Balance Scale, Dynamic Gait Index, and High Level Mobility Assessment tool, Balance Evaluations Systems Test) to detecting whether an individual reported lingering balance deficits post mTBI. Walking perturbations provided greater ability to discriminate than standing perturbations.

## 4 Conclusion

This scoping review presents the available research on RPR after mTBI. The small number of available studies prohibits a definitive conclusion. Instead, our results highlight the lack of knowledge on this topic, and demonstrate methodological variability between existing studies examining reactive postural control post–mTBI.

Two of the three studies included in this review presented results indicating that objective measures of RPR can detect lingering balance deficits after mTBI, while one study reported no change in RPR after mTBI. There was no effect of mTBI on RPR in collegiate athletes in response to upper body perturbations ^36^, nor were there reactive postural deficits in veterans who did not report disequilibrium after perturbation at the waist ^35^. However, both veterans who reported chronic imbalance after mTBI^35^ and adults with previous mTBI who reported lingering balance deficits had impaired RPR when perturbed. Therefore, it is possible that abnormal RPR are evident in individuals with chronic, persisting symptoms from mTBI, but absent in asymptomatic individuals. Impaired RPR suggest increased fall and injury risk, however, the available literature examining reactive balance deficits and their clinical consequences are severely limited and more research is needed.

The studies presented here used different methodology and populations, limiting the ability to compare results. The perturbations were different in magnitude, direction, and method; one study used right and left upper body perturbations through a hand-held apparatus, another study used a forward and rightward perturbation delivered to a waist-belt, while the third study used surface and visual perturbations. Task differences in type and perturbation profile (force and acceleration) likely resulted in different strengths of perturbation, necessitating different magnitudes of postural response ^38, 39^, making it difficult to compare the magnitude of balance deficit after mTBI between the studies. Different muscle activation patterns, joint torques, and sensory feedback may have been used depending on where the perturbation was applied or the magnitude of the perturbation. The populations also varied in age, symptom severity, and time since injury making comparisons between the three studies difficult (Table 1). It is likely that different age groups have inherent differences in RPR; there are significant differences in RPR between healthy young and old adults ^40^ and between children of different motor developmental levels ^41^. Finally, all studies tested outside of what is considered the acute period after mTBI (<48 hours) and did not perform any prospective testing, limiting the ability to determine the immediate and longitudinal effects of mTBI on RPR. Due to conflicting results, inconsistent methods, and extremely limited number of studies, the effects of mTBI on RPR to physical perturbations remains unclear.

### 4.1 Clinical Implications

Balance testing can be useful in determining acute and chronic neuromuscular impairments after mTBI that may be unapparent from symptom scores or cognitive testing alone. Studies of maintenance of posture ^21-23, 42^ and voluntary transitional movement ^17, 19, 24-26, 43, 44^ after mTBI suggest these domains of balance can be impaired both acutely and chronically after injury. Consequently, the quantification of static balance using the Balance Error Scoring System (BESS) ^45^, SOT ^46^, or dynamic balance using the tandem gait test ^47^ have become valuable clinical tools. However, these tests do not examine RPR. Given the critical importance of RPR in the prevention of falls and injury, their inclusion in the comprehensive examination of postural control after mTBI is warranted.

RPR have been used to predict fall risk in balance impaired populations such as: older adults ^48^, stroke ^49^ and Parkinson’s Disease ^50-52^. These studies, using consistent location of perturbation and systematic manipulation of perturbation magnitude, have allowed the characterization of rapid postural response to unpredictable body movement ^53^. These responses involve early automatic postural responses ^12^ and coordination of multiple body segments via neural pathways encompassing spinal, brainstem, and cortical regions ^7^. While traditionally considered a cortical injury, deficits in RPR may indicate mTBI-related dysfunction in subcortical regions and throughout the central nervous system. Increasingly, subtype-based models of care for patients with mTBI ^54, 55^ acknowledge the heterogeneous consequences of mTBI and the need for targeted, specialized treatment strategies. Future research examining RPR as well as the other classes of balance may provide more specificity than single-classed assessments of balance about the neurophysiological processes underlying motor deficits, and appropriate targets for rehabilitative care, in individuals post-mTBI.

## Supporting information

Supplemental Resource 1

Supplemental Resource 2

Supplemental Resource 3

## Data Availability

We will share any review data, including charting tables, upon request.

## 8 List of Supplemental Digital Content

**Supplemental Digital Content 1**. Scoping review protocol.

**Supplemental Digital Content 2**. Detailed search strategies. Microsoft Word Doc

**Supplemental Digital Content 3**. Table of excluded studies with study characteristics. Microsoft Word Doc

## References

1. Pollock AS, Durward BR, Rowe PJ, Paul JP. What is balance? Clinical Rehabilitation. 2000;14(4):402–406. doi:10.1191/0269215500cr342oa

2. Woollacott MH, Shumway-Cook A. Motor Control: Theory and Practical Applications. Second ed. Lippincott Williams and Wilkins; 2001:164–165.

3. Horak FB. Postural orientation and equilibrium: what do we know about neural control of balance to prevent falls? Age and Ageing. 2006;35(S2):ii7–ii11.

4. Nashner LM, Shupert CL, Horak FB, Black FO. Organization of posture controls: An analysis of sensory and mechanical constraints. Progress in Brain Research. 1989;80(C):411–418. doi:10.1016/S0079-6123(08)62237-2

5. Dietz V, Trippel M, Ibrahim IK, Berger W. Human stance on a sinusoidally translating platform: balance control by feedforward and feedback mechanisms. Experimental Brain Research. 1993;93(2):352–362. doi:10.1007/BF00228405

6. Pavol MJ, Pai Y-C. Feedforward adaptations are used to compensate for a potential loss of balance. Exp Brain Res. 2002;145:528–538. doi:10.1007/s00221-002-1143-4

7. Jacobs JV, Horak FB. Cortical control of postural responses. Journal of Neural Transmission. 2007/10// 2007;114:1339–1348. doi:10.1007/s00702-007-0657-0

8. Horak FB. Postural Control. Springer Berlin Heidelberg; 2008:3212–3219.

9. Reimann H, Fettrow T, Thompson ED, Jeka JJ. Neural Control of Balance During Walking. Frontiers in Physiology. 2018;9(SEP):1271–1271. doi:10.3389/fphys.2018.01271

10. Maki BE, Edmondstone MA, McIlroy WE. Age-related differences in laterally directed compensatory stepping behavior. Journals of Gerontology - Series A Biological Sciences and Medical Sciences. 2000;55(5):270–277. doi:10.1093/gerona/55.5.M270

11. Maki BE, McIlroy WE. Cognitive demands and cortical control of human balance-recovery reactions. Journal of Neural Transmission. 2007/10// 2007;114:1279–1296. doi:10.1007/s00702-007-0764-y

12. Horak FB, Henry SM, Shumway-Cook A. Postural Perturbations: New Insights for Treatment of Balance Disorders. Physical Therapy. 1997;77(5)

13. Rasman BG, Forbes PA, Tisserand R, Blouin JS. Sensorimotor manipulations of the balance control loop-beyond imposed external perturbations. Frontiers in Neurology. 2018;9(OCT) doi:10.3389/fneur.2018.00899

14. Guskiewicz KM. Balance Assessment in the Management of Sport-Related Concussion. Clinics in Sports Medicine. 2011;30(1):89–102. doi:10.1016/j.csm.2010.09.004

15. Valovich McLeod TC, Hale TD. Vestibular and balance issues following sport-related concussion. Brain Injury. 2015;29(2):175–184. doi:10.3109/02699052.2014.965206

16. Buckley TA, Oldham JR, Caccese JB. Postural control deficits identify lingering post-concussion neurological deficits. Journal of sport and health science. 2016;5(1):61–69. doi:10.1016/j.jshs.2016.01.007

17. Fino PC, Parrington L, Pitt W, et al. Detecting gait abnormalities after concussion or mild traumatic brain injury: A systematic review of single-task, dual-task, and complex gait. Gait and Posture. 2018;62:157–166. doi:10.1016/j.gaitpost.2018.03.021

18. Martini DN, Broglio SP. Long-term effects of sport concussion on cognitive and motor performance: A review. International Journal of Psychophysiology. 2018;132:25–30. doi:10.1016/j.ijpsycho.2017.09.019

19. Wood TA, Hsieh KL, An R, Ballard RA, Sosnoff JJ. Balance and Gait Alterations Observed More Than 2 Weeks After Concussion. American Journal of Physical Medicine & Rehabilitation. 2019;98(7):566–576. doi:10.1097/PHM.0000000000001152

20. Guskiewicz KM, Ross SE, Marshall SW. Postural Stability and Neuropsychological Deficits after Concussion in Collegiate Athletes. Journal of Athletic Training. 2001;36(3):263–273.

21. DeBeaumont L, Mongeon D, Tremblay S, et al. Persistent motor system abnormalities in formerly concussed athletes. Journal of Athletic Training. 2011;46(3):234–240. doi:10.4085/1062-6050-46.3.234

22. Quatman-Yates CC, Bonnette S, Hugentobler JA, et al. Postconcussion Postural Sway Variability Changes in Youth: The Benefit of Structural Variability Analyses. Pediatric Physical Therapy. 2015;27(4):316–327. doi:10.1097/PEP.0000000000000193

23. Fino PC, Nussbaum MA, Brolinson PG. Decreased high-frequency center-of-pressure complexity in recently concussed asymptomatic athletes. Gait and Posture. 2016;50:69–74. doi:10.1016/j.gaitpost.2016.08.026

24. Buckley TA, Munkasy BA, Tapia-Lovler TG, Wikstrom EA. Altered gait termination strategies following a concussion. Gait and Posture. 2013;38(3):549–551. doi:10.1016/j.gaitpost.2013.02.008

25. Howell DR, Osternig LR, Chou LS. Dual-task effect on gait balance control in adolescents with concussion. Archives of Physical Medicine and Rehabilitation. 2013;94(8):1513–1520. doi:10.1016/j.apmr.2013.04.015

26. Oldham JR, Munkasy BA, Evans KM, Wikstrom EA, Buckley TA. Altered dynamic postural control during gait termination following concussion. Gait and Posture. 2016;49:437–442. doi:10.1016/j.gaitpost.2016.07.327

27. Buckley T, Murray NG, Munkasy BA, Oldham JR, Evans KM, Clouse B. Impairments in Dynamic Postural Control Across Concussion Clinical Milestones. Journal of Neurotrauma. 2020:neu.2019.6910-neu.2019.6910. doi:10.1089/neu.2019.6910

28. Buckley TA, Munkasy BA, Krazeise DA, Oldham JR, Evans KM, Clouse B. Differential Effects of Acute and Multiple Concussions on Gait Initiation Performance. Archives of Physical Medicine and Rehabilitation. 2020;101(8):1347–1354. doi:10.1016/j.apmr.2020.03.018

29. Lynall RC, Campbell KR, Mauntel TC, Blackburn JT, Mihalik JP. Single-Legged Hop and Single-Legged Squat Balance Performance in Recreational Athletes With a History of Concussion. Journal of athletic training. 2020;55(5):488–493. doi:10.4085/1062-6050-185-19

30. The Joanna Briggs I. Methodology for JBI Scoping Reviews. The Joanna Briggs Institute; 2015.

31. Arksey H, O’Malley L. Scoping studies: Towards a methodological framework. International Journal of Social Research Methodology: Theory and Practice. 2005;8(1):19–32. doi:10.1080/1364557032000119616

32. McGowan J, Sampson M, Salzwedel DM, Cogo E, Foerster V, Lefebvre C. PRESS Peer Review of Electronic Search Strategies: 2015 Guideline Statement. Journal of Clinical Epidemiology. 2016;75:40–46. doi:10.1016/j.jclinepi.2016.01.021

33. Walker M, Pan T, Liao K. Dynamic postural instability in OEF/OIF veterans with mild TBI and disequilibrium. Journal of Head Trauma Rehabilitation. 2012;27(5):E34–35.

34. Walker M, Pan T, Liao K, Roenigk K, Daly J. Postural Stability in Veterans with Mild Traumatic Brain Injury and Disequilibrium. Neurology. 2015;84(14):7.161–7.161.

35. Pan T, Liao K, Roenigk K, Daly JJ, Walker MF. Static and dynamic postural stability in veterans with combat-related mild traumatic brain injury. Gait and Posture. 2015;42(4):550–557. doi:10.1016/j.gaitpost.2015.08.012

36. Mang CS, Whitten TA, Dukelow SP, Benson BW. Upper-extremity and postural responses to perturbed reaching movements for assessment of sport-related concussion. Journal of Neurotrauma. 2018;35(16):A245–A245.

37. Rao HM, Talkar T, Ciccarelli G, et al. Sensorimotor conflict tests in an immersive virtual environment reveal subclinical impairments in mild traumatic brain injury. Scientific Reports. 2020/09/08 2020;10(1):14773. doi:10.1038/s41598-020-71611-9

38. Brown LA, Jensen JL, Korff T, Woollacott MH. The translating platform paradigm: perturbation displacement waveform alters the postural response. Gait and Posture. 2001;14(3):256–263. doi:10.1016/S0966-6362(01)00131-X

39. Freyler K, Gollhofer A, Colin R, Brüderlin U, Ritzmann R. Reactive balance control in response to perturbation in unilateral stance: Interaction effects of direction, displacement and velocity on compensatory neuromuscular and kinematic responses. PLoS ONE. 2015;10(12) doi:10.1371/journal.pone.0144529

40. Thelen DG, Wojcik LA, Schultz AB, Ashton-Miller JA, Alexander NB. Age Differences in Using a Rapid Step To Regain Balance During a Forward Fall | The Journals of Gerontology: Series A | Oxford Academic. The Journals of Gerontology: Series A. 1997;52A(1):M8–M13.

41. Sundermier L, Woollacott M, Roncesvalles N, Jensen J. The development of balance control in children: Comparisons of EMG and kinetic variables and chronological and developmental groupings. Experimental Brain Research. 2001;136(3):340–350. doi:10.1007/s002210000579

42. King LA, Horak FB, Mancini M, et al. Instrumenting the balance error scoring system for use with patients reporting persistent balance problems after mild traumatic brain injury. Archives of Physical Medicine and Rehabilitation. 2014;95(2):353–359. doi:10.1016/j.apmr.2013.10.015

43. Register-Mihalik JK, Littleton AC, Guskiewicz KM. Are Divided Attention Tasks Useful in the Assessment and Management of Sport-Related Concussion? : Neuropsychol Rev; 2013. p. 300–313.

44. Kleiner M, Wong L, Dubé A, Wnuk K, Hunter SW, Graham LJ. Dual-Task Assessment Protocols in Concussion Assessment: A Systematic Literature Review. Journal of Orthopaedic and Sports Physical Therapy. 2018;48(2):87–103. doi:10.2519/jospt.2018.7432

45. Bell DR, Guskiewicz KM, Clark MA, Padua DA. Systematic review of the balance error scoring system. Sports Health. 2011;3(3):287–295. doi:10.1177/1941738111403122

46. Broglio SP, Ferrara MS, Sopiarz K, Kelly MS. Reliable change of the sensory organization test. Clinical Journal of Sport Medicine. 2008;18(2):148–154. doi:10.1097/JSM.0b013e318164f42a

47. Oldham JR, Difabio MS, Kaminski TW, Dewolf RM, Howell DR, Buckley TA. Efficacy of Tandem Gait to Identify Impaired Postural Control after Concussion. Medicine and Science in Sports and Exercise. 2018;50(6):1162–1168. doi:10.1249/MSS.0000000000001540

48. Carty C, Cronin N, Nicholson D, et al. Reactive stepping behaviour in response to forward loss of balance predicts future falls in community-dwelling older adults | Age and Ageing | Oxford Academic. Age and Ageing. 2015;44(1):109–115.

49. Mansfield A, Wong JS, McIlroy WE, et al. Do measures of reactive balance control predict falls in people with stroke returning to the community? Physiotherapy (United Kingdom). 2015;101(4):373–380. doi:10.1016/j.physio.2015.01.009

50. Valkovic□ P, Broz□ová H, Bötzel K, Ru□z□ic□ka Ee, Benetin J. Push and release test predicts better Parkinson fallers and nonfallers than the pull test: Comparison in OFF and ON medication states. Movement Disorders. 2008;23(10):1453–1457. doi:10.1002/mds.22131

51. Mak MKY, Auyeung MM. The mini-bestest can predict parkinsonian recurrent fallers: A 6-month prospective study. Journal of Rehabilitation Medicine. 2013;45(6):565–571. doi:10.2340/16501977-1144

52. Schlenstedt C, Brombacher S, Hartwigsen G, Weisser B, Möller B, Deuschl G. Comparison of the Fullerton Advanced Balance Scale, Mini-BESTest, and Berg Balance Scale to Predict Falls in Parkinson Disease. Physical Therapy. 2016;96(4):494–501. doi:10.2522/ptj.20150249

53. Maki BE, McIlroy WE. Control of rapid limb movements for balance recovery: Age-related changes and implications for fall prevention. Age and Ageing. 2006/09// 2006;35:12–18. doi:10.1093/ageing/afl078

54. Kontos AP, Collins MW, Holland CL, et al. Preliminary Evidence for Improvement in Symptoms, Cognitive, Vestibular, and Oculomotor Outcomes Following Targeted Intervention with Chronic mTBI Patients. Mil Med. Mar 1 2018;183(suppl_1):333–338. doi:10.1093/milmed/usx172

55. Möller MC, Lexell J, Wilbe Ramsay K. Effectiveness of specialized rehabilitation after mild traumatic brain injury: A systematic review and meta-analysis. J Rehabil Med. Feb 5 2021;53(2):jrm00149. doi:10.2340/16501977-2791

56. Moher D, Liberati A, Tetzlaff J, Altman DG. Preferred Reporting Items for Systematic Reviews and Meta-Analyses: The PRISMA Statement. PLoS Medicine. 2009;6(7):e1000097–e1000097. doi:10.1371/journal.pmed.1000097

