## Supplemental Resource 1 for "Research Letter: Reactive balance responses after mild traumatic brain injury (mTBI): a scoping review"

**Research question**:

Does reactive postural response (balance recovery) to a postural perturbation change with concussion?

**Secondary research questions:**

Does reactive postural response remain impaired?

***PICO model question:***

**P** Individuals diagnosed with concussion

**I** perturbations for restoring stability

**C** none

**O** Balance recovery

***PCC model question:***

***Population:*** Individuals diagnosed with mTBI or concussion (veterans, athletes, etc.) Previously, the terms concussion and mTBI have been considered synonymous ^1^. Therefore, both terms were included in the scope of this review. Accordingly, only traumatic brain injuries that were classified as mild, according to the diagnostic criteria defined by the Head Injury Interdisciplinary Special Interest Group of the American Congress of Rehabilitation Medicine, were included. Inclusion criteria for mTBI included: “1. Any period of loss of consciousness; 2. Any loss of memory for events immediately before or after the accident; 3. Any alteration in mental state at the time of the accident; and 4. Focal neurological deficit(s) that may or may not be transient; but where the severity of the injury does not exceed the following: loss of consciousness of approximately 30 minutes or less; after 30 minutes an initial Glasgow Coma Scale of 13-15; and posttraumatic amnesia not greater than 24 hours” ^2^.

***Concept:*** We considered the following definition for protocols investigating reactive postural responses: a postural response to an unexpected, external perturbation, defined as a sudden, externally driven change in support structure, visual surround, or vestibular information that takes body posture out of equilibrium ^3^. For example, an unexpected external perturbation could consist of changes in support structure such as removal of hands or a support harness (push-and-release ^4^ or lean-and-release task ^5^), translation or rotation of the surface underneath the feet ^3, 6^ that accelerates the center-of-mass (CoM) with respect to the support surface, vestibular perturbations from electrical stimulation ^7^, visual perturbation from moving visual images, or a sudden change in the support surface (i.e. slip ^8, 9^ or trip ^10^).

The above definition of reactive response protocols requires a discrete, rather than a continuous perturbation. Several studies have used continuous perturbations involving externally-driven changes in visual surround, support structure, or vestibular stimuli ^3, 11^. However, these continuous (oscillatory) perturbation paradigms commonly investigate the steady-state postural response after the body has re-weighted sensory information and reconfigured the sensory-to-motor transformation ^12-14^. These protocols more closely examine maintenance of a posture, albeit in a non-standard environment, rather than a reactive response. Discrete (single) perturbations examine the response to a single, often unexpected, disturbance and involve a rapid response without significant reweighting of sensory information. These perturbations elicit pre-programmed motor responses, triggered by multisensory integration from a primed central set, intended to serve as quick corrections to counteract the mechanical effects of the perturbation ^15^. Because of the difference in neuromechanical response to discrete and continuous perturbations and our interest in reactive postural responses to external disturbances, rather than the multisensory integration to maintain balance ^11^, only discrete perturbations were included in this review. Sway referenced postural control tests in which the tilting of the support surface or visual surround directly follows the participant’s sway, such as the Sensory Organization Test (SOT), were not considered RPR paradigms as these tests do not contain externally driven perturbations.

***Context:*** As we were trying to determine the full scope of literature available on this topic, all timeframes post-injury were included. All ages (child, adolescent, and adult) and all health care settings (veterans, community, athletics, etc.) were also included.

**Purpose of scoping study** The purpose of this scoping review is to examine the extent and range of research available on reactive postural responses post-concussion. We aim to answer whether reactive postural response (balance recovery) to a postural perturbation changes with concussion.

**Rationale for scoping study**

Concussion is the most common form of traumatic brain injury, with 1.6 to 3.8 million concussions occurring in the United States each year ^16^. The International Conference on Concussion in Sport defines concussion as a “complex pathophysiological process affecting the brain, induced by traumatic biomechanical forces” ^17^. Around 80% of concussions that present to the emergency department are classified as mild traumatic brain injury (mTBI) ^17^. There is inconsistency in terminology in concussion literature ^1^. Terms such as brain injury, concussion, mTBI, and head injury are used interchangeably; the lack of consistency in both terminology and diagnostic criteria can make comparing study results difficult ^1^. mTBI, as defined by the Head Injury Interdisciplinary Special Interest Group of the American Congress of Rehabilitation Medicine has the following criteria: “a traumatically induced physiological disruption of brain function, as manifested by one of the following: 1. Any period of loss of consciousness; 2. Any loss of memory for events immediately before or after the accident; 3. Any alteration in mental state at the time of the accident; and 4. Focal neurological deficit(s) that may or may not be transient; but where the severity of the injury does not exceed the following: loss of consciousness of approximately 30 minutes or less; after 30 minutes an initial Glasgow Coma Scale of 13-15; and posttraumatic amnesia not greater than 24 hours” ^2^. For expediency, our review will use concussion to include both mTBI and concussion.

Concussion can result in symptoms such as headache, nausea, confusion, and poor concentration ^18^. One of the most common physical signs of concussion is loss of balance ^18, 19^. Balance consists of three components: 1) maintaining posture (e.g. standing), 2) voluntary movement (e.g. gait or switching between postures), and 3) restoring stability (e.g. reaction to a push or trip) ^20^. Several studies demonstrate impairments in both standing balance ^21-23^ and gait ^24, 25^ after concussion. However, little information on reactive balance (restoring stability after a disturbance) after concussion is available. Further, clinical tests of balance after concussion such as the Balance Error Scoring System (BESS) and Sensory Organization Test (SOT), do not examine reactive balance to an external perturbation. Activities of daily living, as well as optimal athletic performance, require appropriate responses to disturbances in balance to avoid injury. Understanding how concussion affects this understudied component of balance, is crucial in developing concussion rehabilitation strategies.

A postural perturbation is a sudden, externally driven change in support structure, visual surround, or vestibular information that takes body posture out of equilibrium ^3^. For example, a perturbation could consist of changes in support structure such as removal of hands or a support harness (push-and-release ^4^ or lean-and-release task ^26^) or translation of the surface underneath the feet ^6^, vestibular perturbations from electrical stimulation ^7^, or visual perturbation from moving visual images, sudden change in the support surface ^3^. Measured responses to a postural perturbation can provide information about sensory and neuromuscular integration, nervous system performance (preparation for and adaptation to a postural disturbance), and a prediction for fall risk ^3^.

**Methods**

We will follow the methodology of scoping reviews as outlined by Arksey 2005 and expanded by Peters 2015, with Arksey's five stages: 1) identifying the research question, 2) identifying relevant studies, 3) study selection, 4) charting the data and 5) collating, summarizing and reporting the results. Using the PRISMA-ScR checklist, we will ensure best practice for transparency in reporting our scoping review methodology as outlined by Tricco 2018.

We will use Covidence (Veritas Health Innovation,) an online systematic reviewing platform to screen and select studies.

**Literature searching**

Literature searching will be conducted by a librarian (TC) and peer reviewed by an information specialist (MMM) with PRESS guidelines (McGowan 2016). Search strategy will be finalized in Ovid Medline and translated to the other databases. We will use Endnote (Clarivate Analytics) the manage citations and remove duplicates.

**Electronic searches:** The following databases will be used for electronic searches: Medline(Ovid) 1946 - 2021, Embase (embase.com) 1974 - 2021, CINAHL Complete (EBSCOhost) 1937 - 2021, Cochrane Library (wiley.com) 1898 - 2021, Dissertations & Theses Global (ProQuest) 1861 - 2021, PsycINFO (EBSCOhost) 1872 -2021, SportDiscus (EBSCOhost) 1800 - 2020, Web of Science (Clarivate Analytics) 1900 - 2021.

No date limits nor search filters will be applied.

Other sources:

We will check references of included studies for relevancy.

When data is missing or if questions arise that only authors can answer, authors will be contacted twice within 1 month.

American Society of Biomechanics abstracts from 2015 to 2019 will be hand searched by two independent reviewers (AM, BC).

**Grey Literature**

- American College of Sports Medicine (ACSM) Abstracts. Abstracts from ACSM Meetings are contained in the journal *Medicine & Science in Sport & Exercise* website, <https://journals.lww.com/acsm-msse/pages/issuelist.aspx>
- American Society of Biomechanics Abstracts, www.asbweb.org

**Study Selection (eligibility criteria)**

**Inclusion criteria**

- *Types of participants:* individuals with mTBI/Concussion regardless of time of concussion/mTBI (both acute (within 48 hours) and chronic (>3 weeks)), any sex, any age
- *Concept:* Reactive postural response/balance recovery after mTBI in response to a perturbation
  - *Perturbation:* discrete, externally driven change in support structure, visual surround, or vestibular information.
- *Context:* within the context of having mTBI in any care setting (veterans, athletes, etc.)

For example:

Perturbations such as:

- Waist belt[54] (Pan et al. 2015)Lean-and-release /Push-and-release (P&R)- removing hands or lean-and-release via cable and harness
- Moving ground/surface translations/surface tilt
- BESTest/Mini BEST – only if results for compensatory stepping are included
- Motor Control Test, discrete surface translations
- Any trip / slip paradigm : any external disturbance that would cause participant to trip or slip and have to recover their balance
- Treadmill (eg. ActiveStep by Simbex) – belt perturbation
- Vestibular or visual, discrete perturbation

Reactive postural responses such as:

- Pan et al. 2015: Trunk angle, 2D sway path, center of pressure displacement, frequency of postural oscillations[54]
- Response latency (release to step initiation), time to stabilization (time between release of support and full stabilization), heel contact latency, clinical score on P&R (number of steps)

Example Perturbations:

- Pan, T., Liao, K., Roenigk, K., Daly, J. J., & Walker, M. F. (2015). Static and dynamic postural stability in veterans with combat-related mild traumatic brain injury. *Gait & posture*, *42*(4), 550-557.

**Exclusion criteria**

- Confounding population factors: any central or peripheral nervous system dysfunction (e.g., vestibular/somatosensory pathology, Parkinson’s Disease, etc.), pregnancy, orthopedic injuries that affect balance and gait.
- Loss of consciousness > 30 min
- Glasgow Coma Scale <13
- Posttraumatic amnesia (PTA) > 24 h
- Continuous perturbation

**Quality assessment**

No quality assessment will be conducted on our selected studies.

**Strengths and Limitations**

*Limitations:* We anticipate limited information available on this review question. No quality assessment of included studies will be conducted as our goal is to rapidly map the literature.

*Strengths:* This scoping review will determine the extent to which reactive postural responses after concussion have been examined and provide rationale for future studies examining whether rehabilitation that targets reactive responses to perturbations can accelerate the recovery following concussion.

**Charting the data**

We will use Excel (Microsoft) to chart data from our selected studies.

Excel (Microsoft, Redmond, WA) will be used for data charting and the following data will be extracted in duplicate from the selected studies: author, publication year, aims/purpose, population (age if available)/sample size, testing time (time since injury), perturbation method, outcome variables, outcome and interpretation.

**Team** (list all team members - expertise in content and methodology)

Authorship clarification for manuscript

Lead Author: Amanda Morris

Author: Leland Dibble

Senior Author: Peter Fino

Reviewer: Ben Cassidy

Reviewer: Ryan Pelo

Reviewer: Nick Kreter

Librarian/Information Specialist: Tallie Casucci, Mary M. McFarland

Statistician: N/A

Project Manager (if applicable): N/A

_______

Manuscript will include these sections, not necessary for the protocol:

**Results**

Studies identified and selected (PRISMA)

**Discussion**

**Conclusions** (& implications for research or practice)

1.	McKinlay A, Bishop A, McLellan. Public knowledge of 'concussion' and the different terminology used to communicate about mild traumatic brain injury (MTBI). *Brain Injury*. 2011;25(8):761-766. doi:10.3109/02699052.2011.579935

2.	Kay T, Harrington DE, Adams R, et al. Definition of mild traumatic brain injury. *Journal of Head Trauma Rehabilitation*. 1993;8(3):86-87. doi:10.1097/00001199-199309000-00010

3.	Horak FB, Henry SM, Shumway-Cook A. Postural Perturbations: New Insights for Treatment of Balance Disorders. *Physical Therapy*. 1997;77(5)

4.	El-Gohary M, Peterson D, Gera G, Horak FB, Huisinga JM. Validity of the Instrumented Push and Release Test to Quantify Postural Responses in Persons With Multiple Sclerosis. *Archives of Physical Medicine and Rehabilitation*. 2017;98(7):1325-1331. doi:10.1016/j.apmr.2017.01.030

5.	Chan K, Lee JW, Unger J, Yoo J, Masani K, Musselman KE. Reactive stepping after a forward fall in people living with incomplete spinal cord injury or disease. *Spinal Cord*. 2019:1-9. doi:10.1038/s41393-019-0332-y

6.	Lin S-I, Woollacott MH. Postural Muscle Responses Following Changing Balance Threats in Young, Stable Older, and Unstable Older Adults. *Journal of Motor Behavior*. 2002;34(1):37-44. doi:10.1080/00222890209601929

7.	Dalton BH, Blouin JS, Allen MD, Rice CL, Inglis JT. The altered vestibular-evoked myogenic and whole-body postural responses in old men during standing. *Experimental Gerontology*. 2014;60:120-128. doi:10.1016/j.exger.2014.09.020

8.	Tang P-F, Woollacott MH. Inefficient Postural Responses to Unexpected Slips During Walking in Older Adults. *The Journals of Gerontology Series A: Biological Sciences and Medical Sciences*. 1998;53A(6):M471-M480.

9.	Liu J, Lockhart TE. Age-related joint moment characteristics during normal gait and successful reactive-recovery from unexpected slip perturbations. *Gait and Posture*. 2009;30(3):276-281. doi:10.1016/j.gaitpost.2009.04.005

10.	Bieryla KA, Madigan ML, Nussbaum MA. Practicing recovery from a simulated trip improves recovery kinematics after an actual trip. *Gait and Posture*. 2007;26(2):208-213. doi:10.1016/j.gaitpost.2006.09.010

11.	Rasman BG, Forbes PA, Tisserand R, Blouin JS. Sensorimotor manipulations of the balance control loop-beyond imposed external perturbations. *Frontiers in Neurology*. 2018;9(OCT)doi:10.3389/fneur.2018.00899

12.	Peterka RJ. Sensorimotor integration in human postural control. *J Neurophysiol*. 2002;88:1097-1118. doi:10.1152/jn.00605.2001

13.	Peterka RJ, Murchison CF, Parrington L, Fino PC, King LA. Implementation of a Central Sensorimotor Integration Test for Characterization of Human Balance Control During Stance. *Frontiers in Neurology*. 2018;9:1045-1045. doi:10.3389/fneur.2018.01045

14.	Ghai S, Nardone A, Schieppati M. Human Balance in Response to Continuous, Predictable Translations of the Support Base: Integration of Sensory Information, Adaptation to Perturbations, and the Effect of Age, Neuropathy and Parkinson’s Disease. *Applied Sciences*. 2019;9(24):5310-5310. doi:10.3390/app9245310

15.	Latash ML. *Neurophysiological Basis of Movement*. Second ed. Human Kinetics Publishers Inc.; 2008.

16.	Langlois JA, Rutland-Brown W, Wald MM. The Epidemiology and Impact of Traumatic Brain Injury: A Bri... : The Journal of Head Trauma Rehabilitation. *Journal of Head Trauma Rehabilitation*. 2006;21(5):375-378.

17.	Laker SR. Epidemiology of concussion and mild traumatic brain injury. *PM and R*. 2011;3(10 SUPPL. 2)doi:10.1016/j.pmrj.2011.07.017

18.	Aubry M, Cantu R, Dvorak J, et al. Summary and agreement statement of the first international conference on. *The Physician and sportsmedicine*. 2002;30(2):57-63. doi:10.3810/psm.2002.02.176

19.	Guskiewicz KM, Ross SE, Marshall SW. Postural Stability and Neuropsychological Deficits after Concussion in Collegiate Athletes. *Journal of Athletic Training*. 2001;36(3):263-273.

20.	Pollock AS, Durward BR, Rowe PJ, Paul JP. What is balance? *Clinical Rehabilitation*. 2000;14(4):402-406. doi:10.1191/0269215500cr342oa

21.	Quatman-Yates CC, Bonnette S, Hugentobler JA, et al. Postconcussion Postural Sway Variability Changes in Youth: The Benefit of Structural Variability Analyses. *Pediatric Physical Therapy*. 2015;27(4):316-327. doi:10.1097/PEP.0000000000000193

22.	Fino PC, Nussbaum MA, Brolinson PG. Decreased high-frequency center-of-pressure complexity in recently concussed asymptomatic athletes. *Gait and Posture*. 2016;50:69-74. doi:10.1016/j.gaitpost.2016.08.026

23.	DeBeaumont L, Mongeon D, Tremblay S, et al. Persistent motor system abnormalities in formerly concussed athletes. *Journal of Athletic Training*. 2011;46(3):234-240. doi:10.4085/1062-6050-46.3.234

24.	Fino PC, Parrington L, Pitt W, et al. Detecting gait abnormalities after concussion or mild traumatic brain injury: A systematic review of single-task, dual-task, and complex gait. Elsevier B.V.; 2018. p. 157-166.

25.	Howell DR, Osternig LR, Chou LS. Dual-task effect on gait balance control in adolescents with concussion. *Archives of Physical Medicine and Rehabilitation*. 2013;94(8):1513-1520. doi:10.1016/j.apmr.2013.04.015

26.	Chan K, Lee JW, Unger J, Yoo J, Masani K, Musselman KE. Reactive stepping after a forward fall in people living with incomplete spinal cord injury or disease. *Spinal Cord*. 2019;doi:10.1038/s41393-019-0332-y
