## Supplemental Resource 2 for "Research Letter: Reactive balance responses after mild traumatic brain injury (mTBI): a scoping review"

### Final Search Strategies

Original searches done January 31-February 4, 2020. Updated searches on March 9, 2021.

#### **Ovid Medline 2020-01-31**

1	brain injuries, traumatic/ or brain concussion/ or brain contusion/ or chronic traumatic encephalopathy/ or head injuries, closed/	16137

2	(concuss* or commotio or "dementia pugulistica" or mtbi or mtbis or post-concuss* or postconcuss* or sub-concuss* or subconcuss* or tbi or tbis or ((brain or cerebell* or cerebral or cerebrovascular or cortical or encephalopath* or head) adj3 (commotio* or concuss* or contusion* or impact* or injur* or postconcuss* or post-concuss* or posttraumatic or post-traumatic or trauma*))).ti,ab.	121761

3	1 or 2 [concussion]	123894

4	(reacti* or anticipatory or compensatory).ti,ab,kw.	1695681

5	(dynamic adj5 (balance or posture or postural)).ti,ab.	4969

6	Posture/ or (posture or postural).ti,ab.	95265

7	Postural Balance/ or kinesthesis/ or proprioception/ or Perception/ or Sensation/	79392

8	(balance or "body sway" or "deep sensitivity" or imbalance or kinesthes* or perception* or propriocept* or propriocepsis or sensation* or ((body or kinesthetic or kinaesthetic or movement or position) adj3 (discrimination or percept* or sense* or sensory or sensation*)) or ((balance or body or gait or posture or postural or musculoskeletal) adj3 (abnormal* or adaption* or adjust* or control or difficult* or disequilibrium or disturbance* or entropy or equilibrium or error or impair* or instabilit* or oscillat* or orient* or react* or recover* or response* or restor* or stabili* or stance or steadiness or steady or sway* or unsteadiness or unsteady)) or (sensory adj2 (function or functions))).ti,ab.	599516

9	or/7-8 [balance]	634004

10	ear, inner/ or cochlea/ or basilar membrane/ or cochlear aqueduct/ or cochlear duct/ or stria vascularis/ or tectorial membrane/ or "organ of corti"/ or hair cells, auditory/ or hair cells, auditory, inner/ or hair cells, auditory, outer/ or labyrinth supporting cells/ or round window, ear/ or scala tympani/ or scala vestibuli/ or spiral ganglion/ or spiral lamina/ or "spiral ligament of cochlea"/ or labyrinthine fluids/ or semicircular canals/ or semicircular ducts/ or hair cells, ampulla/ or vestibule, labyrinth/ or oval window, ear/ or "saccule and utricle"/ or acoustic maculae/ or hair cells, vestibular/ or otolithic membrane/ or vestibular aqueduct/ or endolymphatic duct/ or endolymphatic sac/	53442

11	("auris interna" or "acoustic macula*" or "basilar papilla" or cochlea* or endolymph* or "fenestra rotunda*" or "internal acoustic pore" or "internal auditory meatus" or labyrinth* or "meatus acusticus" or "macula* acoustic" or otoconia* or otolith* or "papilla acustica basilaris" or "papilla basilar" or perilymph* or "porus acusticus internus" or saccule* or sacculus or "stria vascularis" or statoconi* or statocyst or statocyte or statolith or utricle* or utriculus or "vascularis stria" or vestibul* or ((ear or ears) adj2 (helix or inner or internal or vestibul*)) or ((cochlea* or endolymphatic or perilymphatic or semicircular) adj2 duct*) or (scala adj2 (media* or tympani or tympanus or vestibuli or vestibulus)) or ((basilar or otolithic or statoconia or tectorial) adj2 (lamina or membrane*)) or ((acoustic or auditory or corti* or spiral*) adj2 (gangli* or lamina or laminas or ligament or ligaments or organ or organs or organum)) or ((ampullar* or auditory or cochlea* or inner or outer or vestibular) adj2 ("hair cell" or "hair cells")) or ((auditory or cochlea* or ear or semicircular or vestibul*) adj2 (apparatus or aqueduct* or auris or canal or canals or canaliculus or "oval window*" or "round window*")) or (ampullar* adj2 (crest or crests or crista))).ti,ab.	114696

12	10 or 11 [inner ear anatomy]	123751

13	sensorimotor cortex/ or motor cortex/ or somatosensory cortex/ or visual cortex/ or (("anterior parietal" or Brodmann or extrastriat* or motor or motorsensory or premotor or "post central" or postcentral* or precentral* or "pre central" or Rolandic or sensory or sensorimotor or "sensory motor" or "sensory-motor" or si or sensomotor or somatomotor or somatosensory or "somatic sensory" or striate or visual) adj3 (area or areas or cortex or cortices or gyrus or region or strip or strips or zone)).ti,ab.	98834

14	Feedback, Sensory/ or ((acoustic* or audio* or haptic or propriocept* or sensorimotor or sensory or tactile or visual) adj3 (feedback or feedbacks)).ti,ab.	10857

15	sensory gating/ or prepulse inhibition/ or ((sensory or sensorimotor or startle) adj3 (gating or filtering)).ti,ab. or ((prepulse or pre-pulse) adj2 (facilitate* or inhibit*)).ti,ab.	4923

16	vestibular diseases/ or bilateral vestibulopathy/ or vertigo/ or benign paroxysmal positional vertigo/ or Vestibular Nerve/ or Vestibular Neuronitis/ or (Neurolabyrinthiti* or "Scarpa Ganglion*" or "Scarpas Ganglion*" or "Scarpa's Ganglion*" or vestibul* or vertigo*).ti,ab.	57842

17	or/13-16 [sensorimotor or vestibular]	170105

18	9 or 12 or 17 [large balance set]	848162

19	perturbation*.ti,ab,kw.	73767

20	(ActiveStep or BESTest or CAREN or "Computer Assisted Rehabilitation Environment" or "center of pressure" or "centre of pressure" or "central sensorimotor integration task" or GVS or "galvanic vestibular stimulation" or "heel contact latenc*" or "lean-and-release" or "lean and release" or "lean & release" or "limits of stability" or "mini BEST" or miniBEST or (motion adj3 (track* or control* or test*)) or MCT or "motor control test" or neurocom or posturograph* or "Push-and-release" or "push and release" or "push & release" or P&R or "P and R" or "release-from-lean" or "release from lean" or Simbex or "SMART EquiTest" or "stance slip" or SVS or "vestibular stimulation" or "support surface" or (trunk adj5 (angle or sway)) or VEMP or "vestibular evoked myogenic potential*" or vibrat* or "waist belt" or (waist adj5 belt)).ti,ab,kf. or ((ground or forceplate or "force plate" or platform or surface) adj5 (moving or tilt or translation*)).ti,ab. [specific tests]	168411

21	(("virtual reality" or vr) adj3 (balance or propriocept* or posture or postural)).ti,ab.	124

22	((balance or gait or perturbation* or postural or posture or propriocept* or sensorimotor* or slip or slipping or stance or stand or standing or step or stepping or trip or tripping or walk or walking) adj5 (assess* or dual-task or "dual task" or simulat* or task or tasks or test or tests or testing)).ti,ab.	59090

23	or/19-22 [large perturbation set]	291451

24	3 and 19 and (balance or posture or postural).ti,ab,kw. [concussion + perturbation + balance or posture]	35

25	3 and 5 [concussion + dynamic balance]	100

26	3 and 4 and 18 [concussion + reactive + large balance]	631

27	3 and 18 and 23 [concussion + large balance + large perturbation]	1017

28	3 and 22 [concussion + balance tests]	1303

29	3 and 6 and 9 [concussion + posture + balance]	466

30	or/24-29 [final set]	2197

31	(exp animals/ not humans/) or (animal? or beaver? or beef or bovine or breeding or bull or canine or castoris or cat or cattle or cats or chicken? or chimp* or cow or dog or dogs or equine or foal or foals or fish or insect? or horse or horses or livestock or mice or monkey? or mouse or murine or plant or plants or pork or porcine or protozoa? or purebred or rabbit or rabbits or rat or rats or rodent? or sheep or thoroughbred).ti. or veterinar$.ti,ab.	5224509

32	30 not 31 [all no animals]	1528

33	("26374930" or "27580081" or "26397071" or "21669091" or "23643687").ui. [exemplars]	5

34	32 or 33 [final set includes exemplars]	1528

------

Re-ran search on March 9, 2021

35	limit 34 to yr="2020 -Current"	296

#### **Embase (Embase.com) 2020-01-31**

#34	#32 NOT #33	2294

#33	'animal'/exp NOT 'human'/exp OR animal:ti OR animals:ti OR beaver:ti OR beavers:ti OR beef:ti OR bovine:ti OR breeding:ti OR bull:ti OR canine:ti OR castoris:ti OR cat:ti OR cattle:ti OR cats:ti OR chicken:ti OR chimp*:ti OR cow:ti OR dog:ti OR dogs:ti OR equine:ti OR foal:ti OR foals:ti OR fish:ti OR insect:ti OR insects:ti OR horse:ti OR horses:ti OR livestock:ti OR mice:ti OR monkey:ti OR monkeys:ti OR mouse:ti OR murine:ti OR plant:ti OR plants:ti OR pork:ti OR porcine:ti OR protozoa:ti OR purebred:ti OR rabbit:ti OR rabbits:ti OR rat:ti OR rats:ti OR rodent:ti OR rodents:ti OR sheep:ti OR thoroughbred:ti OR veterinar*:ti,ab	6001858

#32	#26 OR #27 OR #28 OR #29 OR #30 OR #31	3332

#31	#3 AND #6 AND #9	611

#30	#3 AND #24	1975

#29	#3 AND #20 AND #25	1446

#28	#3 AND #4 AND #20	973

#27	#3 AND #5	169

#26	#3 AND #21 AND (balance:ti,ab,kw OR posture:ti,ab,kw OR postural:ti,ab,kw)	51

#25	#21 OR #22 OR #23 OR #24	262339

#24	((balance OR gait OR perturbation* OR postural OR posture OR propriocept* OR sensorimotor* OR slip OR slipping OR stance OR stand OR standing OR step OR stepping OR trip OR tripping OR walk OR walking) NEAR/5 (assess* OR 'dual task' OR 'dual task' OR simulat* OR task OR tasks OR test OR tests OR testing)):ti,ab	86020

#23	(('virtual reality' OR vr) NEAR/3 (balance OR propriocept* OR posture OR postural)):ti,ab	196

#22	activestep:ti,ab,kw OR bestest:ti,ab,kw OR caren:ti,ab,kw OR 'computer assisted rehabilitation environment':ti,ab,kw OR 'center of pressure':ti,ab,kw OR 'centre of pressure':ti,ab,kw OR 'central sensorimotor integration task':ti,ab,kw OR gvs:ti,ab,kw OR 'galvanic vestibular stimulation':ti,ab,kw OR 'heel contact latenc*':ti,ab,kw OR 'limits of stability':ti,ab,kw OR 'mini best':ti,ab,kw OR minibest:ti,ab,kw OR ((motion NEAR/3 (track* OR control* OR test*)):ti,ab,kw) OR mct:ti,ab,kw OR 'motor control test':ti,ab,kw OR neurocom:ti,ab,kw OR posturograph*:ti,ab,kw OR (((push OR lean) NEAR/2 release):ti,ab,kw) OR p&r:ti,ab,kw OR 'release-from-lean':ti,ab,kw OR 'release from lean':ti,ab,kw OR simbex:ti,ab,kw OR 'smart equitest':ti,ab,kw OR 'stance slip':ti,ab,kw OR svs:ti,ab,kw OR 'vestibular stimulation':ti,ab,kw OR 'support surface':ti,ab,kw OR ((trunk NEAR/5 (angle OR sway)):ti,ab,kw) OR vemp:ti,ab,kw OR 'vestibular evoked myogenic potential*':ti,ab,kw OR vibrat*:ti,ab,kw OR 'waist belt':ti,ab,kw OR ((waist NEAR/5 belt):ti,ab,kw) OR (((ground OR forceplate OR 'force plate' OR platform OR surface) NEAR/5 (moving OR tilt OR translation*)):ti,ab)	110608

#21	perturbation*:ti,ab,kw	76053

#20	#9 OR #12 OR #19	2368872

#19	#13 OR #14 OR #15 OR #16 OR #17 OR #18	196993

#18	neurolabyrinthiti*:ti,ab OR 'scarpa ganglion*':ti,ab OR 'scarpas ganglion*':ti,ab OR vestibul*:ti,ab OR vertigo*:ti,ab	66555

#17	'vestibular disorder'/mj OR 'bilateral vestibulopathy'/mj OR 'vertigo'/mj OR 'benign paroxysmal positional vertigo'/mj OR 'canalolithiasis'/mj OR 'cupulolithiasis'/mj OR 'positional vertigo'/mj OR 'vestibular nerve'/mj OR 'vestibular neuronitis'/mj	21651

#16'sensory gating'/mj OR 'prepulse inhibition'/mj OR (((sensory OR sensorimotor OR startle) NEAR/3 (gating OR filtering)):ti,ab) OR (((prepulse OR 'pre pulse') NEAR/2 (facilitate* OR inhibit*)):ti,ab)	6100

#15	'sensory feedback'/mj OR 'proprioceptive feedback'/mj OR 'tactile feedback'/mj OR 'auditory feedback'/mj OR 'visual feedback'/mj OR (((acoustic* OR audio* OR haptic OR propriocept* OR sensorimotor OR sensory OR tactile OR visual) NEAR/3 (feedback OR feedbacks)):ti,ab)	12041

#14	(('anterior parietal' OR brodmann OR extrastriat* OR motor OR motorsensory OR premotor OR 'post central' OR postcentral* OR precentral* OR 'pre central' OR rolandic OR sensory OR sensorimotor OR 'sensory motor' OR 'sensory-motor' OR si OR sensomotor OR somatomotor OR somatosensory OR 'somatic sensory' OR striate OR visual) NEAR/3 (area OR areas OR cortex OR cortices OR gyrus OR region OR strip OR strips OR zone)):ti,ab	99712

#13	'sensorimotor cortex'/mj OR 'motor cortex'/mj OR 'premotor cortex'/mj OR 'primary motor cortex'/mj OR 'supplementary motor area'/mj OR 'somatosensory cortex'/mj OR 'primary somatosensory cortex'/mj OR 'secondary somatosensory cortex'/mj OR 'visual cortex'/mj	32537

#12	#10 OR #11	138061

#11	'auris interna':ti,ab OR 'acoustic macula*':ti,ab OR 'basilar papilla':ti,ab OR cochlea*:ti,ab OR endolymph*:ti,ab OR 'fenestra rotunda*':ti,ab OR 'internal acoustic pore':ti,ab OR 'internal auditory meatus':ti,ab OR labyrinth*:ti,ab OR 'meatus acusticus':ti,ab OR 'macula* acoustic':ti,ab OR otoconia*:ti,ab OR otolith*:ti,ab OR 'papilla acustica basilaris':ti,ab OR 'papilla basilar':ti,ab OR perilymph*:ti,ab OR 'porus acusticus internus':ti,ab OR saccule*:ti,ab OR sacculus:ti,ab OR 'stria vascularis':ti,ab OR statoconi*:ti,ab OR statocyst:ti,ab OR statocyte:ti,ab OR statolith:ti,ab OR utricle*:ti,ab OR utriculus:ti,ab OR 'vascularis stria':ti,ab OR vestibul*:ti,ab OR (((ear OR ears) NEAR/2 (helix OR inner OR internal OR vestibul*)):ti,ab) OR (((cochlea* OR endolymphatic OR perilymphatic OR semicircular) NEAR/2 duct*):ti,ab) OR ((scala NEAR/2 (media* OR tympani OR tympanus OR vestibuli OR vestibulus)):ti,ab) OR (((basilar OR otolithic OR statoconia OR tectorial) NEAR/2 (lamina OR membrane*)):ti,ab) OR (((acoustic OR auditory OR corti* OR spiral*) NEAR/2 (gangli* OR lamina OR laminas OR ligament OR ligaments OR organ OR organs OR organum)):ti,ab) OR (((ampullar* OR auditory OR cochlea* OR inner OR outer OR vestibular) NEAR/2 ('hair cell' OR 'hair cells')):ti,ab) OR (((auditory OR cochlea* OR ear OR semicircular OR vestibul*) NEAR/2 (apparatus OR aqueduct* OR auris OR canal OR canals OR canaliculus OR 'oval window*' OR 'round window*')):ti,ab) OR ((ampullar* NEAR/2 (crest OR crests OR crista)):ti,ab)	133800

#10	'inner ear'/mj OR 'cochlea'/mj OR 'basilar membrane'/mj OR 'cochlea aqueduct'/mj OR 'cochlea duct'/mj OR 'cochlea fenestra'/mj OR 'corti organ'/mj OR 'labyrinth supporting cells'/mj OR 'scala tympani'/mj OR 'scala vestibuli'/mj OR 'spiral ligament'/mj OR 'stria vascularis'/mj OR 'tectorial membrane'/mj OR 'hair cell'/mj OR 'cochlear hair cell'/mj OR 'inner hair cell'/mj OR 'outer hair cell'/mj OR 'vestibular hair cell'/mj OR 'type 1 vestibular hair cell'/mj OR 'type 2 vestibular hair cell'/mj OR 'internal auditory canal'/mj OR 'vestibular labyrinth'/mj OR 'endolymph'/mj OR 'endolymphatic duct'/mj OR 'endolymphatic sac'/mj OR 'otolith'/mj OR 'otolithic membrane'/mj OR 'perilymph'/mj OR 'saccule'/mj OR 'semicircular canal'/mj OR 'semicircular duct'/mj OR 'utricle'/mj OR 'vestibule'/mj OR 'oval window'/mj OR 'vestibule aqueduct'/mj	32984

#9	#7 OR #8	773016

#8	balance:ti,ab OR 'body sway':ti,ab OR 'deep sensitivity':ti,ab OR imbalance:ti,ab OR kinesthes*:ti,ab OR perception*:ti,ab OR propriocept*:ti,ab OR propriocepsis:ti,ab OR sensation*:ti,ab OR (((body OR kinesthetic OR kinaesthetic OR movement OR position) NEAR/3 (discrimination OR percept* OR sense* OR sensory OR sensation*)):ti,ab) OR (((balance OR body OR gait OR posture OR postural OR musculoskeletal) NEAR/3 (abnormal* OR adaption* OR adjust* OR control OR difficult* OR disequilibrium OR disturbance* OR entropy OR equilibrium OR error OR impair* OR instabilit* OR oscillat* OR orient* OR react* OR recover* OR response* OR restor* OR stabili* OR stance OR steadiness OR steady OR sway* OR unsteadiness OR unsteady)):ti,ab) OR ((sensory NEAR/2 (function OR functions)):ti,ab)	770231

#7	'body equilibrium'/mj OR 'proprioception'/mj OR 'kinesthesia'/mj OR 'balance impairment'/mj OR 'balance disorder'/mj	15006

#6	'body position'/mj OR posture:ti,ab OR postural:ti,ab	82186

#5	(dynamic NEAR/5 (balance OR posture OR postural)):ti,ab	6409

#4	reacti*:ti,ab,kw OR anticipatory:ti,ab,kw OR compensatory:ti,ab,kw	2102282

#3	#1 OR #2	59606

#2	concuss*:ti,ab OR commotio:ti,ab OR 'dementia pugulistica':ti,ab OR mtbi:ti,ab OR mtbis:ti,ab OR 'post concuss*':ti,ab OR postconcuss*:ti,ab OR 'sub concuss*':ti,ab OR subconcuss*:ti,ab OR tbi:ti,ab OR tbis:ti,ab OR (((brain OR cerebell* OR cerebral OR cerebrovascular OR cortical OR encephalopath* OR head) NEAR/3 (commotio* OR concuss* OR contusion* OR impact* OR injur* OR postconcuss* OR 'post concuss*' OR posttraumatic OR 'post traumatic' OR trauma*)):ti,ab)	171096

#1	'concussion'/mj OR 'brain concussion'/mj OR 'traumatic brain injury'/mj OR 'pediatric traumatic brain injury'/mj OR 'chronic traumatic encephalopathy'/mj OR 'brain contusion'/mj OR 'postconcussion syndrome'/mj	35537

------

Re-ran search on March 9, 2021

#35	#34 AND (2020:py OR 2021:py)	310

#### **CINAHL Complete (Ebscohost) 2020-02-01**

| **#** | **Query** | **Limiters/Expanders** | **Last Run Via** | **Results** |
| --- | --- | --- | --- | --- |
| S32 | S30 NOT S31 | Expanders - Apply equivalent subjects  Search modes - Boolean/Phrase | Interface - EBSCOhost Research Databases  Search Screen - Basic Search  Database - CINAHL Complete | 945 |
| S31 | (MH "Animals+") NOT (MH "Human") | Expanders - Apply equivalent subjects  Search modes - Boolean/Phrase | Interface - EBSCOhost Research Databases  Search Screen - Advanced Search  Database - CINAHL Complete | Display |
| S30 | S24 OR S25 OR S26 OR S27 OR S28 OR S29 | Expanders - Apply equivalent subjects  Search modes - Boolean/Phrase | Interface - EBSCOhost Research Databases  Search Screen - Advanced Search  Database - CINAHL Complete | Display |
| S29 | S3 AND S6 AND S9 | Expanders - Apply equivalent subjects  Search modes - Boolean/Phrase | Interface - EBSCOhost Research Databases  Search Screen - Advanced Search  Database - CINAHL Complete | Display |
| S28 | S3 AND S22 | Expanders - Apply equivalent subjects  Search modes - Boolean/Phrase | Interface - EBSCOhost Research Databases  Search Screen - Advanced Search  Database - CINAHL Complete | Display |
| S27 | S3 AND S18 AND S23 | Expanders - Apply equivalent subjects  Search modes - Boolean/Phrase | Interface - EBSCOhost Research Databases  Search Screen - Advanced Search  Database - CINAHL Complete | Display |
| S26 | S3 AND S4 AND S18 | Expanders - Apply equivalent subjects  Search modes - Boolean/Phrase | Interface - EBSCOhost Research Databases  Search Screen - Advanced Search  Database - CINAHL Complete | Display |
| S25 | S3 AND S5 | Expanders - Apply equivalent subjects  Search modes - Boolean/Phrase | Interface - EBSCOhost Research Databases  Search Screen - Advanced Search  Database - CINAHL Complete | Display |
| S24 | S3 AND S19 AND (TI (balance or posture or postural) OR AB (balance or posture or postural)) | Expanders - Apply equivalent subjects  Search modes - Boolean/Phrase | Interface - EBSCOhost Research Databases  Search Screen - Advanced Search  Database - CINAHL Complete | Display |
| S23 | S19 OR S20 OR S21 OR S22 | Expanders - Apply equivalent subjects  Search modes - Boolean/Phrase | Interface - EBSCOhost Research Databases  Search Screen - Advanced Search  Database - CINAHL Complete | Display |
| S22 | TI ( ((balance or gait or perturbation* or postural or posture or propriocept* or sensorimotor* or slip or slipping or stance or stand or standing or step or stepping or trip or tripping or walk or walking) N5 (assess* or dual-task or "dual task" or simulat* or task or tasks or test or tests or testing)) ) OR AB ( ((balance or gait or perturbation* or postural or posture or propriocept* or sensorimotor* or slip or slipping or stance or stand or standing or step or stepping or trip or tripping or walk or walking) N5 (assess* or dual-task or "dual task" or simulat* or task or tasks or test or tests or testing)) ) | Expanders - Apply equivalent subjects  Search modes - Boolean/Phrase | Interface - EBSCOhost Research Databases  Search Screen - Advanced Search  Database - CINAHL Complete | Display |
| S21 | TI ( (("virtual reality" or vr) N3 (balance or propriocept* or posture or postural)) ) OR AB ( (("virtual reality" or vr) N3 (balance or propriocept* or posture or postural)) ) | Expanders - Apply equivalent subjects  Search modes - Boolean/Phrase | Interface - EBSCOhost Research Databases  Search Screen - Advanced Search  Database - CINAHL Complete | Display |
| S20 | TI ( ActiveStep or BESTest or CAREN or "Computer Assisted Rehabilitation Environment" or "center of pressure" or "centre of pressure" or "central sensorimotor integration task" or GVS or "galvanic vestibular stimulation" or "heel contact latenc*" or "lean-and-release" or "lean and release" or "lean & release" or "limits of stability" or "mini BEST" or miniBEST or (motion N3 (track* or control* or test*)) or MCT or "motor control test" or neurocom or posturograph* or "Push-and-release" or "push and release" or "push & release" or P&R or "P and R" or "release-from-lean" or "release from lean" or Simbex or "SMART EquiTest" or "stance slip" or SVS or "vestibular stimulation" or "support surface" or (trunk N5 (angle or sway)) or VEMP or "vestibular evoked myogenic potential*" or vibrat* or "waist belt" or (waist N5 belt) or ((ground or forceplate or "force plate" or platform or surface) N5 (moving or tilt or translation*)) ) OR AB ( ActiveStep or BESTest or CAREN or "Computer Assisted Rehabilitation Environment" or "center of pressure" or "centre of pressure" or "central sensorimotor integration task" or GVS or "galvanic vestibular stimulation" or "heel contact latenc*" or "lean-and-release" or "lean and release" or "lean & release" or "limits of stability" or "mini BEST" or miniBEST or (motion N3 (track* or control* or test*)) or MCT or "motor control test" or neurocom or posturograph* or "Push-and-release" or "push and release" or "push & release" or P&R or "P and R" or "release-from-lean" or "release from lean" or Simbex or "SMART EquiTest" or "stance slip" or SVS or "vestibular stimulation" or "support surface" or (trunk N5 (angle or sway)) or VEMP or "vestibular evoked myogenic potential*" or vibrat* or "waist belt" or (waist N5 belt) or ((ground or forceplate or "force plate" or platform or surface) N5 (moving or tilt or translation*)) ) | Expanders - Apply equivalent subjects  Search modes - Boolean/Phrase | Interface - EBSCOhost Research Databases  Search Screen - Advanced Search  Database - CINAHL Complete | Display |
| S19 | TI perturbation* OR AB perturbation* | Expanders - Apply equivalent subjects  Search modes - Boolean/Phrase | Interface - EBSCOhost Research Databases  Search Screen - Advanced Search  Database - CINAHL Complete | Display |
| S18 | S9 OR S12 OR S17 | Expanders - Apply equivalent subjects  Search modes - Boolean/Phrase | Interface - EBSCOhost Research Databases  Search Screen - Advanced Search  Database - CINAHL Complete | Display |
| S17 | S13 OR S14 OR S15 OR S16 | Expanders - Apply equivalent subjects  Search modes - Boolean/Phrase | Interface - EBSCOhost Research Databases  Search Screen - Advanced Search  Database - CINAHL Complete | Display |
| S16 | TI ( (Neurolabyrinthiti* or "Scarpa Ganglion*" or "Scarpas Ganglion*" or "Scarpa's Ganglion*" or vestibul* or vertigo*) ) OR AB ( (Neurolabyrinthiti* or "Scarpa Ganglion*" or "Scarpas Ganglion*" or "Scarpa's Ganglion*" or vestibul* or vertigo*) ) | Expanders - Apply equivalent subjects  Search modes - Boolean/Phrase | Interface - EBSCOhost Research Databases  Search Screen - Advanced Search  Database - CINAHL Complete | Display |
| S15 | TI ( ((sensory or sensorimotor or startle) N3 (gating or filtering)) or ((prepulse or pre-pulse) N2 facilitate* or inhibit*)) ) OR AB ( ((sensory or sensorimotor or startle) N3 (gating or filtering)) or ((prepulse or pre-pulse) N2 (facilitate* or inhibit*)) ) | Expanders - Apply equivalent subjects  Search modes - Boolean/Phrase | Interface - EBSCOhost Research Databases  Search Screen - Advanced Search  Database - CINAHL Complete | Display |
| S14 | TI ( (acoustic* or audio* or haptic or propriocept* or sensorimotor or sensory or tactile or visual) N3 (feedback or feedbacks)) ) OR AB ( (acoustic* or audio* or haptic or propriocept* or sensorimotor or sensory or tactile or visual) N3 (feedback or feedbacks)) ) | Expanders - Apply equivalent subjects  Search modes - Boolean/Phrase | Interface - EBSCOhost Research Databases  Search Screen - Advanced Search  Database - CINAHL Complete | Display |
| S13 | TI ( ("anterior parietal" or Brodmann or extrastriat* or motor or motorsensory or premotor or "post central" or postcentral* or precentral* or "pre central" or Rolandic or sensory or sensorimotor or "sensory motor" or "sensory-motor" or si or sensomotor or somatomotor or somatosensory or "somatic sensory" or striate or visual) N3 (area or areas or cortex or cortices or gyrus or region or strip or strips or zone)) ) OR AB ( ("anterior parietal" or Brodmann or extrastriat* or motor or motorsensory or premotor or "post central" or postcentral* or precentral* or "pre central" or Rolandic or sensory or sensorimotor or "sensory motor" or "sensory-motor" or si or sensomotor or somatomotor or somatosensory or "somatic sensory" or striate or visual) N3 (area or areas or cortex or cortices or gyrus or region or strip or strips or zone)) ) | Expanders - Apply equivalent subjects  Search modes - Boolean/Phrase | Interface - EBSCOhost Research Databases  Search Screen - Advanced Search  Database - CINAHL Complete | Display |
| S12 | S10 OR S11 | Expanders - Apply equivalent subjects  Search modes - Boolean/Phrase | Interface - EBSCOhost Research Databases  Search Screen - Advanced Search  Database - CINAHL Complete | Display |
| S11 | TI ( ("auris interna" or "acoustic macula*" or "basilar papilla" or cochlea* or endolymph* or "fenestra rotunda*" or "internal acoustic pore" or "internal auditory meatus" or labyrinth* or "meatus acusticus" or "macula* acoustic" or otoconia* or otolith* or "papilla acustica basilaris" or "papilla basilar" or perilymph* or "porus acusticus internus" or saccule* or sacculus or "stria vascularis" or statoconi* or statocyst or statocyte or statolith or utricle* or utriculus or "vascularis stria" or vestibul* or ((ear or ears) N2 (helix or inner or internal or vestibul*)) or ((cochlea* or endolymphatic or perilymphatic or semicircular) N2 duct*) or (scala N2 (media* or tympani or tympanus or vestibuli or vestibulus)) or ((basilar or otolithic or statoconia or tectorial) N2 (lamina or membrane*)) or ((acoustic or auditory or corti* or spiral*) N2 (gangli* or lamina or laminas or ligament or ligaments or organ or organs or organum)) or ((ampullar* or auditory or cochlea* or inner or outer or vestibular) N2 ("hair cell" or "hair cells")) or ((auditory or cochlea* or ear or semicircular or vestibul*) N2 (apparatus or aqueduct* or auris or canal or canals or canaliculus or "oval window*" or "round window*")) or (ampullar* N2 (crest or crests or crista))) ) OR AB ( ("auris interna" or "acoustic macula*" or "basilar papilla" or cochlea* or endolymph* or "fenestra rotunda*" or "internal acoustic pore" or "internal auditory meatus" or labyrinth* or "meatus acusticus" or "macula* acoustic" or otoconia* or otolith* or "papilla acustica basilaris" or "papilla basilar" or perilymph* or "porus acusticus internus" or saccule* or sacculus or "stria vascularis" or statoconi* or statocyst or statocyte or statolith or utricle* or utriculus or "vascularis stria" or vestibul* or ((ear or ears) N2 (helix or inner or internal or vestibul*)) or ((cochlea* or endolymphatic or perilymphatic or semicircular) N2 duct*) or (scala N2 (media* or tympani or tympanus or vestibuli or vestibulus)) or ((basilar or otolithic or statoconia or tectorial) N2 (lamina or membrane*)) or ((acoustic or auditory or corti* or spiral*) N2 (gangli* or lamina or laminas or ligament or ligaments or organ or organs or organum)) or ((ampullar* or auditory or cochlea* or inner or outer or vestibular) N2 ("hair cell" or "hair cells")) or ((auditory or cochlea* or ear or semicircular or vestibul*) N2 (apparatus or aqueduct* or auris or canal or canals or canaliculus or "oval window*" or "round window*")) or (ampullar* N2 (crest or crests or crista))) ) | Expanders - Apply equivalent subjects  Search modes - Boolean/Phrase | Interface - EBSCOhost Research Databases  Search Screen - Advanced Search  Database - CINAHL Complete | Display |
| S10 | (MH "Ear, Inner") OR (MH "Cochlea") OR (MH "Basilar Membrane") OR (MH "Hair Cells") OR (MH "Semicircular Canals") OR (MH "Vestibule, Labyrinth") OR (MH "Vestibular Aqueduct") OR (MH "Endolymphatic Duct") OR (MH "Endolymphatic Sac") | Expanders - Apply equivalent subjects  Search modes - Boolean/Phrase | Interface - EBSCOhost Research Databases  Search Screen - Advanced Search  Database - CINAHL Complete | Display |
| S9 | S7 OR S8 | Expanders - Apply equivalent subjects  Search modes - Boolean/Phrase | Interface - EBSCOhost Research Databases  Search Screen - Advanced Search  Database - CINAHL Complete | Display |
| S8 | TI ( (balance or "body sway" or "deep sensitivity" or imbalance or kinesthes* or perception* or propriocept* or propriocepsis or sensation* or ((body or kinesthetic or kinaesthetic or movement or position) N3 (discrimination or percept* or sense* or sensory or sensation*)) or ((balance or body or gait or posture or postural or musculoskeletal) N3 (abnormal* or adaption* or adjust* or control or difficult* or disequilibrium or disturbance* or entropy or equilibrium or error or impair* or instabilit* or oscillat* or orient* or react* or recover* or response* or restor* or stabili* or stance or steadiness or steady or sway* or unsteadiness or unsteady)) or (sensory N2 (function or functions)) ) OR AB ( (balance or "body sway" or "deep sensitivity" or imbalance or kinesthes* or perception* or propriocept* or propriocepsis or sensation* or ((body or kinesthetic or kinaesthetic or movement or position) N3 (discrimination or percept* or sense* or sensory or sensation*)) or ((balance or body or gait or posture or postural or musculoskeletal) N3 (abnormal* or adaption* or adjust* or control or difficult* or disequilibrium or disturbance* or entropy or equilibrium or error or impair* or instabilit* or oscillat* or orient* or react* or recover* or response* or restor* or stabili* or stance or steadiness or steady or sway* or unsteadiness or unsteady)) or (sensory N2 (function or functions)) ) | Expanders - Apply equivalent subjects  Search modes - Boolean/Phrase | Interface - EBSCOhost Research Databases  Search Screen - Advanced Search  Database - CINAHL Complete | Display |
| S7 | (MH "Balance, Postural") OR (MH "Proprioception") OR (MH "Kinesthesis") | Expanders - Apply equivalent subjects  Search modes - Boolean/Phrase | Interface - EBSCOhost Research Databases  Search Screen - Advanced Search  Database - CINAHL Complete | Display |
| S6 | (MM "Posture") OR TI (posture or postural) OR AB (posture OR postural) | Expanders - Apply equivalent subjects  Search modes - Boolean/Phrase | Interface - EBSCOhost Research Databases  Search Screen - Advanced Search  Database - CINAHL Complete | Display |
| S5 | TI ( dynamic N5 (balance or posture or postural) ) OR AB ( dynamic N5 (balance or posture or postural) ) | Expanders - Apply equivalent subjects  Search modes - Boolean/Phrase | Interface - EBSCOhost Research Databases  Search Screen - Advanced Search  Database - CINAHL Complete | Display |
| S4 | TI (reacti* or anticipatory or compensatory) OR AB (reacti* or anticipatory or compensatory) | Expanders - Apply equivalent subjects  Search modes - Boolean/Phrase | Interface - EBSCOhost Research Databases  Search Screen - Advanced Search  Database - CINAHL Complete | Display |
| S3 | (S1 OR S2) | Expanders - Apply equivalent subjects  Search modes - Boolean/Phrase | Interface - EBSCOhost Research Databases  Search Screen - Advanced Search  Database - CINAHL Complete | Display |
| S2 | TI ( concuss* or commotio or "dementia pugulistica" or mtbi or mtbis or post-concuss* or postconcuss* or sub-concuss* or subconcuss* or tbi or tbis or ((brain or cerebell* or cerebral or cerebrovascular or cortical or encephalopath* or head) N3 (commotio* or concuss* or contusion* or impact* or injur* or postconcuss* or post-concuss* or posttraumatic or post-traumatic or trauma*))) ) OR AB ( concuss* or commotio or "dementia pugulistica" or mtbi or mtbis or post-concuss* or postconcuss* or sub-concuss* or subconcuss* or tbi or tbis or ((brain or cerebell* or cerebral or cerebrovascular or cortical or encephalopath* or head) N3 (commotio* or concuss* or contusion* or impact* or injur* or postconcuss* or post-concuss* or posttraumatic or post-traumatic or trauma*))) ) | Expanders - Apply equivalent subjects  Search modes - Boolean/Phrase | Interface - EBSCOhost Research Databases  Search Screen - Advanced Search  Database - CINAHL Complete | Display |
| S1 | (MH "Brain Concussion") OR (MH "Postconcussion Syndrome") OR (MH "Brain Injuries") OR OR (MH "Chronic Traumatic Encephalopathy") OR (MH "Brain Contusions") | Expanders - Apply equivalent subjects  Search modes - Boolean/Phrase | Interface - EBSCOhost Research Databases  Search Screen - Advanced Search  Database - CINAHL Complete | Display |

------

Re-ran search on March 9, 2021

Limiters - Published Date: 20200101-20211231	156

#### **PsycINFO (Ebscohost) 2020-02-01**

| **#** | **Query** | **Limiters/Expanders** | **Last Run Via** | **Results** |
| --- | --- | --- | --- | --- |
| S33 | S31 NOT S32 | Expanders - Apply equivalent subjects  Search modes - Boolean/Phrase | Interface - EBSCOhost Research Databases  Search Screen - Advanced Search  Database - APA PsycInfo | 837 |
| S32 | DE "Animals" OR DE "Species Differences" OR DE "Animal Limb" OR DE "Animal Offspring" OR DE "Female Animals" OR DE "Infants (Animal)" OR DE "Invertebrates" OR DE "Male Animals" OR DE "Pets" OR DE "Service Animals" OR DE "Vertebrates" | Expanders - Apply equivalent subjects  Search modes - Boolean/Phrase | Interface - EBSCOhost Research Databases  Search Screen - Advanced Search  Database - APA PsycInfo | Display |
| S31 | S25 OR S26 OR S27 OR S28 OR S29 OR S30 | Expanders - Apply equivalent subjects  Search modes - Boolean/Phrase | Interface - EBSCOhost Research Databases  Search Screen - Advanced Search  Database - APA PsycInfo | Display |
| S30 | S3 AND S6 AND S9 | Expanders - Apply equivalent subjects  Search modes - Boolean/Phrase | Interface - EBSCOhost Research Databases  Search Screen - Advanced Search  Database - APA PsycInfo | Display |
| S29 | S3 AND S23 | Expanders - Apply equivalent subjects  Search modes - Boolean/Phrase | Interface - EBSCOhost Research Databases  Search Screen - Advanced Search  Database - APA PsycInfo | Display |
| S28 | S3 AND S19 AND S24 | Expanders - Apply equivalent subjects  Search modes - Boolean/Phrase | Interface - EBSCOhost Research Databases  Search Screen - Advanced Search  Database - APA PsycInfo | Display |
| S27 | S3 AND S4 AND S19 | Expanders - Apply equivalent subjects  Search modes - Boolean/Phrase | Interface - EBSCOhost Research Databases  Search Screen - Advanced Search  Database - APA PsycInfo | Display |
| S26 | S3 AND S5 | Expanders - Apply equivalent subjects  Search modes - Boolean/Phrase | Interface - EBSCOhost Research Databases  Search Screen - Advanced Search  Database - APA PsycInfo | Display |
| S25 | S3 AND S20 AND (TI (balance or posture or postural) or AB (balance or posture or postural)) | Expanders - Apply equivalent subjects  Search modes - Boolean/Phrase | Interface - EBSCOhost Research Databases  Search Screen - Advanced Search  Database - APA PsycInfo | Display |
| S24 | S20 OR S21 OR S22 OR S23 | Expanders - Apply equivalent subjects  Search modes - Boolean/Phrase | Interface - EBSCOhost Research Databases  Search Screen - Advanced Search  Database - APA PsycInfo | Display |
| S23 | TI ( ((balance or gait or perturbation* or postural or posture or propriocept* or sensorimotor* or slip or slipping or stance or stand or standing or step or stepping or trip or tripping or walk or walking) N5 (assess* or dual-task or "dual task" or simulat* or task or tasks or test or tests or testing)) ) OR AB ( ((balance or gait or perturbation* or postural or posture or propriocept* or sensorimotor* or slip or slipping or stance or stand or standing or step or stepping or trip or tripping or walk or walking) N5 (assess* or dual-task or "dual task" or simulat* or task or tasks or test or tests or testing)) ) | Expanders - Apply equivalent subjects  Search modes - Boolean/Phrase | Interface - EBSCOhost Research Databases  Search Screen - Advanced Search  Database - APA PsycInfo | Display |
| S22 | TI ( (("virtual reality" or vr) N3 (balance or propriocept* or posture or postural)) ) OR AB ( (("virtual reality" or vr) N3 (balance or propriocept* or posture or postural)) ) | Expanders - Apply equivalent subjects  Search modes - Boolean/Phrase | Interface - EBSCOhost Research Databases  Search Screen - Advanced Search  Database - APA PsycInfo | Display |
| S21 | TI ( (ActiveStep or BESTest or CAREN or "Computer Assisted Rehabilitation Environment" or "center of pressure" or "centre of pressure" or "central sensorimotor integration task" or GVS or "galvanic vestibular stimulation" or "heel contact latenc*" or "lean-and-release" or "lean and release" or "lean & release" or "limits of stability" or "mini BEST" or miniBEST or (motion N3 (track* or control* or test*)) or MCT or "motor control test" or neurocom or posturograph* or "Push-and-release" or "push and release" or "push & release" or P&R or "P and R" or "release-from-lean" or "release from lean" or Simbex or "SMART EquiTest" or "stance slip" or SVS or "vestibular stimulation" or "support surface" or (trunk N5 (angle or sway)) or VEMP or "vestibular evoked myogenic potential*" or vibrat* or "waist belt" or (waist N5 belt) or ((ground or forceplate or "force plate" or platform or surface) N5 (moving or tilt or translation*)) ) OR AB ( (ActiveStep or BESTest or CAREN or "Computer Assisted Rehabilitation Environment" or "center of pressure" or "centre of pressure" or "central sensorimotor integration task" or GVS or "galvanic vestibular stimulation" or "heel contact latenc*" or "lean-and-release" or "lean and release" or "lean & release" or "limits of stability" or "mini BEST" or miniBEST or (motion N3 (track* or control* or test*)) or MCT or "motor control test" or neurocom or posturograph* or "Push-and-release" or "push and release" or "push & release" or P&R or "P and R" or "release-from-lean" or "release from lean" or Simbex or "SMART EquiTest" or "stance slip" or SVS or "vestibular stimulation" or "support surface" or (trunk N5 (angle or sway)) or VEMP or "vestibular evoked myogenic potential*" or vibrat* or "waist belt" or (waist N5 belt) or ((ground or forceplate or "force plate" or platform or surface) N5 (moving or tilt or translation*)) ) | Expanders - Apply equivalent subjects  Search modes - Boolean/Phrase | Interface - EBSCOhost Research Databases  Search Screen - Advanced Search  Database - APA PsycInfo | Display |
| S20 | TI perturbation* OR AB perturbation* | Expanders - Apply equivalent subjects  Search modes - Boolean/Phrase | Interface - EBSCOhost Research Databases  Search Screen - Advanced Search  Database - APA PsycInfo | Display |
| S19 | S9 OR S12 OR S18 | Expanders - Apply equivalent subjects  Search modes - Boolean/Phrase | Interface - EBSCOhost Research Databases  Search Screen - Advanced Search  Database - APA PsycInfo | Display |
| S18 | S13 OR S14 OR S15 OR S16 OR S17 | Expanders - Apply equivalent subjects  Search modes - Boolean/Phrase | Interface - EBSCOhost Research Databases  Search Screen - Advanced Search  Database - APA PsycInfo | Display |
| S17 | DE "Labyrinth Disorders" OR DE "Vertigo" OR TI (Neurolabyrinthiti* or "Scarpa Ganglion*" or "Scarpas Ganglion*" or "Scarpa's Ganglion*" or vestibul* or vertigo*) OR AB (Neurolabyrinthiti* or "Scarpa Ganglion*" or "Scarpas Ganglion*" or "Scarpa's Ganglion*" or vestibul* or vertigo*) | Expanders - Apply equivalent subjects  Search modes - Boolean/Phrase | Interface - EBSCOhost Research Databases  Search Screen - Advanced Search  Database - APA PsycInfo | Display |
| S16 | DE "Sensory Gating" OR DE "Prepulse Inhibition" OR TI (((sensory or sensorimotor or startle) N3 (gating or filtering)) or (prepulse N2 (facilitate* or inhibit*))) OR AB (((sensory or sensorimotor or startle) N3 (gating or filtering)) or (prepulse N2 (facilitate* or inhibit*))) | Expanders - Apply equivalent subjects  Search modes - Boolean/Phrase | Interface - EBSCOhost Research Databases  Search Screen - Advanced Search  Database - APA PsycInfo | Display |
| S15 | DE "Sensory Feedback" OR TI (((acoustic* or audio* or haptic or propriocept* or sensorimotor or sensory or tactile or visual) N3 (feedback or feedbacks))) OR AB (((acoustic* or audio* or haptic or propriocept* or sensorimotor or sensory or tactile or visual) N3 (feedback or feedbacks))) | Expanders - Apply equivalent subjects  Search modes - Boolean/Phrase | Interface - EBSCOhost Research Databases  Search Screen - Advanced Search  Database - APA PsycInfo | Display |
| S14 | TI ( (("anterior parietal" or Brodmann or extrastriat* or motor or motorsensory or premotor or "post central" or postcentral* or precentral* or "pre central" or Rolandic or sensory or sensorimotor or "sensory motor" or "sensory-motor" or si or sensomotor or somatomotor or somatosensory or "somatic sensory" or striate or visual) N3 (area or areas or cortex or cortices or gyrus or region or strip or strips or zone)) ) OR AB ( (("anterior parietal" or Brodmann or extrastriat* or motor or motorsensory or premotor or "post central" or postcentral* or precentral* or "pre central" or Rolandic or sensory or sensorimotor or "sensory motor" or "sensory-motor" or si or sensomotor or somatomotor or somatosensory or "somatic sensory" or striate or visual) N3 (area or areas or cortex or cortices or gyrus or region or strip or strips or zone)) ) | Expanders - Apply equivalent subjects  Search modes - Boolean/Phrase | Interface - EBSCOhost Research Databases  Search Screen - Advanced Search  Database - APA PsycInfo | Display |
| S13 | DE "Visual Cortex" OR DE "Somatosensory Cortex" OR DE "Motor Cortex" | Expanders - Apply equivalent subjects  Search modes - Boolean/Phrase | Interface - EBSCOhost Research Databases  Search Screen - Advanced Search  Database - APA PsycInfo | Display |
| S12 | S10 OR S11 | Expanders - Apply equivalent subjects  Search modes - Boolean/Phrase | Interface - EBSCOhost Research Databases  Search Screen - Advanced Search  Database - APA PsycInfo | Display |
| S11 | TI ( ("auris interna" or "acoustic macula*" or "basilar papilla" or cochlea* or endolymph* or "fenestra rotunda*" or "internal acoustic pore" or "internal auditory meatus" or labyrinth* or "meatus acusticus" or "macula* acoustic" or otoconia* or otolith* or "papilla acustica basilaris" or "papilla basilar" or perilymph* or "porus acusticus internus" or saccule* or sacculus or "stria vascularis" or statoconi* or statocyst or statocyte or statolith or utricle* or utriculus or "vascularis stria" or vestibul* or ((ear or ears) N2 (helix or inner or internal or vestibul*)) or ((cochlea* or endolymphatic or perilymphatic or semicircular) N2 duct*) or (scala N2 (media* or tympani or tympanus or vestibuli or vestibulus)) or ((basilar or otolithic or statoconia or tectorial) N2 (lamina or membrane*)) or ((acoustic or auditory or corti* or spiral*) N2 (gangli* or lamina or laminas or ligament or ligaments or organ or organs or organum)) or ((ampullar* or auditory or cochlea* or inner or outer or vestibular) N2 (hair cell" or "hair cells")) or ((auditory or cochlea* or ear or semicircular or vestibul*) N2 (apparatus or aqueduct* or auris or canal or canals or canaliculus or "oval window*" or "round window*")) or (ampullar* N2 (crest or crests or crista))) ) OR AB ( ("auris interna" or "acoustic macula*" or "basilar papilla" or cochlea* or endolymph* or "fenestra rotunda*" or "internal acoustic pore" or "internal auditory meatus" or labyrinth* or "meatus acusticus" or "macula* acoustic" or otoconia* or otolith* or "papilla acustica basilaris" or "papilla basilar" or perilymph* or "porus acusticus internus" or saccule* or sacculus or "stria vascularis" or statoconi* or statocyst or statocyte or statolith or utricle* or utriculus or "vascularis stria" or vestibul* or ((ear or ears) N2 (helix or inner or internal or vestibul*)) or ((cochlea* or endolymphatic or perilymphatic or semicircular) N2 duct*) or (scala N2 (media* or tympani or tympanus or vestibuli or vestibulus)) or ((basilar or otolithic or statoconia or tectorial) N2 (lamina or membrane*)) or ((acoustic or auditory or corti* or spiral*) N2 (gangli* or lamina or laminas or ligament or ligaments or organ or organs or organum)) or ((ampullar* or auditory or cochlea* or inner or outer or vestibular) N2 (hair cell" or "hair cells")) or ((auditory or cochlea* or ear or semicircular or vestibul*) N2 (apparatus or aqueduct* or auris or canal or canals or canaliculus or "oval window*" or "round window*")) or (ampullar* N2 (crest or crests or crista))) ) | Expanders - Apply equivalent subjects  Search modes - Boolean/Phrase | Interface - EBSCOhost Research Databases  Search Screen - Advanced Search  Database - APA PsycInfo | Display |
| S10 | DE "Cochlea" OR DE "Labyrinth (Anatomy)" | Expanders - Apply equivalent subjects  Search modes - Boolean/Phrase | Interface - EBSCOhost Research Databases  Search Screen - Advanced Search  Database - APA PsycInfo | Display |
| S9 | S7 OR S8 | Expanders - Apply equivalent subjects  Search modes - Boolean/Phrase | Interface - EBSCOhost Research Databases  Search Screen - Advanced Search  Database - APA PsycInfo | Display |
| S8 | TI ( (balance or "body sway" or "deep sensitivity" or imbalance or kinesthes* or perception* or propriocept* or propriocepsis or sensation* or ((body or kinesthetic or kinaesthetic or movement or position) N3 (discrimination or percept* or sense* or sensory or sensation*)) or ((balance or body or gait or posture or postural or musculoskeletal) N3 (abnormal* or adaption* or adjust* or control or difficult* or disequilibrium or disturbance* or entropy or equilibrium or error or impair* or instabilit* or oscillat* or orient* or react* or recover* or response* or restor* or stabili* or stance or steadiness or steady or sway* or unsteadiness or unsteady)) or (sensory N2 (function or functions))) ) OR AB ( (balance or "body sway" or "deep sensitivity" or imbalance or kinesthes* or perception* or propriocept* or propriocepsis or sensation* or ((body or kinesthetic or kinaesthetic or movement or position) N3 (discrimination or percept* or sense* or sensory or sensation*)) or ((balance or body or gait or posture or postural or musculoskeletal) N3 (abnormal* or adaption* or adjust* or control or difficult* or disequilibrium or disturbance* or entropy or equilibrium or error or impair* or instabilit* or oscillat* or orient* or react* or recover* or response* or restor* or stabili* or stance or steadiness or steady or sway* or unsteadiness or unsteady)) or (sensory N2 (function or functions))) ) | Expanders - Apply equivalent subjects  Search modes - Boolean/Phrase | Interface - EBSCOhost Research Databases  Search Screen - Advanced Search  Database - APA PsycInfo | Display |
| S7 | DE "Postural Sway" OR DE "Proprioception" OR DE "Kinesthetic Perception" OR DE "Perception" | Expanders - Apply equivalent subjects  Search modes - Boolean/Phrase | Interface - EBSCOhost Research Databases  Search Screen - Advanced Search  Database - APA PsycInfo | Display |
| S6 | DE "Posture" OR TI (posture or postural) OR AB (posture or postural) | Expanders - Apply equivalent subjects  Search modes - Boolean/Phrase | Interface - EBSCOhost Research Databases  Search Screen - Advanced Search  Database - APA PsycInfo | Display |
| S5 | TI ( (dynamic N5 (balance or posture or postural)) ) OR AB ( (dynamic N5 (balance or posture or postural)) ) | Expanders - Apply equivalent subjects  Search modes - Boolean/Phrase | Interface - EBSCOhost Research Databases  Search Screen - Advanced Search  Database - APA PsycInfo | Display |
| S4 | TI (reacti* or anticipatory or compensatory) OR AB (reacti* or anticipatory or compensatory) | Expanders - Apply equivalent subjects  Search modes - Boolean/Phrase | Interface - EBSCOhost Research Databases  Search Screen - Advanced Search  Database - APA PsycInfo | Display |
| S3 | S1 OR S2 | Expanders - Apply equivalent subjects  Search modes - Boolean/Phrase | Interface - EBSCOhost Research Databases  Search Screen - Advanced Search  Database - APA PsycInfo | Display |
| S2 | TI ( concuss* or commotio or "dementia pugulistica" or mtbi or mtbis or post-concuss* or postconcuss* or sub-concuss* or subconcuss* or tbi or tbis or ((brain or cerebell* or cerebral or cerebrovascular or cortical or encephalopath* or head) N3 (commotio* or concuss* or contusion* or impact* or injur* or postconcuss* or post-concuss* or posttraumatic or post-traumatic or trauma*))) ) OR AB ( concuss* or commotio or "dementia pugulistica" or mtbi or mtbis or post-concuss* or postconcuss* or sub-concuss* or subconcuss* or tbi or tbis or ((brain or cerebell* or cerebral or cerebrovascular or cortical or encephalopath* or head) N3 (commotio* or concuss* or contusion* or impact* or injur* or postconcuss* or post-concuss* or posttraumatic or post-traumatic or trauma*))) ) | Expanders - Apply equivalent subjects  Search modes - Boolean/Phrase | Interface - EBSCOhost Research Databases  Search Screen - Advanced Search  Database - APA PsycInfo | Display |
| S1 | DE "Brain Concussion" OR DE "Traumatic Brain Injury" | Expanders - Apply equivalent subjects  Search modes - Boolean/Phrase | Interface - EBSCOhost Research Databases  Search Screen - Advanced Search  Database - APA PsycInfo | Display |

------

Re-ran search on March 9, 2021

Limiters - Published Date: 20200101-20211231	85

#### **SportDiscus (Ebscohost) 2020-02-01**

| **#** | **Query** | **Limiters/Expanders** | **Last Run Via** | **Results** |
| --- | --- | --- | --- | --- |
| S30 | S28 NOT S29 | Expanders - Apply related words; Apply equivalent subjects  Search modes - Boolean/Phrase | Interface - EBSCOhost Research Databases  Search Screen - Advanced Search  Database - SPORTDiscus with Full Text | 533 |
| S29 | TI ( animal or animals or beaver or beavers or beef or bovine or breeding or bull or canine or castoris or cat or cattle or cats or chicken or chickens or chimp* or cow or dog or dogs or equine or foal or foals or fish or insect or insects or horse or horses or livestock or mice or monkey or monkeys or mouse or murine or plant or plants or pork or porcine or protozoa or protozoas or purebred or rabbit or rabbits or rat or rats or rodent or rodents or sheep or thoroughbred or veterinar* ) OR AB veterinar* | Expanders - Apply related words; Apply equivalent subjects  Search modes - Boolean/Phrase | Interface - EBSCOhost Research Databases  Search Screen - Advanced Search  Database - SPORTDiscus with Full Text | 43,070 |
| S28 | S22 OR S23 OR S24 OR S25 OR S26 OR S27 | Expanders - Apply related words; Apply equivalent subjects  Search modes - Boolean/Phrase | Interface - EBSCOhost Research Databases  Search Screen - Advanced Search  Database - SPORTDiscus with Full Text | 537 |
| S27 | S3 AND S6 AND S9 | Expanders - Apply related words; Apply equivalent subjects  Search modes - Boolean/Phrase | Interface - EBSCOhost Research Databases  Search Screen - Advanced Search  Database - SPORTDiscus with Full Text | 205 |
| S26 | S3 AND S20 | Expanders - Apply related words; Apply equivalent subjects  Search modes - Boolean/Phrase | Interface - EBSCOhost Research Databases  Search Screen - Advanced Search  Database - SPORTDiscus with Full Text | 383 |
| S25 | S3 AND S16 AND S21 | Expanders - Apply related words; Apply equivalent subjects  Search modes - Boolean/Phrase | Interface - EBSCOhost Research Databases  Search Screen - Advanced Search  Database - SPORTDiscus with Full Text | 371 |
| S24 | S3 AND S4 AND S16 | Expanders - Apply related words; Apply equivalent subjects  Search modes - Boolean/Phrase | Interface - EBSCOhost Research Databases  Search Screen - Advanced Search  Database - SPORTDiscus with Full Text | 76 |
| S23 | S3 AND S5 | Expanders - Apply related words; Apply equivalent subjects  Search modes - Boolean/Phrase | Interface - EBSCOhost Research Databases  Search Screen - Advanced Search  Database - SPORTDiscus with Full Text | 43 |
| S22 | S3 AND S17 AND (TI (balance or posture or postural) OR AB (balance or posture or postural)) | Expanders - Apply related words; Apply equivalent subjects  Search modes - Boolean/Phrase | Interface - EBSCOhost Research Databases  Search Screen - Advanced Search  Database - SPORTDiscus with Full Text | 7 |
| S21 | S17 OR S18 OR S19 OR S20 | Expanders - Apply related words; Apply equivalent subjects  Search modes - Boolean/Phrase | Interface - EBSCOhost Research Databases  Search Screen - Advanced Search  Database - SPORTDiscus with Full Text | 48,943 |
| S20 | TI ( ((balance or gait or perturbation* or postural or posture or propriocept* or sensorimotor* or slip or slipping or stance or stand or standing or step or stepping or trip or tripping or walk or walking) N5 (assess* or dual-task or "dual task" or simulat* or task or tasks or test or tests or testing)) ) OR AB ( ((balance or gait or perturbation* or postural or posture or propriocept* or sensorimotor* or slip or slipping or stance or stand or standing or step or stepping or trip or tripping or walk or walking) N5 (assess* or dual-task or "dual task" or simulat* or task or tasks or test or tests or testing)) ) | Expanders - Apply related words; Apply equivalent subjects  Search modes - Boolean/Phrase | Interface - EBSCOhost Research Databases  Search Screen - Advanced Search  Database - SPORTDiscus with Full Text | 14,565 |
| S19 | TI ( (("virtual reality" or vr) N3 (balance or propriocept* or posture or postural)) ) OR AB ( (("virtual reality" or vr) N3 (balance or propriocept* or posture or postural)) ) | Expanders - Apply related words; Apply equivalent subjects  Search modes - Boolean/Phrase | Interface - EBSCOhost Research Databases  Search Screen - Advanced Search  Database - SPORTDiscus with Full Text | 45 |
| S18 | TI ( ActiveStep or BESTest or CAREN or "Computer Assisted Rehabilitation Environment" or "center of pressure" or "centre of pressure" or "central sensorimotor integration task" or GVS or "galvanic vestibular stimulation" or "heel contact latenc*" or "lean-and-release" or "lean and release" or "lean & release" or "limits of stability" or "mini BEST" or miniBEST or (motion N3 (track* or control* or test*)) or MCT or "motor control test" or neurocom or posturograph* or "Push-and-release" or "push and release" or "push & release" or P&R or "P and R" or "release-from-lean" or "release from lean" or Simbex or "SMART EquiTest" or "stance slip" or SVS or "vestibular stimulation" or "support surface" or (trunk N5 (angle or sway)) or VEMP or "vestibular evoked myogenic potential*" or vibrat* or "waist belt" or (waist N5 belt) or ((ground or forceplate or "force plate" or platform or surface) N5 (moving or tilt or translation*)) ) OR AB ( ActiveStep or BESTest or CAREN or "Computer Assisted Rehabilitation Environment" or "center of pressure" or "centre of pressure" or "central sensorimotor integration task" or GVS or "galvanic vestibular stimulation" or "heel contact latenc*" or "lean-and-release" or "lean and release" or "lean & release" or "limits of stability" or "mini BEST" or miniBEST or (motion N3 (track* or control* or test*)) or MCT or "motor control test" or neurocom or posturograph* or "Push-and-release" or "push and release" or "push & release" or P&R or "P and R" or "release-from-lean" or "release from lean" or Simbex or "SMART EquiTest" or "stance slip" or SVS or "vestibular stimulation" or "support surface" or (trunk N5 (angle or sway)) or VEMP or "vestibular evoked myogenic potential*" or vibrat* or "waist belt" or (waist N5 belt) or ((ground or forceplate or "force plate" or platform or surface) N5 (moving or tilt or translation*)) ) | Expanders - Apply related words; Apply equivalent subjects  Search modes - Boolean/Phrase | Interface - EBSCOhost Research Databases  Search Screen - Advanced Search  Database - SPORTDiscus with Full Text | 35,505 |
| S17 | TI perturbation* OR AB perturbation* | Expanders - Apply related words; Apply equivalent subjects  Search modes - Boolean/Phrase | Interface - EBSCOhost Research Databases  Search Screen - Advanced Search  Database - SPORTDiscus with Full Text | 2,222 |
| S16 | (S9 OR S10 OR S11 OR S12 OR S13 OR S14 OR S15) | Expanders - Apply related words; Apply equivalent subjects  Search modes - Boolean/Phrase | Interface - EBSCOhost Research Databases  Search Screen - Advanced Search  Database - SPORTDiscus with Full Text | 79,944 |
| S15 | DE "VERTIGO" OR TI ( (Neurolabyrinthiti* or "Scarpa Ganglion*" or "Scarpas Ganglion*" or "Scarpa's Ganglion*" or vestibul* or vertigo*) ) OR AB ( (Neurolabyrinthiti* or "Scarpa Ganglion*" or "Scarpas Ganglion*" or "Scarpa's Ganglion*" or vestibul* or vertigo*) ) | Expanders - Apply related words; Apply equivalent subjects  Search modes - Boolean/Phrase | Interface - EBSCOhost Research Databases  Search Screen - Advanced Search  Database - SPORTDiscus with Full Text | 1,464 |
| S14 | TI ( ((sensory or sensorimotor or startle) N3 (gating or filtering)) or ((prepulse or pre-pulse) N2 facilitate* or inhibit*)) ) OR AB ( ((sensory or sensorimotor or startle) N3 (gating or filtering)) or ((prepulse or pre-pulse) N2 (facilitate* or inhibit*)) ) | Expanders - Apply related words; Apply equivalent subjects  Search modes - Boolean/Phrase | Interface - EBSCOhost Research Databases  Search Screen - Advanced Search  Database - SPORTDiscus with Full Text | 4,293 |
| S13 | TI ( (acoustic* or audio* or haptic or propriocept* or sensorimotor or sensory or tactile or visual) N3 (feedback or feedbacks)) ) OR AB ( (acoustic* or audio* or haptic or propriocept* or sensorimotor or sensory or tactile or visual) N3 (feedback or feedbacks)) ) | Expanders - Apply related words; Apply equivalent subjects  Search modes - Boolean/Phrase | Interface - EBSCOhost Research Databases  Search Screen - Advanced Search  Database - SPORTDiscus with Full Text | 1,159 |
| S12 | TI ( (("anterior parietal" or Brodmann or extrastriat* or motor or motorsensory or premotor or "post central" or postcentral* or precentral* or "pre central" or Rolandic or sensory or sensorimotor or "sensory motor" or "sensory-motor" or si or sensomotor or somatomotor or somatosensory or "somatic sensory" or striate or visual) N3 (area or areas or cortex or cortices or gyrus or region or strip or strips or zone)) ) OR AB ( (("anterior parietal" or Brodmann or extrastriat* or motor or motorsensory or premotor or "post central" or postcentral* or precentral* or "pre central" or Rolandic or sensory or sensorimotor or "sensory motor" or "sensory-motor" or si or sensomotor or somatomotor or somatosensory or "somatic sensory" or striate or visual) N3 (area or areas or cortex or cortices or gyrus or region or strip or strips or zone)) ) | Expanders - Apply related words; Apply equivalent subjects  Search modes - Boolean/Phrase | Interface - EBSCOhost Research Databases  Search Screen - Advanced Search  Database - SPORTDiscus with Full Text | 1,424 |
| S11 | TI ( ("auris interna" or "acoustic macula*" or "basilar papilla" or cochlea* or endolymph* or "fenestra rotunda*" or "internal acoustic pore" or "internal auditory meatus" or labyrinth* or "meatus acusticus" or "macula* acoustic" or otoconia* or otolith* or "papilla acustica basilaris" or "papilla basilar" or perilymph* or "porus acusticus internus" or saccule* or sacculus or "stria vascularis" or statoconi* or statocyst or statocyte or statolith or utricle* or utriculus or "vascularis stria" or vestibul* or ((ear or ears) N2 (helix or inner or internal or vestibul*)) or ((cochlea* or endolymphatic or perilymphatic or semicircular) N2 duct*) or (scala N2 (media* or tympani or tympanus or vestibuli or vestibulus)) or ((basilar or otolithic or statoconia or tectorial) N2 (lamina or membrane*)) or ((acoustic or auditory or corti* or spiral*) N2 (gangli* or lamina or laminas or ligament or ligaments or organ or organs or organum)) or ((ampullar* or auditory or cochlea* or inner or outer or vestibular) N2 ("hair cell" or "hair cells")) or ((auditory or cochlea* or ear or semicircular or vestibul*) N2 (apparatus or aqueduct* or auris or canal or canals or canaliculus or "oval window*" or "round window*")) or (ampullar* N2 (crest or crests or crista))) ) OR AB ( ("auris interna" or "acoustic macula*" or "basilar papilla" or cochlea* or endolymph* or "fenestra rotunda*" or "internal acoustic pore" or "internal auditory meatus" or labyrinth* or "meatus acusticus" or "macula* acoustic" or otoconia* or otolith* or "papilla acustica basilaris" or "papilla basilar" or perilymph* or "porus acusticus internus" or saccule* or sacculus or "stria vascularis" or statoconi* or statocyst or statocyte or statolith or utricle* or utriculus or "vascularis stria" or vestibul* or ((ear or ears) N2 (helix or inner or internal or vestibul*)) or ((cochlea* or endolymphatic or perilymphatic or semicircular) N2 duct*) or (scala N2 (media* or tympani or tympanus or vestibuli or vestibulus)) or ((basilar or otolithic or statoconia or tectorial) N2 (lamina or membrane*)) or ((acoustic or auditory or corti* or spiral*) N2 (gangli* or lamina or laminas or ligament or ligaments or organ or organs or organum)) or ((ampullar* or auditory or cochlea* or inner or outer or vestibular) N2 ("hair cell" or "hair cells")) or ((auditory or cochlea* or ear or semicircular or vestibul*) N2 (apparatus or aqueduct* or auris or canal or canals or canaliculus or "oval window*" or "round window*")) or (ampullar* N2 (crest or crests or crista))) ) | Expanders - Apply related words; Apply equivalent subjects  Search modes - Boolean/Phrase | Interface - EBSCOhost Research Databases  Search Screen - Advanced Search  Database - SPORTDiscus with Full Text | 1,519 |
| S10 | DE "LABYRINTH (Ear)" OR DE "COCHLEA" OR DE "VESTIBULAR apparatus" OR DE "UTRICLE & Saccule" | Expanders - Apply related words; Apply equivalent subjects  Search modes - Boolean/Phrase | Interface - EBSCOhost Research Databases  Search Screen - Advanced Search  Database - SPORTDiscus with Full Text | 216 |
| S9 | S7 OR S8 | Expanders - Apply related words; Apply equivalent subjects  Search modes - Boolean/Phrase | Interface - EBSCOhost Research Databases  Search Screen - Advanced Search  Database - SPORTDiscus with Full Text | 72,788 |
| S8 | TI ( (balance or "body sway" or "deep sensitivity" or imbalance or kinesthes* or perception* or propriocept* or propriocepsis or sensation* or ((body or kinesthetic or kinaesthetic or movement or position) N3 (discrimination or percept* or sense* or sensory or sensation*)) or ((balance or body or gait or posture or postural or musculoskeletal) N3 (abnormal* or adaption* or adjust* or control or difficult* or disequilibrium or disturbance* or entropy or equilibrium or error or impair* or instabilit* or oscillat* or orient* or react* or recover* or response* or restor* or stabili* or stance or steadiness or steady or sway* or unsteadiness or unsteady)) or (sensory N2 (function or functions))) ) OR AB ( (balance or "body sway" or "deep sensitivity" or imbalance or kinesthes* or perception* or propriocept* or propriocepsis or sensation* or ((body or kinesthetic or kinaesthetic or movement or position) N3 (discrimination or percept* or sense* or sensory or sensation*)) or ((balance or body or gait or posture or postural or musculoskeletal) N3 (abnormal* or adaption* or adjust* or control or difficult* or disequilibrium or disturbance* or entropy or equilibrium or error or impair* or instabilit* or oscillat* or orient* or react* or recover* or response* or restor* or stabili* or stance or steadiness or steady or sway* or unsteadiness or unsteady)) or (sensory N2 (function or functions))) ) | Expanders - Apply related words; Apply equivalent subjects  Search modes - Boolean/Phrase | Interface - EBSCOhost Research Databases  Search Screen - Advanced Search  Database - SPORTDiscus with Full Text | 68,958 |
| S7 | DE "EQUILIBRIUM (Physiology)" OR DE "MUSCULAR sense" OR DE "PROPRIOCEPTION" OR DE "PERCEPTION" | Expanders - Apply related words; Apply equivalent subjects  Search modes - Boolean/Phrase | Interface - EBSCOhost Research Databases  Search Screen - Advanced Search  Database - SPORTDiscus with Full Text | 12,846 |
| S6 | DE "POSTURE" OR TI (posture or postural) OR AB (posture or postural) | Expanders - Apply related words; Apply equivalent subjects  Search modes - Boolean/Phrase | Interface - EBSCOhost Research Databases  Search Screen - Advanced Search  Database - SPORTDiscus with Full Text | 17,166 |
| S5 | TI ( (dynamic N5 (balance or posture or postural)) ) OR AB ( (dynamic N5 (balance or posture or postural)) ) | Expanders - Apply related words; Apply equivalent subjects  Search modes - Boolean/Phrase | Interface - EBSCOhost Research Databases  Search Screen - Advanced Search  Database - SPORTDiscus with Full Text | 2,156 |
| S4 | TI ( (reacti* or anticipatory or compensatory) ) OR AB ( (reacti* or anticipatory or compensatory) ) | Expanders - Apply related words; Apply equivalent subjects  Search modes - Boolean/Phrase | Interface - EBSCOhost Research Databases  Search Screen - Advanced Search  Database - SPORTDiscus with Full Text | 31,946 |
| S3 | S1 OR S2 | Expanders - Apply related words; Apply equivalent subjects  Search modes - Boolean/Phrase | Interface - EBSCOhost Research Databases  Search Screen - Advanced Search  Database - SPORTDiscus with Full Text | 12,017 |
| S2 | TI ( (concuss* or commotio or "dementia pugulistica" or mtbi or mtbis or post-concuss* or postconcuss* or sub-concuss* or subconcuss* or tbi or tbis or ((brain or cerebell* or cerebral or cerebrovascular or cortical or encephalopath* or head) N3 (commotio* or concuss* or contusion* or impact* or injur* or postconcuss* or post-concuss* or posttraumatic or post-traumatic or trauma*))) ) OR AB ( (concuss* or commotio or "dementia pugulistica" or mtbi or mtbis or post-concuss* or postconcuss* or sub-concuss* or subconcuss* or tbi or tbis or ((brain or cerebell* or cerebral or cerebrovascular or cortical or encephalopath* or head) N3 (commotio* or concuss* or contusion* or impact* or injur* or postconcuss* or post-concuss* or posttraumatic or post-traumatic or trauma*))) ) | Expanders - Apply related words; Apply equivalent subjects  Search modes - Boolean/Phrase | Interface - EBSCOhost Research Databases  Search Screen - Advanced Search  Database - SPORTDiscus with Full Text | 11,705 |
| S1 | DE "BRAIN concussion" OR DE "POSTCONCUSSION syndrome" OR DE "CHRONIC traumatic encephalopathy" OR DE "BRAIN injuries" | Expanders - Apply related words; Apply equivalent subjects  Search modes - Boolean/Phrase | Interface - EBSCOhost Research Databases  Search Screen - Basic Search  Database - SPORTDiscus with Full Text | 5,016 |

------

Re-ran search on March 9, 2021

Limiters - Published Date: 20200101-20211231	85

#### **Cochrane Library (Wiley.com) 2021-03-09**

ID	Search	Hits

#1	MeSH descriptor: [Brain Injuries, Traumatic] this term only	591

#2	MeSH descriptor: [Brain Concussion] this term only	298

#3	MeSH descriptor: [Brain Contusion] this term only	3

#4	MeSH descriptor: [Chronic Traumatic Encephalopathy] this term only	1

#5	MeSH descriptor: [Head Injuries, Closed] this term only	79

#6	((concuss* or commotio or "dementia pugulistica" or mtbi or mtbis or post-concuss* or postconcuss* or sub-concuss* or subconcuss* or tbi or tbis or ((brain or cerebell* or cerebral or cerebrovascular or cortical or encephalopath* or head) adj3 (commotio* or concuss* or contusion* or impact* or injur* or postconcuss* or post-concuss* or posttraumatic or post-traumatic or trauma*)))):ti AND ((concuss* or commotio or "dementia pugulistica" or mtbi or mtbis or post-concuss* or postconcuss* or sub-concuss* or subconcuss* or tbi or tbis or ((brain or cerebell* or cerebral or cerebrovascular or cortical or encephalopath* or head) adj3 (commotio* or concuss* or contusion* or impact* or injur* or postconcuss* or post-concuss* or posttraumatic or post-traumatic or trauma*)))):ab (Word variations have been searched)	738

#7	#1 OR #2 OR #3 OR #4 OR #5 OR #6	1382

#8	((reacti* or anticipatory or compensatory)):ti OR ((reacti* or anticipatory or compensatory)):ab (Word variations have been searched)	80493

#9	(dynamic NEAR/5 (balance or posture or postural)):ti,ab,kw	1692

#10	MeSH descriptor: [Posture] this term only	3381

#11	(posture or postural):ti,ab,kw	13011

#12	#10 OR #11	13011

#13	MeSH descriptor: [Postural Balance] this term only	2776

#14	MeSH descriptor: [Kinesthesis] this term only	149

#15	MeSH descriptor: [Proprioception] this term only	538

#16	MeSH descriptor: [Perception] this term only	1499

#17	MeSH descriptor: [Sensation] this term only	821

#18	(balance or "body sway" or "deep sensitivity" or imbalance or kinesthes* or perception* or propriocept* or propriocepsis or sensation* or ((body or kinesthetic or kinaesthetic or movement or position) NEAR/3 (discrimination or percept* or sense* or sensory or sensation*)) or ((balance or body or gait or posture or postural or musculoskeletal) NEAR/3 (abnormal* or adaption* or adjust* or control or difficult* or disequilibrium or disturbance* or entropy or equilibrium or error or impair* or instabilit* or oscillat* or orient* or react* or recover* or response* or restor* or stabili* or stance or steadiness or steady or sway* or unsteadiness or unsteady)) or (sensory NEAR/2 (function or functions))):ti,ab,kw	78142

#19	#13 OR #14 OR #15 OR #16 OR #17 OR #18	78142

#20	MeSH descriptor: [Ear, Inner] this term only	48

#21	MeSH descriptor: [Cochlea] this term only	67

#22	MeSH descriptor: [Basilar Membrane] this term only	3

#23	MeSH descriptor: [Cochlear Aqueduct] this term only	0

#24	MeSH descriptor: [Cochlear Duct] this term only	1

#25	MeSH descriptor: [Stria Vascularis] this term only	0

#26	MeSH descriptor: [Tectorial Membrane] this term only	0

#27	MeSH descriptor: [Organ of Corti] this term only	3

#28	MeSH descriptor: [Hair Cells, Auditory] this term only	1

#29	MeSH descriptor: [Hair Cells, Auditory, Inner] this term only	1

#30	MeSH descriptor: [Hair Cells, Auditory, Outer] this term only	5

#31	MeSH descriptor: [Labyrinth Supporting Cells] this term only	0

#32	MeSH descriptor: [Round Window, Ear] this term only	12

#33	MeSH descriptor: [Scala Tympani] this term only	2

#34	MeSH descriptor: [Scala Vestibuli] this term only	0

#35	MeSH descriptor: [Spiral Ganglion] this term only	0

#36	MeSH descriptor: [Spiral Lamina] this term only	0

#37	MeSH descriptor: [Spiral Ligament of Cochlea] this term only	0

#38	MeSH descriptor: [Labyrinthine Fluids] this term only	3

#39	MeSH descriptor: [Semicircular Canals] this term only	57

#40	MeSH descriptor: [Semicircular Ducts] this term only	0

#41	MeSH descriptor: [Hair Cells, Ampulla] this term only	0

#42	MeSH descriptor: [Vestibule, Labyrinth] this term only	160

#43	MeSH descriptor: [Oval Window, Ear] this term only	1

#44	MeSH descriptor: [Saccule and Utricle] this term only	13

#45	MeSH descriptor: [Acoustic Maculae] this term only	0

#46	MeSH descriptor: [Hair Cells, Vestibular] this term only	0

#47	MeSH descriptor: [Otolithic Membrane] this term only	26

#48	MeSH descriptor: [Vestibular Aqueduct] this term only	2

#49	MeSH descriptor: [Endolymphatic Duct] this term only	3

#50	MeSH descriptor: [Endolymphatic Sac] this term only	11

#51	("auris interna" or "acoustic macula*" or "basilar papilla" or cochlea* or endolymph* or "fenestra rotunda*" or "internal acoustic pore" or "internal auditory meatus" or labyrinth* or "meatus acusticus" or "macula* acoustic" or otoconia* or otolith* or "papilla acustica basilaris" or "papilla basilar" or perilymph* or "porus acusticus internus" or saccule* or sacculus or "stria vascularis" or statoconi* or statocyst or statocyte or statolith or utricle* or utriculus or "vascularis stria" or vestibul* or ((ear or ears) NEAR/2 (helix or inner or internal or vestibul*)) or ((cochlea* or endolymphatic or perilymphatic or semicircular) NEAR/2 duct*) or (scala NEAR/2 (media* or tympani or tympanus or vestibuli or vestibulus)) or ((basilar or otolithic or statoconia or tectorial) NEAR/2 (lamina or membrane*)) or ((acoustic or auditory or corti* or spiral*) NEAR/2 (gangli* or lamina or laminas or ligament or ligaments or organ or organs or organum)) or ((ampullar* or auditory or cochlea* or inner or outer or vestibular) NEAR/2 ("hair cell" or "hair cells")) or ((auditory or cochlea* or ear or semicircular or vestibul*) NEAR/2 (apparatus or aqueduct* or auris or canal or canals or canaliculus or "oval window*" or "round window*")) or (ampullar* NEAR/2 (crest or crests or crista))):ti,ab,kw	4313

#52	MeSH descriptor: [Sensorimotor Cortex] this term only	31

#53	MeSH descriptor: [Motor Cortex] this term only	793

#54	MeSH descriptor: [Somatosensory Cortex] this term only	159

#55	MeSH descriptor: [Visual Cortex] this term only	173

#56	(("anterior parietal" or Brodmann or extrastriat* or motor or motorsensory or premotor or "post central" or postcentral* or precentral* or "pre central" or Rolandic or sensory or sensorimotor or "sensory motor" or "sensory-motor" or si or sensomotor or somatomotor or somatosensory or "somatic sensory" or striate or visual) NEAR/3 (area or areas or cortex or cortices or gyrus or region or strip or strips or zone)):ti,ab,kw	5752

#57	MeSH descriptor: [Feedback, Sensory] this term only	243

#58	((acoustic* or audio* or haptic or propriocept* or sensorimotor or sensory or tactile or visual) NEAR/3 (feedback or feedbacks)):ti,ab,kw	1554

#59	MeSH descriptor: [Sensory Gating] this term only	46

#60	MeSH descriptor: [Prepulse Inhibition] this term only	4

#61	((sensory or sensorimotor or startle) NEAR/3 (gating or filtering)):ti,ab,kw or ((prepulse or pre-pulse) NEAR/2 (facilitate* or inhibit*)):ti,ab,kw	268

#62	MeSH descriptor: [Vestibular Diseases] this term only	168

#63	MeSH descriptor: [Bilateral Vestibulopathy] this term only	1

#64	MeSH descriptor: [Vertigo] this term only	466

#65	MeSH descriptor: [Benign Paroxysmal Positional Vertigo] this term only	86

#66	MeSH descriptor: [Vestibular Nerve] this term only	21

#67	MeSH descriptor: [Vestibular Neuronitis] this term only	27

#68	(Neurolabyrinthiti* or "Scarpa Ganglion*" or "Scarpas Ganglion*" or "Scarpa's Ganglion*" or vestibul* or vertigo*):ti,ab,kw	6736

#69	#19 or #20 OR #21 OR #22 OR #23 OR #24 OR #25 OR #26 OR #27 OR #28 OR #29 OR #30 OR #31 OR #32 OR #33 OR #34 OR #35 OR #36 OR #37 OR #38 OR #39 OR #40 OR #41 OR #42 OR #43 OR #44 OR #45 OR #46 OR #47 OR #48 OR #49 OR #50 OR #51 OR #52 OR #53 OR #54 OR #55 OR #56 OR #57 OR #58 OR #59 OR #60 OR #61 OR #62 OR #63 OR #64 OR #65 OR #66 OR #67 OR #68	90559

#70	perturbation*:ti,ab,kw	1302

#71	(ActiveStep or BESTest or CAREN or "Computer Assisted Rehabilitation Environment" or "center of pressure" or "centre of pressure" or "central sensorimotor integration task" or GVS or "galvanic vestibular stimulation" or "heel contact latenc*" or "lean-and-release" or "lean and release" or "lean & release" or "limits of stability" or "mini BEST" or miniBEST or (motion NEAR/3 (track* or control* or test*)) or MCT or "motor control test" or neurocom or posturograph* or "Push-and-release" or "push and release" or "push & release" or P&R or "P and R" or "release-from-lean" or "release from lean" or Simbex or "SMART EquiTest" or "stance slip" or SVS or "vestibular stimulation" or "support surface" or (trunk NEAR/5 (angle or sway)) or VEMP or "vestibular evoked myogenic potential*" or vibrat* or "waist belt" or (waist NEAR/5 belt)):ti,ab,kw or ((ground or forceplate or "force plate" or platform or surface) NEAR/5 (moving or tilt or translation*)):ti,ab,kw	8587

#72	(("virtual reality" or vr) NEAR/3 (balance or propriocept* or posture or postural)):ti,ab,kw	153

#73	((balance or gait or perturbation* or postural or posture or propriocept* or sensorimotor* or slip or slipping or stance or stand or standing or step or stepping or trip or tripping or walk or walking) NEAR/5 (assess* or dual-task or "dual task" or simulat* or task or tasks or test or tests or testing)):ti,ab,kw	20313

#74	#70 OR #71 OR #72 OR #73	28433

#75	(balance or posture or postural):ti,ab,kw	33979

#76	#7 AND #70 AND #75	0

#77	#7 AND #9	11

#78	#7 AND #8 AND #69	13

#79	#7 AND #69 AND #74	61

#80	#7 AND #73	59

#81	#7 AND #12 AND #19	33

#82	#76 OR #77 OR #78 OR #79 OR #80 OR #81	85

#83	(animal or animals or beaver or beavers or beef or bovine or breeding or bull or canine or castoris or cat or cattle or cats or chicken or chickens or chimp* or cow or dog or dogs or equine or foal or foals or fish or insect or insects or horse or horses or livestock or mice or monkey or monkeys or mouse or murine or plant or plants or pork or porcine or protozoa? or purebred or rabbit or rabbits or rat or rats or rodent or rodents or sheep or thoroughbred):ti or (veterinar*):ti,ab	10107

#84	#82 NOT #83	85

#### **Web of Science (Clarivate Analytics) 2020-02-03**

| # 26 | [1,845](http://apps.webofknowledge.com/summary.do;jsessionid=F8E63CDD2A44FEADE42E163384D2A58D?product=WOS&doc=1&qid=37&SID=8EMF6vM4VEGGXsb3ob8&search_mode=AdvancedSearch&update_back2search_link_param=yes) | #24 NOT #25  Indexes=SCI-EXPANDED, SSCI, A&HCI, ESCI Timespan=1900-2020 | [Edit](http://apps.webofknowledge.com/WOS_AdvancedSearch_input.do?product=WOS&SID=8EMF6vM4VEGGXsb3ob8&search_mode=AdvancedSearch&replaceSetId=26&editState=init) |
| --- | --- | --- | --- |
| # 25 | [3,599,295](http://apps.webofknowledge.com/summary.do;jsessionid=F8E63CDD2A44FEADE42E163384D2A58D?product=WOS&doc=1&qid=34&SID=8EMF6vM4VEGGXsb3ob8&search_mode=GeneralSearch&update_back2search_link_param=yes) | **TITLE:** (animal or animals or beaver or beavers or beef or bovine or breeding or bull or canine or castoris or cat or cattle or cats or chicken or chickens or chimp* or cow or dog or dogs or equine or foal or foals or fish or insect? or horse or horses or livestock or mice or monkey or monkeys or mouse or murine or plant or plants or pork or porcine or protozoa or protozoas or purebred or rabbit or rabbits or rat or rats or rodent or rodents or sheep or thoroughbred or veterinar*)  Indexes=SCI-EXPANDED, SSCI, A&HCI, ESCI Timespan=1900-2020 | [Edit](http://apps.webofknowledge.com/WOS_AdvancedSearch_input.do?product=WOS&SID=8EMF6vM4VEGGXsb3ob8&search_mode=AdvancedSearch&replaceSetId=25&editState=init) |
| # 24 | [2,338](http://apps.webofknowledge.com/summary.do;jsessionid=F8E63CDD2A44FEADE42E163384D2A58D?product=WOS&doc=1&qid=33&SID=8EMF6vM4VEGGXsb3ob8&search_mode=CombineSearches&update_back2search_link_param=yes) | #23 OR #22 OR #21 OR #20 OR #19 OR #18  Indexes=SCI-EXPANDED, SSCI, A&HCI, ESCI Timespan=1900-2020 | [Edit](http://apps.webofknowledge.com/WOS_AdvancedSearch_input.do?product=WOS&SID=8EMF6vM4VEGGXsb3ob8&search_mode=AdvancedSearch&replaceSetId=24&editState=init) |
| # 23 | [835](http://apps.webofknowledge.com/summary.do;jsessionid=F8E63CDD2A44FEADE42E163384D2A58D?product=WOS&doc=1&qid=32&SID=8EMF6vM4VEGGXsb3ob8&search_mode=CombineSearches&update_back2search_link_param=yes) | #5 AND #4 AND #1  Indexes=SCI-EXPANDED, SSCI, A&HCI, ESCI Timespan=1900-2020 | [Edit](http://apps.webofknowledge.com/WOS_AdvancedSearch_input.do?product=WOS&SID=8EMF6vM4VEGGXsb3ob8&search_mode=AdvancedSearch&replaceSetId=23&editState=init) |
| # 22 | [1,750](http://apps.webofknowledge.com/summary.do;jsessionid=F8E63CDD2A44FEADE42E163384D2A58D?product=WOS&doc=1&qid=31&SID=8EMF6vM4VEGGXsb3ob8&search_mode=CombineSearches&update_back2search_link_param=yes) | #15 AND #1  Indexes=SCI-EXPANDED, SSCI, A&HCI, ESCI Timespan=1900-2020 | [Edit](http://apps.webofknowledge.com/WOS_AdvancedSearch_input.do?product=WOS&SID=8EMF6vM4VEGGXsb3ob8&search_mode=AdvancedSearch&replaceSetId=22&editState=init) |
| # 21 | [1,170](http://apps.webofknowledge.com/summary.do;jsessionid=F8E63CDD2A44FEADE42E163384D2A58D?product=WOS&doc=1&qid=30&SID=8EMF6vM4VEGGXsb3ob8&search_mode=CombineSearches&update_back2search_link_param=yes) | #16 AND #11 AND #1  Indexes=SCI-EXPANDED, SSCI, A&HCI, ESCI Timespan=1900-2020 | [Edit](http://apps.webofknowledge.com/WOS_AdvancedSearch_input.do?product=WOS&SID=8EMF6vM4VEGGXsb3ob8&search_mode=AdvancedSearch&replaceSetId=21&editState=init) |
| # 20 | [222](http://apps.webofknowledge.com/summary.do;jsessionid=F8E63CDD2A44FEADE42E163384D2A58D?product=WOS&doc=1&qid=29&SID=8EMF6vM4VEGGXsb3ob8&search_mode=CombineSearches&update_back2search_link_param=yes) | #16 AND #2 AND #1  Indexes=SCI-EXPANDED, SSCI, A&HCI, ESCI Timespan=1900-2020 | [Edit](http://apps.webofknowledge.com/WOS_AdvancedSearch_input.do?product=WOS&SID=8EMF6vM4VEGGXsb3ob8&search_mode=AdvancedSearch&replaceSetId=20&editState=init) |
| # 19 | [137](http://apps.webofknowledge.com/summary.do;jsessionid=F8E63CDD2A44FEADE42E163384D2A58D?product=WOS&doc=1&qid=28&SID=8EMF6vM4VEGGXsb3ob8&search_mode=CombineSearches&update_back2search_link_param=yes) | #3 AND #1  Indexes=SCI-EXPANDED, SSCI, A&HCI, ESCI Timespan=1900-2020 | [Edit](http://apps.webofknowledge.com/WOS_AdvancedSearch_input.do?product=WOS&SID=8EMF6vM4VEGGXsb3ob8&search_mode=AdvancedSearch&replaceSetId=19&editState=init) |
| # 18 | [54](http://apps.webofknowledge.com/summary.do;jsessionid=F8E63CDD2A44FEADE42E163384D2A58D?product=WOS&doc=1&qid=27&SID=8EMF6vM4VEGGXsb3ob8&search_mode=CombineSearches&update_back2search_link_param=yes) | #17 AND #12 AND #1  Indexes=SCI-EXPANDED, SSCI, A&HCI, ESCI Timespan=1900-2020 | [Edit](http://apps.webofknowledge.com/WOS_AdvancedSearch_input.do?product=WOS&SID=8EMF6vM4VEGGXsb3ob8&search_mode=AdvancedSearch&replaceSetId=18&editState=init) |
| # 17 | [657,469](http://apps.webofknowledge.com/summary.do;jsessionid=F8E63CDD2A44FEADE42E163384D2A58D?product=WOS&doc=1&qid=26&SID=8EMF6vM4VEGGXsb3ob8&search_mode=GeneralSearch&update_back2search_link_param=yes) | **TOPIC:** (balance or posture or postural)  Indexes=SCI-EXPANDED, SSCI, A&HCI, ESCI Timespan=1900-2020 | [Edit](http://apps.webofknowledge.com/WOS_AdvancedSearch_input.do?product=WOS&SID=8EMF6vM4VEGGXsb3ob8&search_mode=AdvancedSearch&replaceSetId=17&editState=init) |
| # 16 | [405,431](http://apps.webofknowledge.com/summary.do;jsessionid=F8E63CDD2A44FEADE42E163384D2A58D?product=WOS&doc=1&qid=24&SID=8EMF6vM4VEGGXsb3ob8&search_mode=CombineSearches&update_back2search_link_param=yes) | #15 OR #14 OR #13 OR #12  Indexes=SCI-EXPANDED, SSCI, A&HCI, ESCI Timespan=1900-2020 | [Edit](http://apps.webofknowledge.com/WOS_AdvancedSearch_input.do?product=WOS&SID=8EMF6vM4VEGGXsb3ob8&search_mode=AdvancedSearch&replaceSetId=16&editState=init) |
| # 15 | [121,101](http://apps.webofknowledge.com/summary.do;jsessionid=F8E63CDD2A44FEADE42E163384D2A58D?product=WOS&doc=1&qid=23&SID=8EMF6vM4VEGGXsb3ob8&search_mode=GeneralSearch&update_back2search_link_param=yes) | **TOPIC:** (((balance or gait or perturbation* or postural or posture or propriocept* or sensorimotor* or slip or slipping or stance or stand or standing or step or stepping or trip or tripping or walk or walking) NEAR/5 (assess* or dual-task or "dual task" or simulat* or task or tasks or test or tests or testing)))  Indexes=SCI-EXPANDED, SSCI, A&HCI, ESCI Timespan=1900-2020 | [Edit](http://apps.webofknowledge.com/WOS_AdvancedSearch_input.do?product=WOS&SID=8EMF6vM4VEGGXsb3ob8&search_mode=AdvancedSearch&replaceSetId=15&editState=init) |
| # 14 | [213](http://apps.webofknowledge.com/summary.do;jsessionid=F8E63CDD2A44FEADE42E163384D2A58D?product=WOS&doc=1&qid=21&SID=8EMF6vM4VEGGXsb3ob8&search_mode=GeneralSearch&update_back2search_link_param=yes) | **TOPIC:** ((("virtual reality" or vr) NEAR/3 (balance or propriocept* or posture or postural)))  Indexes=SCI-EXPANDED, SSCI, A&HCI, ESCI Timespan=1900-2020 | [Edit](http://apps.webofknowledge.com/WOS_AdvancedSearch_input.do?product=WOS&SID=8EMF6vM4VEGGXsb3ob8&search_mode=AdvancedSearch&replaceSetId=14&editState=init) |
| # 13 | [21,902](http://apps.webofknowledge.com/summary.do;jsessionid=F8E63CDD2A44FEADE42E163384D2A58D?product=WOS&doc=1&qid=20&SID=8EMF6vM4VEGGXsb3ob8&search_mode=GeneralSearch&update_back2search_link_param=yes) | **TOPIC:** ((ActiveStep or BESTest or CAREN or "Computer Assisted Rehabilitation Environment" or "center of pressure" or "centre of pressure" or "central sensorimotor integration task" or GVS or "galvanic vestibular stimulation" or "heel contact latenc*" or "lean-and-release" or "lean and release" or "lean & release" or "limits of stability" or "mini BEST" or miniBEST or (motion NEAR/3 (track* or control* or test*)) or MCT or "motor control test" or neurocom or posturograph* or "Push-and-release" or "push and release" or "push & release" or P&R or "P and R" or "release-from-lean" or "release from lean" or Simbex or "SMART EquiTest" or "stance slip" or SVS or "vestibular stimulation" or "support surface" or (trunk NEAR/5 (angle or sway)) or VEMP or "vestibular evoked myogenic potential*" or vibrat* or "waist belt" or (waist NEAR/5 belt)).ti,ab,kf. or ((ground or forceplate or "force plate" or platform or surface) NEAR/5 (moving or tilt or translation*)))  Indexes=SCI-EXPANDED, SSCI, A&HCI, ESCI Timespan=1900-2020 | [Edit](http://apps.webofknowledge.com/WOS_AdvancedSearch_input.do?product=WOS&SID=8EMF6vM4VEGGXsb3ob8&search_mode=AdvancedSearch&replaceSetId=13&editState=init) |
| # 12 | [270,733](http://apps.webofknowledge.com/summary.do;jsessionid=F8E63CDD2A44FEADE42E163384D2A58D?product=WOS&doc=1&qid=19&SID=8EMF6vM4VEGGXsb3ob8&search_mode=GeneralSearch&update_back2search_link_param=yes) | **TOPIC:** (perturbation*)  Indexes=SCI-EXPANDED, SSCI, A&HCI, ESCI Timespan=1900-2020 | [Edit](http://apps.webofknowledge.com/WOS_AdvancedSearch_input.do?product=WOS&SID=8EMF6vM4VEGGXsb3ob8&search_mode=AdvancedSearch&replaceSetId=12&editState=init) |
| # 11 | [1,565,941](http://apps.webofknowledge.com/summary.do;jsessionid=F8E63CDD2A44FEADE42E163384D2A58D?product=WOS&doc=1&qid=18&SID=8EMF6vM4VEGGXsb3ob8&search_mode=CombineSearches&update_back2search_link_param=yes) | #10 OR #9 OR #8 OR #7 OR #6 OR #5  Indexes=SCI-EXPANDED, SSCI, A&HCI, ESCI Timespan=1900-2020 | [Edit](http://apps.webofknowledge.com/WOS_AdvancedSearch_input.do?product=WOS&SID=8EMF6vM4VEGGXsb3ob8&search_mode=AdvancedSearch&replaceSetId=11&editState=init) |
| # 10 | [49,336](http://apps.webofknowledge.com/summary.do;jsessionid=F8E63CDD2A44FEADE42E163384D2A58D?product=WOS&doc=1&qid=17&SID=8EMF6vM4VEGGXsb3ob8&search_mode=GeneralSearch&update_back2search_link_param=yes) | **TOPIC:** ((Neurolabyrinthiti* or "Scarpa Ganglion*" or "Scarpas Ganglion*" or "Scarpa's Ganglion*" or vestibul* or vertigo*))  Indexes=SCI-EXPANDED, SSCI, A&HCI, ESCI Timespan=1900-2020 | [Edit](http://apps.webofknowledge.com/WOS_AdvancedSearch_input.do?product=WOS&SID=8EMF6vM4VEGGXsb3ob8&search_mode=AdvancedSearch&replaceSetId=10&editState=init) |
| # 9 | [5,630](http://apps.webofknowledge.com/summary.do;jsessionid=F8E63CDD2A44FEADE42E163384D2A58D?product=WOS&doc=1&qid=16&SID=8EMF6vM4VEGGXsb3ob8&search_mode=GeneralSearch&update_back2search_link_param=yes) | **TOPIC:** (((sensory or sensorimotor or startle) NEAR/3 (gating or filtering)).ti,ab. or ((prepulse or pre-pulse) NEAR/2 (facilitate* or inhibit*)))  Indexes=SCI-EXPANDED, SSCI, A&HCI, ESCI Timespan=1900-2020 | [Edit](http://apps.webofknowledge.com/WOS_AdvancedSearch_input.do?product=WOS&SID=8EMF6vM4VEGGXsb3ob8&search_mode=AdvancedSearch&replaceSetId=9&editState=init) |
| # 8 | [13,185](http://apps.webofknowledge.com/summary.do;jsessionid=F8E63CDD2A44FEADE42E163384D2A58D?product=WOS&doc=1&qid=15&SID=8EMF6vM4VEGGXsb3ob8&search_mode=GeneralSearch&update_back2search_link_param=yes) | **TOPIC:** (((acoustic* or audio* or haptic or propriocept* or sensorimotor or sensory or tactile or visual) NEAR/3 (feedback or feedbacks)))  Indexes=SCI-EXPANDED, SSCI, A&HCI, ESCI Timespan=1900-2020 | [Edit](http://apps.webofknowledge.com/WOS_AdvancedSearch_input.do?product=WOS&SID=8EMF6vM4VEGGXsb3ob8&search_mode=AdvancedSearch&replaceSetId=8&editState=init) |
| # 7 | [124,259](http://apps.webofknowledge.com/summary.do;jsessionid=F8E63CDD2A44FEADE42E163384D2A58D?product=WOS&doc=1&qid=12&SID=8EMF6vM4VEGGXsb3ob8&search_mode=GeneralSearch&update_back2search_link_param=yes) | **TOPIC:** ((("anterior parietal" or Brodmann or extrastriat* or motor or motorsensory or premotor or "post central" or postcentral* or precentral* or "pre central" or Rolandic or sensory or sensorimotor or "sensory motor" or "sensory-motor" or si or sensomotor or somatomotor or somatosensory or "somatic sensory" or striate or visual) NEAR/3 (area or areas or cortex or cortices or gyrus or region or strip or strips or zone)))  Indexes=SCI-EXPANDED, SSCI, A&HCI, ESCI Timespan=1900-2020 | [Edit](http://apps.webofknowledge.com/WOS_AdvancedSearch_input.do?product=WOS&SID=8EMF6vM4VEGGXsb3ob8&search_mode=AdvancedSearch&replaceSetId=7&editState=init) |
| # 6 | [122,710](http://apps.webofknowledge.com/summary.do;jsessionid=F8E63CDD2A44FEADE42E163384D2A58D?product=WOS&doc=1&qid=9&SID=8EMF6vM4VEGGXsb3ob8&search_mode=GeneralSearch&update_back2search_link_param=yes) | **TOPIC:** (("auris interna" or "acoustic macula*" or "basilar papilla" or cochlea* or endolymph* or "fenestra rotunda*" or "internal acoustic pore" or "internal auditory meatus" or labyrinth* or "meatus acusticus" or "macula* acoustic" or otoconia* or otolith* or "papilla acustica basilaris" or "papilla basilar" or perilymph* or "porus acusticus internus" or saccule* or sacculus or "stria vascularis" or statoconi* or statocyst or statocyte or statolith or utricle* or utriculus or "vascularis stria" or vestibul* or ((ear or ears) NEAR/2 (helix or inner or internal or vestibul*)) or ((cochlea* or endolymphatic or perilymphatic or semicircular) NEAR/2 duct*) or (scala NEAR/2 (media* or tympani or tympanus or vestibuli or vestibulus)) or ((basilar or otolithic or statoconia or tectorial) NEAR/2 (lamina or membrane*)) or ((acoustic or auditory or corti* or spiral*) NEAR/2 (gangli* or lamina or laminas or ligament or ligaments or organ or organs or organum)) or ((ampullar* or auditory or cochlea* or inner or outer or vestibular) NEAR/2 ("hair cell" or "hair cells")) or ((auditory or cochlea* or ear or semicircular or vestibul*) NEAR/2 (apparatus or aqueduct* or auris or canal or canals or canaliculus or "oval window*" or "round window*")) or (ampullar* NEAR/2 (crest or crests or crista))))  Indexes=SCI-EXPANDED, SSCI, A&HCI, ESCI Timespan=1900-2020 | [Edit](http://apps.webofknowledge.com/WOS_AdvancedSearch_input.do?product=WOS&SID=8EMF6vM4VEGGXsb3ob8&search_mode=AdvancedSearch&replaceSetId=6&editState=init) |
| # 5 | [1,340,395](http://apps.webofknowledge.com/summary.do;jsessionid=F8E63CDD2A44FEADE42E163384D2A58D?product=WOS&doc=1&qid=6&SID=8EMF6vM4VEGGXsb3ob8&search_mode=GeneralSearch&update_back2search_link_param=yes) | **TOPIC:** ((balance or "body sway" or "deep sensitivity" or imbalance or kinesthes* or perception* or propriocept* or propriocepsis or sensation* or ((body or kinesthetic or kinaesthetic or movement or position) NEAR/3 (discrimination or percept* or sense* or sensory or sensation*)) or ((balance or body or gait or posture or postural or musculoskeletal) NEAR/3 (abnormal* or adaption* or adjust* or control or difficult* or disequilibrium or disturbance* or entropy or equilibrium or error or impair* or instabilit* or oscillat* or orient* or react* or recover* or response* or restor* or stabili* or stance or steadiness or steady or sway* or unsteadiness or unsteady)) or (sensory NEAR/2 (function or functions))))  Indexes=SCI-EXPANDED, SSCI, A&HCI, ESCI Timespan=1900-2020 | [Edit](http://apps.webofknowledge.com/WOS_AdvancedSearch_input.do?product=WOS&SID=8EMF6vM4VEGGXsb3ob8&search_mode=AdvancedSearch&replaceSetId=5&editState=init) |
| # 4 | [71,414](http://apps.webofknowledge.com/summary.do;jsessionid=F8E63CDD2A44FEADE42E163384D2A58D?product=WOS&doc=1&qid=4&SID=8EMF6vM4VEGGXsb3ob8&search_mode=GeneralSearch&update_back2search_link_param=yes) | **TOPIC:** ((posture or postural))  Indexes=SCI-EXPANDED, SSCI, A&HCI, ESCI Timespan=1900-2020 | [Edit](http://apps.webofknowledge.com/WOS_AdvancedSearch_input.do?product=WOS&SID=8EMF6vM4VEGGXsb3ob8&search_mode=AdvancedSearch&replaceSetId=4&editState=init) |
| # 3 | [12,836](http://apps.webofknowledge.com/summary.do;jsessionid=F8E63CDD2A44FEADE42E163384D2A58D?product=WOS&doc=1&qid=3&SID=8EMF6vM4VEGGXsb3ob8&search_mode=GeneralSearch&update_back2search_link_param=yes) | **TOPIC:** ((dynamic NEAR/5 (balance or posture or postural)))  Indexes=SCI-EXPANDED, SSCI, A&HCI, ESCI Timespan=1900-2020 | [Edit](http://apps.webofknowledge.com/WOS_AdvancedSearch_input.do?product=WOS&SID=8EMF6vM4VEGGXsb3ob8&search_mode=AdvancedSearch&replaceSetId=3&editState=init) |
| # 2 | [3,301,955](http://apps.webofknowledge.com/summary.do;jsessionid=F8E63CDD2A44FEADE42E163384D2A58D?product=WOS&doc=1&qid=2&SID=8EMF6vM4VEGGXsb3ob8&search_mode=GeneralSearch&update_back2search_link_param=yes) | **TOPIC:** ((reacti* or anticipatory or compensatory))  Indexes=SCI-EXPANDED, SSCI, A&HCI, ESCI Timespan=1900-2020 | [Edit](http://apps.webofknowledge.com/WOS_AdvancedSearch_input.do?product=WOS&SID=8EMF6vM4VEGGXsb3ob8&search_mode=AdvancedSearch&replaceSetId=2&editState=init) |
| # 1 | [157,782](http://apps.webofknowledge.com/summary.do;jsessionid=F8E63CDD2A44FEADE42E163384D2A58D?product=WOS&doc=1&qid=1&SID=8EMF6vM4VEGGXsb3ob8&search_mode=GeneralSearch&update_back2search_link_param=yes) | **TOPIC:** ((concuss* or commotio or "dementia pugulistica" or mtbi or mtbis or post-concuss* or postconcuss* or sub-concuss* or subconcuss* or tbi or tbis or ((brain or cerebell* or cerebral or cerebrovascular or cortical or encephalopath* or head) NEAR/3 (commotio* or concuss* or contusion* or impact* or injur* or postconcuss* or post-concuss* or posttraumatic or post-traumatic or trauma*))))  Indexes=SCI-EXPANDED, SSCI, A&HCI, ESCI Timespan=All years |  |

------

Re-ran search on March 9, 2021

Refined by: PUBLICATION YEARS: ( 2021 OR 2020 )	253

#### **Dissertations & Theses Global (ProQuest) 2020-02-04**

| **Select all** | [**Set**](https://search-proquest-com.ezproxy.lib.utah.edu/recentsearches.recentsearchtabview.recentsearchesgridview:toggellistorder?site=pqdtglobal&t:ac=RecentSearches) | **Search** | **Databases** | **Results** |
| --- | --- | --- | --- | --- |
| Select item 19 | **S19** | [((noft((concuss* OR commotio OR "dementia pugulistica" OR mtbi OR mtbis OR post-concuss* OR postconcuss* OR sub-concuss* OR subconcuss* OR tbi OR tbis OR ((brain OR cerebell* OR cerebral OR cerebrovascular OR cortical OR encephalopath* OR head) NEAR/3 (commotio* OR concuss* OR contusion* OR impact* OR injur* OR postconcuss* OR post-concuss* OR posttraumatic OR post-traumatic OR trauma*)))) AND noft(perturbation*) AND noft(balance or posture or postural)) OR (noft((concuss* OR commotio OR "dementia pugulistica" OR mtbi OR mtbis OR post-concuss* OR postconcuss* OR sub-concuss* OR subconcuss* OR tbi OR tbis OR ((brain OR cerebell* OR cerebral OR cerebrovascular OR cortical OR encephalopath* OR head) NEAR/3 (commotio* OR concuss* OR contusion* OR impact* OR injur* OR postconcuss* OR post-concuss* OR posttraumatic OR post-traumatic OR trauma*)))) AND noft(dynamic NEAR/5 (balance or posture or postural))) OR (noft((concuss* OR commotio OR "dementia pugulistica" OR mtbi OR mtbis OR post-concuss* OR postconcuss* OR sub-concuss* OR subconcuss* OR tbi OR tbis OR ((brain OR cerebell* OR cerebral OR cerebrovascular OR cortical OR encephalopath* OR head) NEAR/3 (commotio* OR concuss* OR contusion* OR impact* OR injur* OR postconcuss* OR post-concuss* OR posttraumatic OR post-traumatic OR trauma*)))) AND noft(reacti* or anticipatory or compensatory) AND noft(balance or "body sway" or "deep sensitivity" or imbalance or kinesthes* or perception* or propriocept* or propriocepsis or sensation* or ((body or kinesthetic or kinaesthetic or movement or position) NEAR/3 (discrimination or percept* or sense* or sensory or sensation*)) or ((balance or body or gait or posture or postural or musculoskeletal) NEAR/3 (abnormal* or adaption* or adjust* or control or difficult* or disequilibrium or disturbance* or entropy or equilibrium or error or impair* or instabilit* or oscillat* or orient* or react* or recover* or response* or restor* or stabili* or stance or steadiness or steady or sway* or unsteadiness or unsteady)) or (sensory NEAR/2 (function or functions)))) OR (noft((concuss* OR commotio OR "dementia pugulistica" OR mtbi OR mtbis OR post-concuss* OR postconcuss* OR sub-concuss* OR subconcuss* OR tbi OR tbis OR ((brain OR cerebell* OR cerebral OR cerebrovascular OR cortical OR encephalopath* OR head) NEAR/3 (commotio* OR concuss* OR contusion* OR impact* OR injur* OR postconcuss* OR post-concuss* OR posttraumatic OR post-traumatic OR trauma*)))) AND noft(reacti* or anticipatory or compensatory) AND (noft(perturbation*) OR (noft(ActiveStep or BESTest or CAREN or "Computer Assisted Rehabilitation Environment" or "center of pressure" or "centre of pressure" or "central sensorimotor integration task" or GVS or "galvanic vestibular stimulation" or "heel contact latenc*" or "lean-and-release" or "lean and release" or "lean & release" or "limits of stability" or "mini BEST" or miniBEST or (motion NEAR/3 (track* or control* or test*)) or MCT or "motor control test" or neurocom or posturograph* or "Push-and-release" or "push and release" or "push & release" or P&R or "P and R" or "release-from-lean" or "release from lean" or Simbex or "SMART EquiTest" or "stance slip" or SVS or "vestibular stimulation" or "support surface" or (trunk NEAR/5 (angle or sway)) or VEMP or "vestibular evoked myogenic potential*" or vibrat* or "waist belt" or (waist NEAR/5 belt)).ti,ab,kf. or ((ground or forceplate or "force plate" or platform or surface) NEAR/5 (moving or tilt or translation*))) OR noft(("virtual reality" or vr) NEAR/3 (balance or propriocept* or posture or postural)) OR noft((balance or gait or perturbation* or postural or posture or propriocept* or sensorimotor* or slip or slipping or stance or stand or standing or step or stepping or trip or tripping or walk or walking) NEAR/5 (assess* or dual-task or "dual task" or simulat* or task or tasks or test or tests or testing)))) OR (noft((concuss* OR commotio OR "dementia pugulistica" OR mtbi OR mtbis OR post-concuss* OR postconcuss* OR sub-concuss* OR subconcuss* OR tbi OR tbis OR ((brain OR cerebell* OR cerebral OR cerebrovascular OR cortical OR encephalopath* OR head) NEAR/3 (commotio* OR concuss* OR contusion* OR impact* OR injur* OR postconcuss* OR post-concuss* OR posttraumatic OR post-traumatic OR trauma*)))) AND noft((balance or gait or perturbation* or postural or posture or propriocept* or sensorimotor* or slip or slipping or stance or stand or standing or step or stepping or trip or tripping or walk or walking) NEAR/5 (assess* or dual-task or "dual task" or simulat* or task or tasks or test or tests or testing))) OR (noft((concuss* OR commotio OR "dementia pugulistica" OR mtbi OR mtbis OR post-concuss* OR postconcuss* OR sub-concuss* OR subconcuss* OR tbi OR tbis OR ((brain OR cerebell* OR cerebral OR cerebrovascular OR cortical OR encephalopath* OR head) NEAR/3 (commotio* OR concuss* OR contusion* OR impact* OR injur* OR postconcuss* OR post-concuss* OR posttraumatic OR post-traumatic OR trauma*)))) AND noft(posture or postural) AND noft(balance or "body sway" or "deep sensitivity" or imbalance or kinesthes* or perception* or propriocept* or propriocepsis or sensation* or ((body or kinesthetic or kinaesthetic or movement or position) NEAR/3 (discrimination or percept* or sense* or sensory or sensation*)) or ((balance or body or gait or posture or postural or musculoskeletal) NEAR/3 (abnormal* or adaption* or adjust* or control or difficult* or disequilibrium or disturbance* or entropy or equilibrium or error or impair* or instabilit* or oscillat* or orient* or react* or recover* or response* or restor* or stabili* or stance or steadiness or steady or sway* or unsteadiness or unsteady)) or (sensory NEAR/2 (function or functions))))) NOT ti(animal OR animals OR beaver OR beavers OR beef OR bovine OR breeding OR bull OR canine OR castoris OR cat OR cattle OR cats OR chicken OR chickens OR chimp* OR cow OR dog OR dogs OR equine OR foal OR foals OR fish OR insect? OR horse OR horses OR livestock OR mice OR monkey OR monkeys OR mouse OR murine OR plant OR plants OR pork OR porcine OR protozoa OR protozoas OR purebred OR rabbit OR rabbits OR rat OR rats OR rodent OR rodents OR sheep OR thoroughbred OR veterinar*)](https://search-proquest-com.ezproxy.lib.utah.edu/recentsearches.recentsearchtabview.recentsearchesgridview.scrolledrecentsearchlist.checkdbssearchlink:rerunsearch/CA9A916FD0174004PQ/None?site=pqdtglobal&t:ac=RecentSearches) | ProQuest Dissertations & Theses Global | [**276**](https://search-proquest-com.ezproxy.lib.utah.edu/recentsearches.recentsearchtabview.recentsearchesgridview.scrolledrecentsearchlist.checkdbssearchlink_0:rerunsearch/CA9A916FD0174004PQ/None?site=pqdtglobal&t:ac=RecentSearches) |
| Select item 18 | **S18** | [ti(animal or animals or beaver or beavers or beef or bovine or breeding or bull or canine or castoris or cat or cattle or cats or chicken or chickens or chimp* or cow or dog or dogs or equine or foal or foals or fish or insect? or horse or horses or livestock or mice or monkey or monkeys or mouse or murine or plant or plants or pork or porcine or protozoa or protozoas or purebred or rabbit or rabbits or rat or rats or rodent or rodents or sheep or thoroughbred or veterinar*)](https://search-proquest-com.ezproxy.lib.utah.edu/recentsearches.recentsearchtabview.recentsearchesgridview.scrolledrecentsearchlist.checkdbssearchlink:rerunsearch/9BF0574CC3634879PQ/None?site=pqdtglobal&t:ac=RecentSearches) | ProQuest Dissertations & Theses Global | [**144,530**](https://search-proquest-com.ezproxy.lib.utah.edu/recentsearches.recentsearchtabview.recentsearchesgridview.scrolledrecentsearchlist.checkdbssearchlink_0:rerunsearch/9BF0574CC3634879PQ/None?site=pqdtglobal&t:ac=RecentSearches) |
| Select item 17 | **S17** | [(noft((concuss* OR commotio OR "dementia pugulistica" OR mtbi OR mtbis OR post-concuss* OR postconcuss* OR sub-concuss* OR subconcuss* OR tbi OR tbis OR ((brain OR cerebell* OR cerebral OR cerebrovascular OR cortical OR encephalopath* OR head) NEAR/3 (commotio* OR concuss* OR contusion* OR impact* OR injur* OR postconcuss* OR post-concuss* OR posttraumatic OR post-traumatic OR trauma*)))) AND noft(perturbation*) AND noft(balance or posture or postural)) OR (noft((concuss* OR commotio OR "dementia pugulistica" OR mtbi OR mtbis OR post-concuss* OR postconcuss* OR sub-concuss* OR subconcuss* OR tbi OR tbis OR ((brain OR cerebell* OR cerebral OR cerebrovascular OR cortical OR encephalopath* OR head) NEAR/3 (commotio* OR concuss* OR contusion* OR impact* OR injur* OR postconcuss* OR post-concuss* OR posttraumatic OR post-traumatic OR trauma*)))) AND noft(dynamic NEAR/5 (balance or posture or postural))) OR (noft((concuss* OR commotio OR "dementia pugulistica" OR mtbi OR mtbis OR post-concuss* OR postconcuss* OR sub-concuss* OR subconcuss* OR tbi OR tbis OR ((brain OR cerebell* OR cerebral OR cerebrovascular OR cortical OR encephalopath* OR head) NEAR/3 (commotio* OR concuss* OR contusion* OR impact* OR injur* OR postconcuss* OR post-concuss* OR posttraumatic OR post-traumatic OR trauma*)))) AND noft(reacti* or anticipatory or compensatory) AND noft(balance or "body sway" or "deep sensitivity" or imbalance or kinesthes* or perception* or propriocept* or propriocepsis or sensation* or ((body or kinesthetic or kinaesthetic or movement or position) NEAR/3 (discrimination or percept* or sense* or sensory or sensation*)) or ((balance or body or gait or posture or postural or musculoskeletal) NEAR/3 (abnormal* or adaption* or adjust* or control or difficult* or disequilibrium or disturbance* or entropy or equilibrium or error or impair* or instabilit* or oscillat* or orient* or react* or recover* or response* or restor* or stabili* or stance or steadiness or steady or sway* or unsteadiness or unsteady)) or (sensory NEAR/2 (function or functions)))) OR (noft((concuss* OR commotio OR "dementia pugulistica" OR mtbi OR mtbis OR post-concuss* OR postconcuss* OR sub-concuss* OR subconcuss* OR tbi OR tbis OR ((brain OR cerebell* OR cerebral OR cerebrovascular OR cortical OR encephalopath* OR head) NEAR/3 (commotio* OR concuss* OR contusion* OR impact* OR injur* OR postconcuss* OR post-concuss* OR posttraumatic OR post-traumatic OR trauma*)))) AND noft(reacti* or anticipatory or compensatory) AND (noft(perturbation*) OR (noft(ActiveStep or BESTest or CAREN or "Computer Assisted Rehabilitation Environment" or "center of pressure" or "centre of pressure" or "central sensorimotor integration task" or GVS or "galvanic vestibular stimulation" or "heel contact latenc*" or "lean-and-release" or "lean and release" or "lean & release" or "limits of stability" or "mini BEST" or miniBEST or (motion NEAR/3 (track* or control* or test*)) or MCT or "motor control test" or neurocom or posturograph* or "Push-and-release" or "push and release" or "push & release" or P&R or "P and R" or "release-from-lean" or "release from lean" or Simbex or "SMART EquiTest" or "stance slip" or SVS or "vestibular stimulation" or "support surface" or (trunk NEAR/5 (angle or sway)) or VEMP or "vestibular evoked myogenic potential*" or vibrat* or "waist belt" or (waist NEAR/5 belt)).ti,ab,kf. or ((ground or forceplate or "force plate" or platform or surface) NEAR/5 (moving or tilt or translation*))) OR noft(("virtual reality" or vr) NEAR/3 (balance or propriocept* or posture or postural)) OR noft((balance or gait or perturbation* or postural or posture or propriocept* or sensorimotor* or slip or slipping or stance or stand or standing or step or stepping or trip or tripping or walk or walking) NEAR/5 (assess* or dual-task or "dual task" or simulat* or task or tasks or test or tests or testing)))) OR (noft((concuss* OR commotio OR "dementia pugulistica" OR mtbi OR mtbis OR post-concuss* OR postconcuss* OR sub-concuss* OR subconcuss* OR tbi OR tbis OR ((brain OR cerebell* OR cerebral OR cerebrovascular OR cortical OR encephalopath* OR head) NEAR/3 (commotio* OR concuss* OR contusion* OR impact* OR injur* OR postconcuss* OR post-concuss* OR posttraumatic OR post-traumatic OR trauma*)))) AND noft((balance or gait or perturbation* or postural or posture or propriocept* or sensorimotor* or slip or slipping or stance or stand or standing or step or stepping or trip or tripping or walk or walking) NEAR/5 (assess* or dual-task or "dual task" or simulat* or task or tasks or test or tests or testing))) OR (noft((concuss* OR commotio OR "dementia pugulistica" OR mtbi OR mtbis OR post-concuss* OR postconcuss* OR sub-concuss* OR subconcuss* OR tbi OR tbis OR ((brain OR cerebell* OR cerebral OR cerebrovascular OR cortical OR encephalopath* OR head) NEAR/3 (commotio* OR concuss* OR contusion* OR impact* OR injur* OR postconcuss* OR post-concuss* OR posttraumatic OR post-traumatic OR trauma*)))) AND noft(posture or postural) AND noft(balance or "body sway" or "deep sensitivity" or imbalance or kinesthes* or perception* or propriocept* or propriocepsis or sensation* or ((body or kinesthetic or kinaesthetic or movement or position) NEAR/3 (discrimination or percept* or sense* or sensory or sensation*)) or ((balance or body or gait or posture or postural or musculoskeletal) NEAR/3 (abnormal* or adaption* or adjust* or control or difficult* or disequilibrium or disturbance* or entropy or equilibrium or error or impair* or instabilit* or oscillat* or orient* or react* or recover* or response* or restor* or stabili* or stance or steadiness or steady or sway* or unsteadiness or unsteady)) or (sensory NEAR/2 (function or functions))))](https://search-proquest-com.ezproxy.lib.utah.edu/recentsearches.recentsearchtabview.recentsearchesgridview.scrolledrecentsearchlist.checkdbssearchlink:rerunsearch/587D7DF82DDD4F43PQ/None?site=pqdtglobal&t:ac=RecentSearches) | ProQuest Dissertations & Theses Global | [**302**](https://search-proquest-com.ezproxy.lib.utah.edu/recentsearches.recentsearchtabview.recentsearchesgridview.scrolledrecentsearchlist.checkdbssearchlink_0:rerunsearch/587D7DF82DDD4F43PQ/None?site=pqdtglobal&t:ac=RecentSearches) |
| Select item 16 | **S16** | noft((concuss* OR commotio OR "dementia pugulistica" OR mtbi OR mtbis OR post-concuss* OR postconcuss* OR sub-concuss* OR subconcuss* OR tbi OR tbis OR ((brain OR cerebell* OR cerebral OR cerebrovascular OR cortical OR encephalopath* OR head) NEAR/3 (commotio* OR concuss* OR contusion* OR impact* OR injur* OR postconcuss* OR post-concuss* OR posttraumatic OR post-traumatic OR trauma*)))) AND noft(posture or postural) AND noft(balance or "body sway" or "deep sensitivity" or imbalance or kinesthes* or perception* or propriocept* or propriocepsis or sensation* or ((body or kinesthetic or kinaesthetic or movement or position) NEAR/3 (discrimination or percept* or sense* or sensory or sensation*)) or ((balance or body or gait or posture or postural or musculoskeletal) NEAR/3 (abnormal* or adaption* or adjust* or control or difficult* or disequilibrium or disturbance* or entropy or equilibrium or error or impair* or instabilit* or oscillat* or orient* or react* or recover* or response* or restor* or stabili* or stance or steadiness or steady or sway* or unsteadiness or unsteady)) or (sensory NEAR/2 (function or functions))) | ProQuest Dissertations & Theses Global | [**81**](https://search-proquest-com.ezproxy.lib.utah.edu/recentsearches.recentsearchtabview.recentsearchesgridview.scrolledrecentsearchlist.checkdbssearchlink_0:rerunsearch/F31B613C2EC2432BPQ/None?site=pqdtglobal&t:ac=RecentSearches) |
| Select item 15 | **S15** | [noft((concuss* OR commotio OR "dementia pugulistica" OR mtbi OR mtbis OR post-concuss* OR postconcuss* OR sub-concuss* OR subconcuss* OR tbi OR tbis OR ((brain OR cerebell* OR cerebral OR cerebrovascular OR cortical OR encephalopath* OR head) NEAR/3 (commotio* OR concuss* OR contusion* OR impact* OR injur* OR postconcuss* OR post-concuss* OR posttraumatic OR post-traumatic OR trauma*)))) AND noft((balance or gait or perturbation* or postural or posture or propriocept* or sensorimotor* or slip or slipping or stance or stand or standing or step or stepping or trip or tripping or walk or walking) NEAR/5 (assess* or dual-task or "dual task" or simulat* or task or tasks or test or tests or testing))](https://search-proquest-com.ezproxy.lib.utah.edu/recentsearches.recentsearchtabview.recentsearchesgridview.scrolledrecentsearchlist.checkdbssearchlink:rerunsearch/D7A2BFC9BEBE4253PQ/None?site=pqdtglobal&t:ac=RecentSearches) | ProQuest Dissertations & Theses Global | [**181**](https://search-proquest-com.ezproxy.lib.utah.edu/recentsearches.recentsearchtabview.recentsearchesgridview.scrolledrecentsearchlist.checkdbssearchlink_0:rerunsearch/D7A2BFC9BEBE4253PQ/None?site=pqdtglobal&t:ac=RecentSearches) |
| Select item 14 | **S14** | [noft((concuss* OR commotio OR "dementia pugulistica" OR mtbi OR mtbis OR post-concuss* OR postconcuss* OR sub-concuss* OR subconcuss* OR tbi OR tbis OR ((brain OR cerebell* OR cerebral OR cerebrovascular OR cortical OR encephalopath* OR head) NEAR/3 (commotio* OR concuss* OR contusion* OR impact* OR injur* OR postconcuss* OR post-concuss* OR posttraumatic OR post-traumatic OR trauma*)))) AND noft(reacti* or anticipatory or compensatory) AND (noft(perturbation*) OR (noft(ActiveStep or BESTest or CAREN or "Computer Assisted Rehabilitation Environment" or "center of pressure" or "centre of pressure" or "central sensorimotor integration task" or GVS or "galvanic vestibular stimulation" or "heel contact latenc*" or "lean-and-release" or "lean and release" or "lean & release" or "limits of stability" or "mini BEST" or miniBEST or (motion NEAR/3 (track* or control* or test*)) or MCT or "motor control test" or neurocom or posturograph* or "Push-and-release" or "push and release" or "push & release" or P&R or "P and R" or "release-from-lean" or "release from lean" or Simbex or "SMART EquiTest" or "stance slip" or SVS or "vestibular stimulation" or "support surface" or (trunk NEAR/5 (angle or sway)) or VEMP or "vestibular evoked myogenic potential*" or vibrat* or "waist belt" or (waist NEAR/5 belt)).ti,ab,kf. or ((ground or forceplate or "force plate" or platform or surface) NEAR/5 (moving or tilt or translation*))) OR noft(("virtual reality" or vr) NEAR/3 (balance or propriocept* or posture or postural)) OR noft((balance or gait or perturbation* or postural or posture or propriocept* or sensorimotor* or slip or slipping or stance or stand or standing or step or stepping or trip or tripping or walk or walking) NEAR/5 (assess* or dual-task or "dual task" or simulat* or task or tasks or test or tests or testing)))](https://search-proquest-com.ezproxy.lib.utah.edu/recentsearches.recentsearchtabview.recentsearchesgridview.scrolledrecentsearchlist.checkdbssearchlink:rerunsearch/C068D98858604F75PQ/None?site=pqdtglobal&t:ac=RecentSearches) | ProQuest Dissertations & Theses Global | [**50**](https://search-proquest-com.ezproxy.lib.utah.edu/recentsearches.recentsearchtabview.recentsearchesgridview.scrolledrecentsearchlist.checkdbssearchlink_0:rerunsearch/C068D98858604F75PQ/None?site=pqdtglobal&t:ac=RecentSearches) |
| Select item 13 | **S13** | [noft((concuss* OR commotio OR "dementia pugulistica" OR mtbi OR mtbis OR post-concuss* OR postconcuss* OR sub-concuss* OR subconcuss* OR tbi OR tbis OR ((brain OR cerebell* OR cerebral OR cerebrovascular OR cortical OR encephalopath* OR head) NEAR/3 (commotio* OR concuss* OR contusion* OR impact* OR injur* OR postconcuss* OR post-concuss* OR posttraumatic OR post-traumatic OR trauma*)))) AND noft(reacti* or anticipatory or compensatory) AND noft(balance or "body sway" or "deep sensitivity" or imbalance or kinesthes* or perception* or propriocept* or propriocepsis or sensation* or ((body or kinesthetic or kinaesthetic or movement or position) NEAR/3 (discrimination or percept* or sense* or sensory or sensation*)) or ((balance or body or gait or posture or postural or musculoskeletal) NEAR/3 (abnormal* or adaption* or adjust* or control or difficult* or disequilibrium or disturbance* or entropy or equilibrium or error or impair* or instabilit* or oscillat* or orient* or react* or recover* or response* or restor* or stabili* or stance or steadiness or steady or sway* or unsteadiness or unsteady)) or (sensory NEAR/2 (function or functions)))](https://search-proquest-com.ezproxy.lib.utah.edu/recentsearches.recentsearchtabview.recentsearchesgridview.scrolledrecentsearchlist.checkdbssearchlink:rerunsearch/48FAB07BC2374861PQ/None?site=pqdtglobal&t:ac=RecentSearches) | ProQuest Dissertations & Theses Global | [**93**](https://search-proquest-com.ezproxy.lib.utah.edu/recentsearches.recentsearchtabview.recentsearchesgridview.scrolledrecentsearchlist.checkdbssearchlink_0:rerunsearch/48FAB07BC2374861PQ/None?site=pqdtglobal&t:ac=RecentSearches) |
| Select item 12 | **S12** | [noft((concuss* OR commotio OR "dementia pugulistica" OR mtbi OR mtbis OR post-concuss* OR postconcuss* OR sub-concuss* OR subconcuss* OR tbi OR tbis OR ((brain OR cerebell* OR cerebral OR cerebrovascular OR cortical OR encephalopath* OR head) NEAR/3 (commotio* OR concuss* OR contusion* OR impact* OR injur* OR postconcuss* OR post-concuss* OR posttraumatic OR post-traumatic OR trauma*)))) AND noft(dynamic NEAR/5 (balance or posture or postural))](https://search-proquest-com.ezproxy.lib.utah.edu/recentsearches.recentsearchtabview.recentsearchesgridview.scrolledrecentsearchlist.checkdbssearchlink:rerunsearch/7CE618359820415CPQ/None?site=pqdtglobal&t:ac=RecentSearches) | ProQuest Dissertations & Theses Global | [**17**](https://search-proquest-com.ezproxy.lib.utah.edu/recentsearches.recentsearchtabview.recentsearchesgridview.scrolledrecentsearchlist.checkdbssearchlink_0:rerunsearch/7CE618359820415CPQ/None?site=pqdtglobal&t:ac=RecentSearches) |
| Select item 11 | **S11** | [noft((concuss* OR commotio OR "dementia pugulistica" OR mtbi OR mtbis OR post-concuss* OR postconcuss* OR sub-concuss* OR subconcuss* OR tbi OR tbis OR ((brain OR cerebell* OR cerebral OR cerebrovascular OR cortical OR encephalopath* OR head) NEAR/3 (commotio* OR concuss* OR contusion* OR impact* OR injur* OR postconcuss* OR post-concuss* OR posttraumatic OR post-traumatic OR trauma*)))) AND noft(perturbation*) AND noft(balance or posture or postural)](https://search-proquest-com.ezproxy.lib.utah.edu/recentsearches.recentsearchtabview.recentsearchesgridview.scrolledrecentsearchlist.checkdbssearchlink:rerunsearch/5FE963EE313146A8PQ/None?site=pqdtglobal&t:ac=RecentSearches) | ProQuest Dissertations & Theses Global | [**7**](https://search-proquest-com.ezproxy.lib.utah.edu/recentsearches.recentsearchtabview.recentsearchesgridview.scrolledrecentsearchlist.checkdbssearchlink_0:rerunsearch/5FE963EE313146A8PQ/None?site=pqdtglobal&t:ac=RecentSearches) |
| Select item 10 | **S10** | [noft(balance or posture or postural)](https://search-proquest-com.ezproxy.lib.utah.edu/recentsearches.recentsearchtabview.recentsearchesgridview.scrolledrecentsearchlist.checkdbssearchlink:rerunsearch/7BFFC2E3E2914F05PQ/None?site=pqdtglobal&t:ac=RecentSearches) | ProQuest Dissertations & Theses Global | [**82,613**](https://search-proquest-com.ezproxy.lib.utah.edu/recentsearches.recentsearchtabview.recentsearchesgridview.scrolledrecentsearchlist.checkdbssearchlink_0:rerunsearch/7BFFC2E3E2914F05PQ/None?site=pqdtglobal&t:ac=RecentSearches) |
| Select item 9 | **S9** | [noft((balance or gait or perturbation* or postural or posture or propriocept* or sensorimotor* or slip or slipping or stance or stand or standing or step or stepping or trip or tripping or walk or walking) NEAR/5 (assess* or dual-task or "dual task" or simulat* or task or tasks or test or tests or testing))](https://search-proquest-com.ezproxy.lib.utah.edu/recentsearches.recentsearchtabview.recentsearchesgridview.scrolledrecentsearchlist.checkdbssearchlink:rerunsearch/1D9AFD5EB6FD4D57PQ/None?site=pqdtglobal&t:ac=RecentSearches) | ProQuest Dissertations & Theses Global | [**16,307**](https://search-proquest-com.ezproxy.lib.utah.edu/recentsearches.recentsearchtabview.recentsearchesgridview.scrolledrecentsearchlist.checkdbssearchlink_0:rerunsearch/1D9AFD5EB6FD4D57PQ/None?site=pqdtglobal&t:ac=RecentSearches) |
| Select item 8 | **S8** | [noft(("virtual reality" or vr) NEAR/3 (balance or propriocept* or posture or postural))](https://search-proquest-com.ezproxy.lib.utah.edu/recentsearches.recentsearchtabview.recentsearchesgridview.scrolledrecentsearchlist.checkdbssearchlink:rerunsearch/EA1D5F1D60A94930PQ/None?site=pqdtglobal&t:ac=RecentSearches) | ProQuest Dissertations & Theses Global | [**22**](https://search-proquest-com.ezproxy.lib.utah.edu/recentsearches.recentsearchtabview.recentsearchesgridview.scrolledrecentsearchlist.checkdbssearchlink_0:rerunsearch/EA1D5F1D60A94930PQ/None?site=pqdtglobal&t:ac=RecentSearches) |
| Select item 7 | **S7** | [noft(ActiveStep or BESTest or CAREN or "Computer Assisted Rehabilitation Environment" or "center of pressure" or "centre of pressure" or "central sensorimotor integration task" or GVS or "galvanic vestibular stimulation" or "heel contact latenc*" or "lean-and-release" or "lean and release" or "lean & release" or "limits of stability" or "mini BEST" or miniBEST or (motion NEAR/3 (track* or control* or test*)) or MCT or "motor control test" or neurocom or posturograph* or "Push-and-release" or "push and release" or "push & release" or P&R or "P and R" or "release-from-lean" or "release from lean" or Simbex or "SMART EquiTest" or "stance slip" or SVS or "vestibular stimulation" or "support surface" or (trunk NEAR/5 (angle or sway)) or VEMP or "vestibular evoked myogenic potential*" or vibrat* or "waist belt" or (waist NEAR/5 belt)).ti,ab,kf. or ((ground or forceplate or "force plate" or platform or surface) NEAR/5 (moving or tilt or translation*))](https://search-proquest-com.ezproxy.lib.utah.edu/recentsearches.recentsearchtabview.recentsearchesgridview.scrolledrecentsearchlist.checkdbssearchlink:rerunsearch/3BCDFE657E464A58PQ/None?site=pqdtglobal&t:ac=RecentSearches) | ProQuest Dissertations & Theses Global | [**63,134**](https://search-proquest-com.ezproxy.lib.utah.edu/recentsearches.recentsearchtabview.recentsearchesgridview.scrolledrecentsearchlist.checkdbssearchlink_0:rerunsearch/3BCDFE657E464A58PQ/None?site=pqdtglobal&t:ac=RecentSearches) |
| Select item 6 | **S6** | [noft(perturbation*)](https://search-proquest-com.ezproxy.lib.utah.edu/recentsearches.recentsearchtabview.recentsearchesgridview.scrolledrecentsearchlist.checkdbssearchlink:rerunsearch/2B18C46E52154F15PQ/None?site=pqdtglobal&t:ac=RecentSearches) | ProQuest Dissertations & Theses Global | [**27,781**](https://search-proquest-com.ezproxy.lib.utah.edu/recentsearches.recentsearchtabview.recentsearchesgridview.scrolledrecentsearchlist.checkdbssearchlink_0:rerunsearch/2B18C46E52154F15PQ/None?site=pqdtglobal&t:ac=RecentSearches) |
| Select item 5 | **S5** | [noft(balance or "body sway" or "deep sensitivity" or imbalance or kinesthes* or perception* or propriocept* or propriocepsis or sensation* or ((body or kinesthetic or kinaesthetic or movement or position) NEAR/3 (discrimination or percept* or sense* or sensory or sensation*)) or ((balance or body or gait or posture or postural or musculoskeletal) NEAR/3 (abnormal* or adaption* or adjust* or control or difficult* or disequilibrium or disturbance* or entropy or equilibrium or error or impair* or instabilit* or oscillat* or orient* or react* or recover* or response* or restor* or stabili* or stance or steadiness or steady or sway* or unsteadiness or unsteady)) or (sensory NEAR/2 (function or functions)))](https://search-proquest-com.ezproxy.lib.utah.edu/recentsearches.recentsearchtabview.recentsearchesgridview.scrolledrecentsearchlist.checkdbssearchlink:rerunsearch/AC92542DBE4746D6PQ/None?site=pqdtglobal&t:ac=RecentSearches) | ProQuest Dissertations & Theses Global | [**304,199**](https://search-proquest-com.ezproxy.lib.utah.edu/recentsearches.recentsearchtabview.recentsearchesgridview.scrolledrecentsearchlist.checkdbssearchlink_0:rerunsearch/AC92542DBE4746D6PQ/None?site=pqdtglobal&t:ac=RecentSearches) |
| Select item 4 | **S4** | [noft(posture or postural)](https://search-proquest-com.ezproxy.lib.utah.edu/recentsearches.recentsearchtabview.recentsearchesgridview.scrolledrecentsearchlist.checkdbssearchlink:rerunsearch/A7DC289474254336PQ/None?site=pqdtglobal&t:ac=RecentSearches) | ProQuest Dissertations & Theses Global | [**8,156**](https://search-proquest-com.ezproxy.lib.utah.edu/recentsearches.recentsearchtabview.recentsearchesgridview.scrolledrecentsearchlist.checkdbssearchlink_0:rerunsearch/A7DC289474254336PQ/None?site=pqdtglobal&t:ac=RecentSearches) |
| Select item 3 | **S3** | [noft(dynamic NEAR/5 (balance or posture or postural))](https://search-proquest-com.ezproxy.lib.utah.edu/recentsearches.recentsearchtabview.recentsearchesgridview.scrolledrecentsearchlist.checkdbssearchlink:rerunsearch/C80658F2003E4ACBPQ/None?site=pqdtglobal&t:ac=RecentSearches) | ProQuest Dissertations & Theses Global | [**1,857**](https://search-proquest-com.ezproxy.lib.utah.edu/recentsearches.recentsearchtabview.recentsearchesgridview.scrolledrecentsearchlist.checkdbssearchlink_0:rerunsearch/C80658F2003E4ACBPQ/None?site=pqdtglobal&t:ac=RecentSearches) |
| Select item 2 | **S2** | [noft(reacti* or anticipatory or compensatory)](https://search-proquest-com.ezproxy.lib.utah.edu/recentsearches.recentsearchtabview.recentsearchesgridview.scrolledrecentsearchlist.checkdbssearchlink:rerunsearch/AE850F9336CB4C36PQ/None?site=pqdtglobal&t:ac=RecentSearches) | ProQuest Dissertations & Theses Global | [**267,314**](https://search-proquest-com.ezproxy.lib.utah.edu/recentsearches.recentsearchtabview.recentsearchesgridview.scrolledrecentsearchlist.checkdbssearchlink_0:rerunsearch/AE850F9336CB4C36PQ/None?site=pqdtglobal&t:ac=RecentSearches) |
| Select item 1 | **S1** | [noft((concuss* or commotio or "dementia pugulistica" or mtbi or mtbis or post-concuss* or postconcuss* or sub-concuss* or subconcuss* or tbi or tbis or ((brain or cerebell* or cerebral or cerebrovascular or cortical or encephalopath* or head) NEAR/3 (commotio* or concuss* or contusion* or impact* or injur* or postconcuss* or post-concuss* or posttraumatic or post-traumatic or trauma*))))](https://search-proquest-com.ezproxy.lib.utah.edu/recentsearches.recentsearchtabview.recentsearchesgridview.scrolledrecentsearchlist.checkdbssearchlink:rerunsearch/2C5ADDFD67204F73PQ/None?site=pqdtglobal&t:ac=RecentSearches) | ProQuest Dissertations & Theses Global | [**8,581**](https://search-proquest-com.ezproxy.lib.utah.edu/recentsearches.recentsearchtabview.recentsearchesgridview.scrolledrecentsearchlist.checkdbssearchlink_0:rerunsearch/2C5ADDFD67204F73PQ/None?site=pqdtglobal&t:ac=RecentSearches) |

------

Re-ran search on March 9, 2021

Applied Filters: 2020-01-01 - 2021-12-31	16
