## Supplemental Resource 3 for "Research Letter: Reactive balance responses after mild traumatic brain injury (mTBI): a scoping review"

**Supplemental Resource 3: Summary of Excluded Studies**

| **Study/Exclusion Reason** | **Population** | **N , groups** | **Testing Timeline** | **Perturbation Used** | **Outcome measures** | **Interpretation** |
| --- | --- | --- | --- | --- | --- | --- |
| Slobounov et al. ^1^  Wrong perturbation type (Continuous Perturbation) | Collegiate athletes | **N = 160**  Control = 122  mTBI = 38  2^nd^ mTBI = 9 | Baseline, 10,15,30 days after injury | VR moving room: anterior-posterior oscillations at 18 cm, 0.2 Hz;  lateral roll 10-30 deg, 0.2 hz | COP and coherence of COP to moving room | Posturography during head rotation is able to distinguish between control and TBI patients. Severity of neuro-ophthalmic deficit is correlated with severity of balance impairment. |
| Teel et al. ^2^  Wrong perturbation type  (Continuous Perturbation) | Collegiate athletes  18-24 years | **N = 121**  Control = 94  mTBI = 27 | Within 7-10 days of injury | VR moving room: rotation of room (yaw, pitch, and roll directions) | Score for each direction and composite of all directions; VR balance outcome scores based off of a 0–10 scale, with 10 being the best possible performance | The VR balance module has high sensitivity and specificity for detecting subacute balance deficits after concussive injury. |
| Teel et al. ^3^  Wrong perturbation type  (Continuous Perturbation) | Students and Athletes | **N = 88**  Control = 60  mTBI = 28 | Within 7-10 days of injury | VR moving room: rotation of room (yaw, pitch, and roll directions) | Score for each direction and composite of all directions; VR balance outcome scores based off of a 0–10 scale, with 10 being the best possible performance | VR balance module can be used as a valid clinical tool |

| **Study/Exclusion Reason** | **Population** | **N , groups** | **Testing Timeline** | **Perturbation Used** | **Outcome measures** | **Interpretation** |
| --- | --- | --- | --- | --- | --- | --- |
| Wright et al. ^4^  Wrong perturbation type  (Continuous Perturbation) | College students | **N = 67**  Control = 56  mTBI = 11 |  | VR rotating scene about the subject’s roll axis at 60 deg/s | COP velocity, standard deviation in anterior-posterior (AP) direction and the mediolateral (ML) direction, and COP sway area. | COP sway area, velocity and standard deviation in the AP direction were significantly different between controls and concussed. |
| Wright et al. ^5^  Wrong perturbation type  (Continuous Perturbation) | College students  21.7 ± 3.5 years | **N = 72**  Control = 58  mTBI = 14 | Within 6 months of injury | VR rotating scene about the subject’s roll axis at 60 deg/s | COP Sway, medial-lateral standard deviation | The concussed group had greater COP sway area than healthy |
| Greffou et al. ^6a^  Wrong perturbation type  (Continuous Perturbation) | Children  9 – 18 years | **N= 73**  Control: 36  Low Symptoms: 19  Moderate Symptoms: 18 | 2 weeks, 12 weeks and 1 year post-injury | Virtual reality tunnel, oscillating at 3 different frequencies | vRMS – root mean square of total body velocity in the horizontal plane,  sway amplitude (anterior-posterior displacement) | Children with moderate symptoms post mTBI show increased postural instability with exposure to optic flow stimuli up to 12 weeks post mTBI. Appears to be no difference at 12 months post. |

^a^This study was also included in a dissertation document Greffou et al. ^7^

| **Study/Exclusion Reason** | **Population** | **N , groups** | **Testing Timeline** | **Perturbation Used** | **Outcome measures** | **Interpretation** |
| --- | --- | --- | --- | --- | --- | --- |
| Cripps et al.  ^8^  Wrong perturbation type  (Continuous Perturbation) | Intercollegiate, collegiate, club sport athletes  17.1 ± 3 years | **N= 14**  Control = 7  mTBI = 7 | 24-48 hours and 10 days post injury | Visual perturbation (radial optic flow) during SOT and mCTSIB | SOT: equilibrium score, sensory analysis ration  mCTSIB: mean COG sway velocity, composite score, and COG alignment | Those with mTBI improved postural stability in presence of continuous visual perturbation. |
| Slobounov et al. ^9^  Wrong perturbation type (Continuous Perturbation) | Collegiate Athletes  18-25 years | **N = 55**  BL: 55  Post-mTBI: 10 | BL and  3, 10 , 30 days post injury | Virtual reality room, forward-backward and lateral-roll room  movement presented a)while subject is quietly standing  b)while subject is moving | Center of pressure, coherence between moving room and postural responses | Responses to visual field motion induced postural dysfunctions at least 30 days post mTBI. These findings support both that a concussion changes postural responses and the impairment lasts at least 30 days post mTBI. |

| **Study/Exclusion Reason** | **Population** | **N , groups** | **Testing Timeline** | **Perturbation Used** | **Outcome measures** | **Interpretation** |
| --- | --- | --- | --- | --- | --- | --- |
| Slobounov et al. ^10^  Wrong perturbation type (Continuous Perturbation) | Collegiate Athletes  18-25 years | **N = 48**  BL: 48  Post-mTBI: 8 | BL and  3,10, 30 days post injury | Virtual reality room, forward-backward and lateral-roll room movement presented while subject is quietly standing | Center of pressure, coherence between moving room and postural responses | None of the concussed subjects were able to preserve balance while viewing a moving room on day 3 post-injury and some continued to have deficits at 30 days post-concussion |
| Allexandre et al.  ^11^  Wrong perturbation type  (Continuous Perturbation) | Community  Control: 41 ± 15.6  TBI: 53.8 ± 7.8 | **N = 18**  Control: 6  TBI: 12  (5 mild, 2 moderate, 5 severe) | 1.6-57.8 years post injury, tested once | NeuroCom balance platform, 5 blocks of 10 random anterior-posterior sinusoidal translations (0.5 Hz) at low (0.5 cm) and high (2 cm) amplitude | High amplitude backward perturbations only: center of pressure displacement, electroencephalography (P1N1 amplitude, P1 latency,N1 latency),  Electromyography (onset time) | TBI was worse at maintaining balance in response to an unpredictable perturbation and had impaired cortical response (greater COP displacement and lower N1 amplitude). |
| Bailey et al.^12^ | Young adults  SRC: 21.9 ± 2.8 years  Control: 21.5 ± 1.2 years | **16**  SRC: 8  Control: 8 | Within 6 months of concussion, after clearance for RTP | Sinusoidal platform rotations around pitch axis at 0.8 Hz | Margin of stability, anchoring index (absolute pitch of head to external lab coordinate system and relative pitch of head) | Athletes with SRC within 6 months had greater reduction in postural control during perturbation trials with eyes closed. |

| **Study/Exclusion Reason** | **Population** | **N , groups** | **Testing Timeline** | **Perturbation Used** | **Outcome measures** | **Interpretation** |
| --- | --- | --- | --- | --- | --- | --- |
| Caccese et al.^13^ ^b^ 2020  Wrong perturbation type  (Continuous Perturbation) | 2 weeks – 6 months post-concussion: 21 ± 3 years  >1 year: 21 ± 1 years  Controls: 22 ± 3 years | **N = 52**  Recent Concussion: 13  >1 year: 13  Control: 26 | >2 weeks post-concussion | Sinusoidal translation of visual scene at 0.2 Hz, GVS at 0.36 Hz, and tendon vibration at 0.28 Hz | Center of mass gain | The recent concussion group and >1 year group had higher gains to visual and vestibular stimuli than the control group. |
| Rosen et al.^14^  Wrong perturbation type  (Continuous Perturbation) | Service Members  34.15 ± 7.75 years | **112**  3-6 months post mTBI: 14  6-12 months post mTBI: 22  >12 months: 76 | > 3 months after mTBI | Immersive balance tests in Computer Assisted Rehabilitation Environment (CAREN): Shark Hunt and Balance Cubes | Time to complete immersive balance tests, clinical score of SOT and FGA | SOT and FGA had the strongest relationships to immersive balance tests. Immersive balance tasks may be affective to further examine balance post-mTBI. |

^b^This study was also included in a journal article Caccese et al.^15^

| **Study/Exclusion Reason** | **Population** | **N , groups** | **Testing Timeline** | **Perturbation Used** | **Outcome measures** | **Interpretation** |
| --- | --- | --- | --- | --- | --- | --- |
| Chow et al.  ^16^  Wrong injury severity  (No mTBI’s included, only healthy) | 18-35 years | **N = 72**  Control: 31  Rugby Players: 41 | No injuries in past 12 months, tested once | NeuroCom, motor control test (anterior-posterior perturbations) | Latency of force response, composite response time | Composite response time was longer in rugby group indicating slower lower extremity muscle reactions to postural perturbations. This could be due to sport specific training overriding reflexive postural responses. |
| Damiano et al.  ^17^  Wrong injury severity  (Greater than mild severity) | 19-44 years | **N = 24**  Control: 12  TBI: 12 | At least 6 months post injury and after an 8 week daily exercise program (rapid, mild resistance elliptical) | NeuroCom, motor control test (MCT) | latency of response | No difference between groups in latency of response during MCT. After intervention latency during MCT improved providing evidence for improvement of reactive balance with 8 weeks of rapid, coordinated reciprocal movements of arms and legs. |

| **Study/Exclusion Reason** | **Population** | **N , groups** | **Testing Timeline** | **Perturbation Used** | **Outcome measures** | **Interpretation** |
| --- | --- | --- | --- | --- | --- | --- |
| Pilkar et al.  ^18^  Wrong injury severity  (Greater than mild severity) | Adult  46 years | **N = 1**  Severe TBI | At least 6 months post injury  Two time points: before intervention and after 3 sessions of biofeedback intervention | NeuroCom, anterior-posterior perturbations (20 trials with, 10 without) | RMS electromyography, participant perception of perturbation (can the participant detect the movement) | After intervention the participant with TBI improved ability to detect perturbations. Biofeedback intervention could be used to improve perception and neuromuscular response to perturbations. |
| Pilkar et al. ^19^  Wrong injury severity (Greater than mild severity and continuous perturbations) | Adult | **N = 14**  TBI = 9  Control = 5 | Baseline and after 4 weeks of computerized biofeedback intervention | Neurocom sinusoidal perturbations in AP direction with amplitude of 10mm at 1hz | EMG – anticipatory (APA) and compensatory postural adjustments (CPA) | TBI had larger magnitude CPA responses and more inconsistent APA responses compared to controls. This indicates that APA and CPA may be affected by TBI. |
| Ustinova et al.  ^20^  Wrong injury severity (Greater than mild severity and continuous perturbation) | Rehabilitation centers  33.4 ± 9.1 years | N = 30  Control = 15  TBI = 15 | At least 6 months after injury | VR sinusoidal pitch at 15 deg/sec | Amplitude and velocity peaks of COM | Those with TBI had greater COM sway amplitude and velocity in the sagittal and frontal planes than control. |

| **Study/Exclusion Reason** | **Population** | **N , groups** | **Testing Timeline** | **Perturbation Used** | **Outcome measures** | **Interpretation** |
| --- | --- | --- | --- | --- | --- | --- |
| Tanis et al.  ^21^  Wrong injury severity  (Greater than mild severity and continuous perturbations) | 45-60 years | Control = 4  TBI = 7 | ~ 4 years post injury  Baseline and 4 weeks after dynamic computerized biofeedback intervention (CBB). | Participants asked if they could perceive perturbation – sinusoidal platform translation in AP direction at 3 different frequencies; Dynamic balance assessment: asked to stand as still as possible in response to sinusoidal perturbations in AP direction at 3 diff frequencies and amplitudes | Hit: correctly ID  Miss: did not detect  False alarm: ID but no perturbation; RMS COP and RMS COP velocity in AP direction | HC group  displayed stable postural responses compared to TBI. TBI less likely to detect smaller perturbations that HC could perceive. Post CBB intervention TBI showed improvements in postural stability and balance. |
| Tefertiller et al.  ^22^  Wrong injury severity  (Greater than mild severity) | 48.1 ± 12.1 years | Moderate to severe TBI  VR program = 31  Traditional home exercise program (HEP) = 32 | At least 1 year post injury  Baseline, 6 weeks, 12 weeks, 24 weeks | BESTest – reactive postural responses | Composite score ranging from 0 -108 | This study found no between-group differences in balance in  individuals with chronic TBI who received VR in comparison to a  traditional HEP. |

| **Study** | **Reason** |
| --- | --- |
| Agostini et al. ^23^ | Non-external balance disturbance: Head turning |
| Chang ^24^ | Non-external balance disturbance: Limits of stability testing and static balance  Injury Severity: greater than mild, cerebellar blood flow abnormalities |
| Dalla Toffola et al. ^25^ | Not available in English |
| Gagnon et al. ^26^ | No postural outcome measure: amount of weight used to disturb balance |
| Habib Perez et al. ^27^ | Injury severity: Did not test any mTBI |
| Hays et al. ^28^ | Injury severity: greater than mild  No postural outcomes reported, only BESTest scores |
| Hays et al. ^29^ | Injury severity: moderate to severe TBI with ongoing balance impairments |
| Hays et al. ^30^ | Injury severity: history of TBI with balance impairments  Non-external balance disturbance: only sensory subscale of BESTest included |
| Wilson et al. ^31^ | Non-external balance disturbance - mCTSIB |

**References**

1.	Slobounov S, Slobounov E, Sebastianelli W, Cao C, Newell K. Differential rate of recovery in athletes after first and second concussion episodes. *Neurosurgery*. 2007;61(2):338-344. doi:10.1227/01.NEU.0000280001.03578.FF

2.	Teel EF, Gay MR, Arnett PA, Slobounov SM. Differential sensitivity between a virtual reality balance module and clinically used concussion balance modalities. *Clinical Journal of Sport Medicine*. 2016;26(2):162-166. doi:10.1097/JSM.0000000000000210

3.	Teel EF, Slobounov SM. Validation of a virtual reality balance module for use in clinical concussion assessment and management. *Clinical Journal of Sport Medicine*. 2015;25(2):144-148. doi:10.1097/JSM.0000000000000109

4.	Wright WG, McDevitt J, Tierney R, Haran FJ, Appiah-Kubi KO, Dumont A. Assessing subacute mild traumatic brain injury with a portable virtual reality balance device. *Disability and Rehabilitation*. 2017;39(15):1564-1572. doi:10.1080/09638288.2016.1226432

5.	Wright WG, Tierney RT, McDevitt J. Visual-vestibular processing deficits in mild traumatic brain injury. *Journal of Vestibular Research: Equilibrium and Orientation*. 2017;27(1):27-37. doi:10.3233/VES-170607

6.	Greffou S, McKerra M, Gagnon I, Forget R, Faubert J. Prolonged alterations of visually-driven balance control in children following a mild traumatic brain injury: A virtual reality study. *Brain Injury*. 2014;28(5-6):679-679.

7.	Greffou S. *A virtual reality approach to the study of visually driven postural control in developing and aging humans*. 2013. <https://papyrus.bib.umontreal.ca/xmlui/handle/1866/10815>

8.	Cripps A, Livingston S, Jiang Y, et al. Visual perturbation impacts upright postural stability in athletes with an acute concussion. *Brain Injury*. 2018;32(12):1566-1575. doi:10.1080/02699052.2018.1497812

9.	Slobounov S, Slobounov E, Newell K. Application of Virtual Reality Graphics in Assessment of Concussion. *CyberPsychology & Behavior*. 2006;9(2):188-191. doi:10.1089/cpb.2006.9.188

10.	Slobounov S, Tutwiler R, Sebastianelli W, Slobounov E. Alteration of Postural Responses to Visual Field Motion in Mild Traumatic Brain Injury. *Neurosurgery*. 2006;59(1):134-193. doi:10.1227/01.neu.0000243292.38695.2d

11.	Allexandre D, Hoxha A, Handiru VS, Saleh S, Selvan SE, Yue GH. Altered Cortical and Postural Response to Balance Perturbation in Traumatic Brain Injury -An EEG Pilot Study∗. Institute of Electrical and Electronics Engineers Inc.; 2019:1543-1546.

12.	Bailey HGB, Kirk C, Mills RS, Foster RJ. A preliminary cross-sectional assessment of postural control responses to continuous platform rotations following a sport-related concussion. *Gait & Posture*. 2020/09/01/ 2020;81:213-217. doi:<https://doi.org/10.1016/j.gaitpost.2020.08.105>

13.	Caccese JB, Vanderlinde Santos F, Yamaguchi FK, Jeka JJ. Persistent visual and vestibular impairments for Postural control following concussion. American Academy of Neurology; 2020:S14-S15.

14.	Rosen KB, Delpy KB, Pape MM, Kodosky PN, Kruger SE. Examining the Relationship Between Conventional Outcomes and Immersive Balance Task Performance in Service Members With Mild Traumatic Brain Injury. *Military Medicine*. 2021;186(5-6):577-586. doi:10.1093/milmed/usaa578

15.	Caccese JB, Santos FV, Yamaguchi FK, Buckley TA, Jeka JJ. Persistent Visual and Vestibular Impairments for Postural Control Following Concussion: A Cross-Sectional Study in University Students. *Sports Medicine*. 2021/04/21 2021;doi:10.1007/s40279-021-01472-3

16.	Chow GCC, Chung JWY, Ma AWW, Macfarlane DJ, Fong SSM. Sensory organisation and reactive balance control of amateur rugby players: A cross-sectional study. *European Journal of Sport Science*. 2017;17(4):400-406. doi:10.1080/17461391.2016.1257656

17.	Damiano DL, Zampieri C, Ge J, Acevedo A, Dsurney J. Effects of a rapid-resisted elliptical training program on motor, cognitive and neurobehavioral functioning in adults with chronic traumatic brain injury. *Experimental Brain Research*. 2016;234(8):2245-2252. doi:10.1007/s00221-016-4630-8

18.	Pilkar R, Arzouni N, Ramanujam A, Chervin K, Nolan KJ. Postural responses after utilization of a computerized biofeedback based intervention aimed at improving static and dynamic balance in traumatic brain injury: A case study. Institute of Electrical and Electronics Engineers Inc.; 2016:25-28.

19.	Pilkar R, Ibironke O, Ehrenberg N, Nolan KJ. Anticipatory and Compensatory Postural Responses during Perturbed Standing in Individuals with Traumatic Brain Injury. Institute of Electrical and Electronics Engineers Inc.; 2019:5080-5083.

20.	Ustinova KI, Silkwood-Sherer DJ. Postural perturbations induced by a moving virtual environment are reduced in persons with brain injury when gripping a mobile object. *Journal of Neurologic Physical Therapy*. 2014;38(2):125-133. doi:10.1097/NPT.0000000000000035

21.	Tanis D, Pilkar R, Ibironke O, Nolan KJ. Improving Perception of Perturbations and Balance Using a Novel Dynamic Computerized Biofeedback Based Intervention after Traumatic Brain Injury. *Conference proceedings : Annual International Conference of the IEEE Engineering in Medicine and Biology Society IEEE Engineering in Medicine and Biology Society Annual Conference*. 2018;2018:5594-5597. doi:10.1109/EMBC.2018.8513663

22.	Tefertiller C, Hays K, Natale A, et al. Results From a Randomized Controlled Trial to Address Balance Deficits After Traumatic Brain Injury. *Archives of Physical Medicine and Rehabilitation*. 2019;100(8):1409-1416. doi:10.1016/j.apmr.2019.03.015

23.	Agostini V, Chiaramello E, Bredariol C, Cavallini C, Knaflitz M. Postural control after traumatic brain injury in patients with neuro-ophthalmic deficits. *Gait and Posture*. 2011;34(2):248-253. doi:10.1016/j.gaitpost.2011.05.008

24.	Chang S-T. Evaluation of postural function with abnormal cerebellar blood flow in patients with chronic mild brain injury. *Brain Injury*. 2014;28(5-6):517-878.

25.	Dalla Toffola E, Quarenghi A, Gasperini G, Losio L, Pistorio A, Salvi GP. Alterazioni posturali residue in esiti di trauma cranico [Residual postural changes after cranial trauma]. *G Ital Med Lav Ergon*. 1999;21(3):226-232.

26.	Gagnon I, Swaine B, Friedman D, Forget R. Children show decreased dynamic balance after mild traumatic brain injury. *Archives of Physical Medicine and Rehabilitation*. 2004;85(3):444-452. doi:10.1016/j.apmr.2003.06.014

27.	Habib Perez OD. *Inter-limb Coordination in Balance Control: Implications for understanding Balance after Traumatic Brain Injury*. 2018. <https://tspace.library.utoronto.ca/handle/1807/89690>

28.	Hays K, Tefertiller C, Ketchum J, et al. The use of the balance evaluation systems test for individuals with chronic traumatic brain injury. *Journal of Head Trauma Rehabilitation*. 2018;33(3):E81-E82.

29.	Hays K, Tefertiller C, Ketchum JM, et al. Balance in chronic traumatic brain injury: correlations between clinical measures and a self-report measure. *Brain Injury*. 2019;33(4):435-441. doi:10.1080/02699052.2019.1565900

30.	Hays K, Tefertiller C, O'Dell D, Natale A, Ketchum J. Comparison of the sensory organization test and the balance evaluation systems test in traumatic brain injury. *Brain Injury*. 2017;31(6-7):922-922.

31.	Wilson K, Dick T, Hunt A, et al. Postural stability in youth athletes postconcussion. *Brain Injury*. 2016;30(5-6):711-711.
